# Altered Heterogeneity of Ageing Lung Endothelium is a Hallmark of Idiopathic Pulmonary Fibrosis

**DOI:** 10.1101/2022.03.08.22272025

**Authors:** Eamon C. Faulkner, Adam A. Moverley, Simon P. Hart, Leonid L. Nikitenko

## Abstract

**Background:** Older age is the main risk factor for chronic lung diseases including idiopathic pulmonary fibrosis (IPF). Halting or reversing progression of IPF remains an unmet clinical need due to limited knowledge of underlying mechanisms. The lung circulatory system, composed of blood (pulmonary and bronchial) and lymphatic vessels networks, has been implicated in IPF pathophysiology in elderly people, based solely on reports of altered density and increased permeability of vessels.

**Aim:** We aimed to define heterogeneity and IPF-associated changes of lung endothelial cells (EC or endothelium) by comparing gene expression in tissues from elderly people - transplant donors and recipients with IPF.

**Methods:** Single-cell RNA sequencing (scRNAseq) datasets of “ageing lung” tissues were selected only from those publicly available sources that contain age-matching samples for both groups (49- 77 years old donors and IPF patients; nine pairs in total), integrated and compared. Findings were validated by immunohistochemistry using EC-specific markers.

**Results:** The generation of integrated single-cell maps of ageing lung tissues revealed 17 subpopulations of endothelium (12 for blood and 5 for lymphatic vessels, including 9 novel), with distinct transcriptional profiles. In IPF lung, the heterogeneity of ageing lung endothelium was significantly altered - both in terms of cell numbers (linked to disease- related changes in tissue composition) and differentially expressed genes (associated with fibrosis, inflammation, differentiation and vasodilation) in individual pulmonary, bronchial and lymphatic EC subpopulations.

**Conclusions:** These findings reveal underappreciated extent of heterogeneity and IPF-associated changes of ageing lung endothelium. Our data suggest direct involvement of specific subpopulations of ageing lung endothelium in IPF pathophysiology, uncovering cellular and molecular targets which may have potential diagnostic, prognostic and therapeutic relevance. This study creates a conceptual framework for appreciating the disease-specific heterogeneity of ageing lung endothelium as a hallmark of IPF.

## Background

Increasing age is the main risk factor for diagnosis and prognosis of major non- communicable lung diseases, including lung cancer, chronic obstructive pulmonary disease (COPD) and idiopathic pulmonary fibrosis (IPF) [1, 2]. IPF is the most common, progressive and lethal interstitial lung disease with a prevalence of 33 to 450 per million people globally [3]. IPF progression leads to declining quality of life, respiratory failure and eventually lung transplantation or death. While the number of older people suffering from this disease is steadily increasing and histopathological pattern of usual interstitial pneumonia (UIP), hallmarks of ageing, particularly cellular senescence and extracellular matrix (ECM) dysregulation, are prominent in the IPF lung, the aetiology of this disease remains unclear [2, 4–7]. Detailed understanding of cellular and molecular mechanisms underlying IPF- associated changes in ageing human lung will be essential for diagnostics or prognostics of IPF, and for developing effective therapies to halt or reverse disease progression.

Several lines of evidence implicate a role for the circulatory system, composed of blood (pulmonary and bronchial) and lymphatic vessel networks, in IPF [7–18]. Some studies report structural or functional changes in blood or lymphatic vessels, which are lined with a monolayer of endothelial cells (EC or endothelium), although often without a comparison between IPF patients and age-matched controls [10, 15, 17]. The precise contribution of specialized lung vessel subtypes [19, 20] and distinct EC subpopulations that comprise them to the pathophysiology and progression of IPF remain insufficiently characterized [21, 22].

Recent advances in single-cell RNA sequencing (scRNAseq) [23] have allowed characterisation of cellular heterogeneity in healthy and diseased lung, resulting in the generation of a human lung cell atlas (HLCA; [22]) and the IPF cell atlas [24]. Studies of heterogeneity of human lung endothelium have reported up to nine subpopulations of EC in healthy and/or diseased lung [22, 25, 26]. These reports utilised data from either a diverse population of subjects (patients with COPD, IPF or other lung diseases, commonly placed together for comparison) with a range of ages (varying from 21 to 80 years in some cases), ethnicities and tobacco smoker statuses, or used samples from different areas of the lung, e.g. distal lung parenchyma [27], longitudinal sections [25] or multiple lobes [28]. One recent study generated a cell atlas of ageing lungs in COPD, comparing age-matched healthy subjects and patients, and identifying 6 subpopulations of EC [21]. However, currently there is still a major gap in knowledge about properties and roles of individual cell subpopulations composing ageing human lung endothelium in other chronic lung diseases, including IPF. Our report is the first to define (characterize, compare and contrast) the heterogeneity and disease-associated changes of ageing lung endothelium in IPF patients and age-matched older healthy subjects.

## Results

### Integrated Single Cell Map of Ageing Human Lungs Displays Endothelial Cell Contribution in Disease

To characterise ageing human lung endothelium in IPF and to account for its intra- and inter-lung heterogeneity within and between individuals [29], we performed comparative analysis of scRNAseq data from lung tissue samples selected only from age-matched (63± 14 years old) transplant donors and recipients with IPF. Specifically, we selected data from only those studies (cohorts) that contained samples for both conditions (transplant donors and IPF patients). From all publicly available scRNAseq datasets of human IPF lungs (4 in total, to our knowledge; [25, 27, 28, 30, 31]), only two, representing independent cohorts of subjects/individuals of different ethnic backgrounds, with controlled sample locations, fulfilled these selection criteria [25, 27] and therefore were used for generating a single dataset of ageing human lungs for integrative comparative scRNAseq analysis (Table 1; Figure 1A; for full details on dataset selection see Additional file 1: Detailed methods / Dataset Selection).

**Figure 1.**
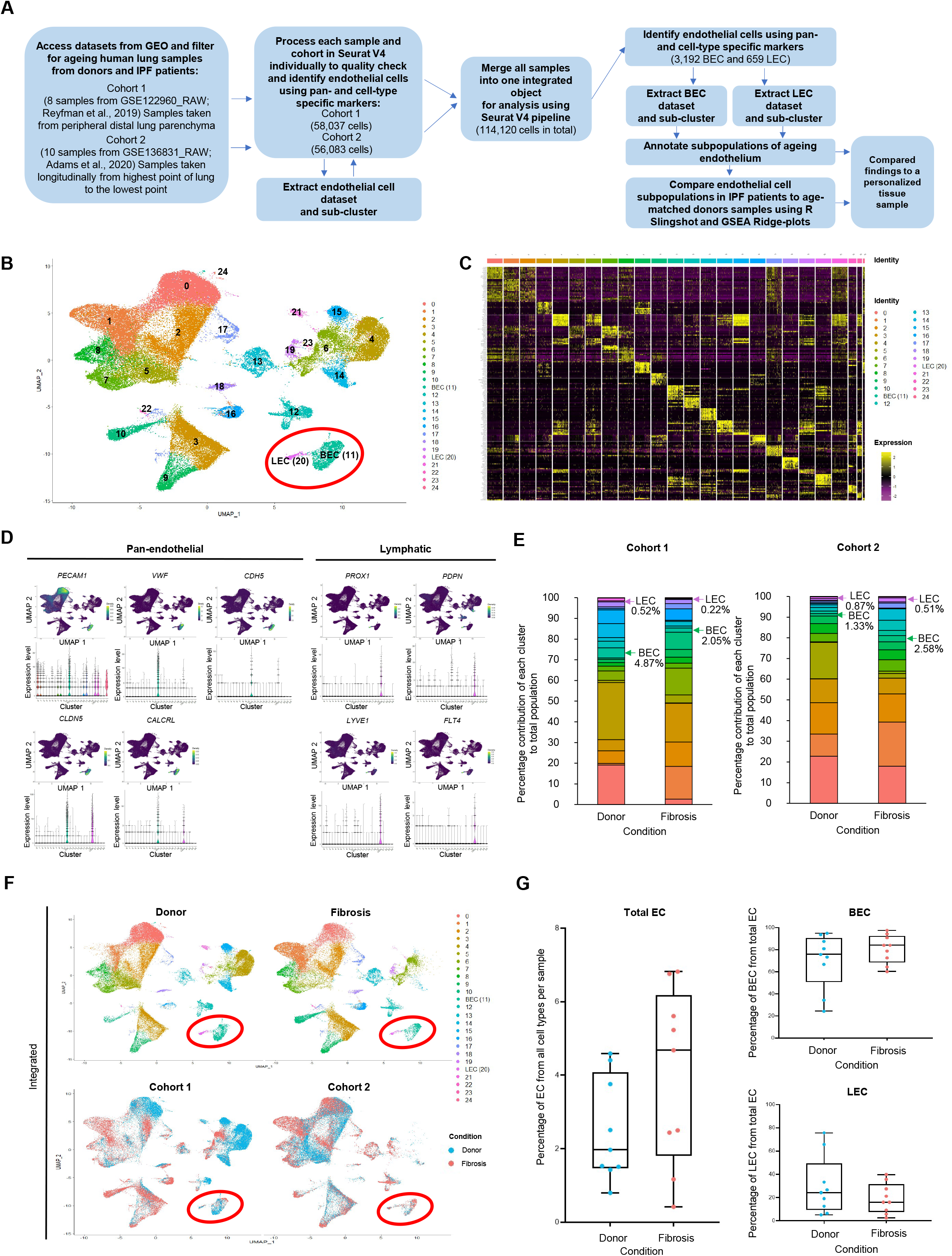
Cell type-specific composition of ageing human lung tissues from donors and IPF patients from two independent cohorts. (A) Schematic of methodology for samples integration and comprehensive data analysis. Selected data from two studies were grouped into two cohorts (18 samples in total; see Table 1; [25, 27]) and subjected to quality control (Additional file 2: Figures S1, 2B) and merging into an integrated Seurat V4 object for subsequent analysis. The resolution was determined using the clustree R package, with 0.5 value used for this analysis and throughput the study to avoid over-clustering. Endothelial cell (EC) clusters were identified using pan-endothelial and lymphatic-specific markers. 3192 blood vessel endothelial cells (BEC) and 659 lymphatic vessel endothelial cells (LEC) were identified, and sub-clustered for their characterization and comparison in donor and IPF samples. Full details can be found in Additional file 1: Detailed methods. **(B)** UMAP representation of all cells and cell clusters from all 18 samples (pooled). Clusters were numbered, annotated and labelled according to their detected gene markers. Specific details about each cell population can be found in Additional file 2: Figure S2 and Additional file 3, Table S1. Two clusters (LEC and BEC) are labelled with a red ellipse. **(C)** Heatmap of top ten differentially expressed genes by cells in the cluster. Each column represents the average expression for a cell, hierarchically grouped by cell type. Gene expression values are scaled from 2 (yellow) to -2 (purple) across rows within each cell type. **(D)** Violin plots and density plots representation of expression data for pan-EC and LEC-specific markers expression in all identified clusters. Density plot expression values are scaled from between 0.04 and 1 (*yellow –* high expression) to 0 (*purple* – low expression). **(E)** Stacked bar chart of percentage contribution of each cluster to total lung population, split by cohort and condition - donor or fibrosis (IPF). Colour keys for cluster identity is the same as in **B**. Percent contributions of 3,192 BEC and 659 LEC to the total cell population in ageing human lung are shown with *arrows*. Specific details for each cell population can be found in the Additional file 3: Table S3. **(F)** UMAP representation of all cells and clusters from all 18 samples (pooled), split by condition (donor or fibrosis) and labelled by cluster (top panel), or split by cohort and labelled by condition (*blue* – donor; *red* – fibrosis; bottom panel). **(G)** Box and whiskers plots of EC percentage all cell types in the lung (*left*) or blood or lymphatic endothelial cells (BEC and LEC respectively) from total EC per sample in individual lung samples percentage from all other cell types in individual lung samples (*right*), split by condition (*blue* – donor; *red* – IPF/fibrosis). The lines within the boxes denotes the medians, the ‘box’ contains the 25th to 75th percentiles and the ‘whiskers’ mark the range of the data. Statistical analysis was done using Shapiro-Wilk test (p=0.162 for donor and p=0.367 for fibrosis) and unpaired t-test (p = 0.139). Means ± standard deviation (2.501±1.407 for donor and 3.959±2.388 for fibrosis). UMAP - Uniform Manifold Approximation and Projection.

**Table 1.**
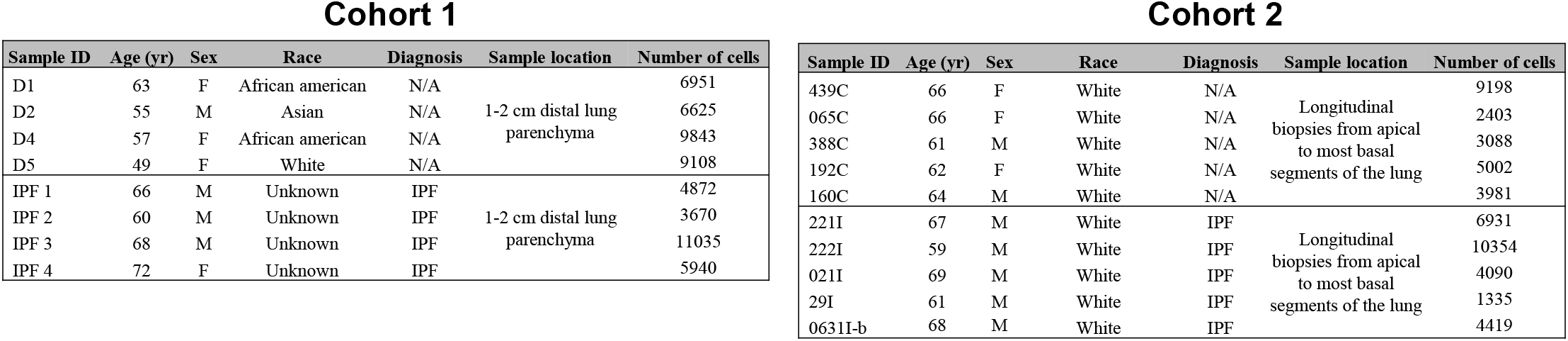
Characteristics of aging human lung tissue samples split by cohort. Cohort 1 (8 samples) was from GSE122960_RAW (Reyfman et al., 2019) and cohort 2 (10 samples) was from GSE136831 (Adams et al., 2020). M = male, F = female, IPF = idiopathic pulmonary fibrosis, N/A = not applicable.

The integration of the two datasets resulted in generation of a multi cohort single cell map revealing 25 distinct populations (Figure 1A-B, Additional file 2: Figure S1), based on specific gene expression patterns (Figures 1C, Additional file 2: Figure S2, Additional file 3: Table S1) that include known cell type-specific transcriptomic markers [22, 25–27]. Identities of two closely related blood and lymphatic vessel EC (BEC and LEC respectively) populations (Figure 1B) were confirmed by detailed analysis of expression of a panel of pan- EC and lymphatic EC-specific markers (Figure 1D).

Quantitative analysis of cell numbers for each of 25 populations in age-matched groups revealed IPF-associated changes in ageing human lungs in both cohorts (Figure 1E). The analysis reflected differences in location of tissue samples (Table 1), matching contribution of different cell types to the total human lung cell population and alterations in IPF lung reported in age non-matching studies [21, 22, 25–27, 30, 32] (Additional file 3: Table S2). Also, it uncovered statistically insignificant changes in proportions of ageing human lung BEC and LEC populations between cohorts and conditions (Figure 1E, Additional file 3: Table S3), which is in agreement with similar analysis in age non-matching study of IPF lungs [25]. Data integration revealed comparable spatial profiles of ageing human lung EC populations in two conditions and two cohorts (Figure 1F) and their similar contribution in donors and IPF patients (Figure 1G).

### Blood Vessel Endothelium in Ageing Human Lung Exhibits Disease-Specific Heterogeneity and Alterations in Cell Numbers of Individual Subpopulations

Unsupervised clustering of ageing human lung BEC alone (n=3192) identified 12 distinct subpopulations (Figures 1A, 2A) based on their differentially expressed genes (DEG; Figures 2B, Additional file 2: Figure S3). Expression analysis of pan-EC markers and LEC-specific genes confirmed their identities as BEC (Figure 2C). The analysis of top 10 DEG for each subpopulation (110 in total) determined their selection for inclusion (or not) in 10 signatures of ageing human lung BEC subpopulations (Figures 2B, D, Additional file 2: Figure S3, S4; Additional file 3: Figure Table S4). 49 selected subpopulation-specific DEG contribute to distinct transcriptional profiles for subpopulations 1-4 and 6-11 (Figure 2D). Subpopulations zero and 5 lack obvious markers or signatures (Figures 2D, Additional file 2: Figure S5).

**Figure 2.**
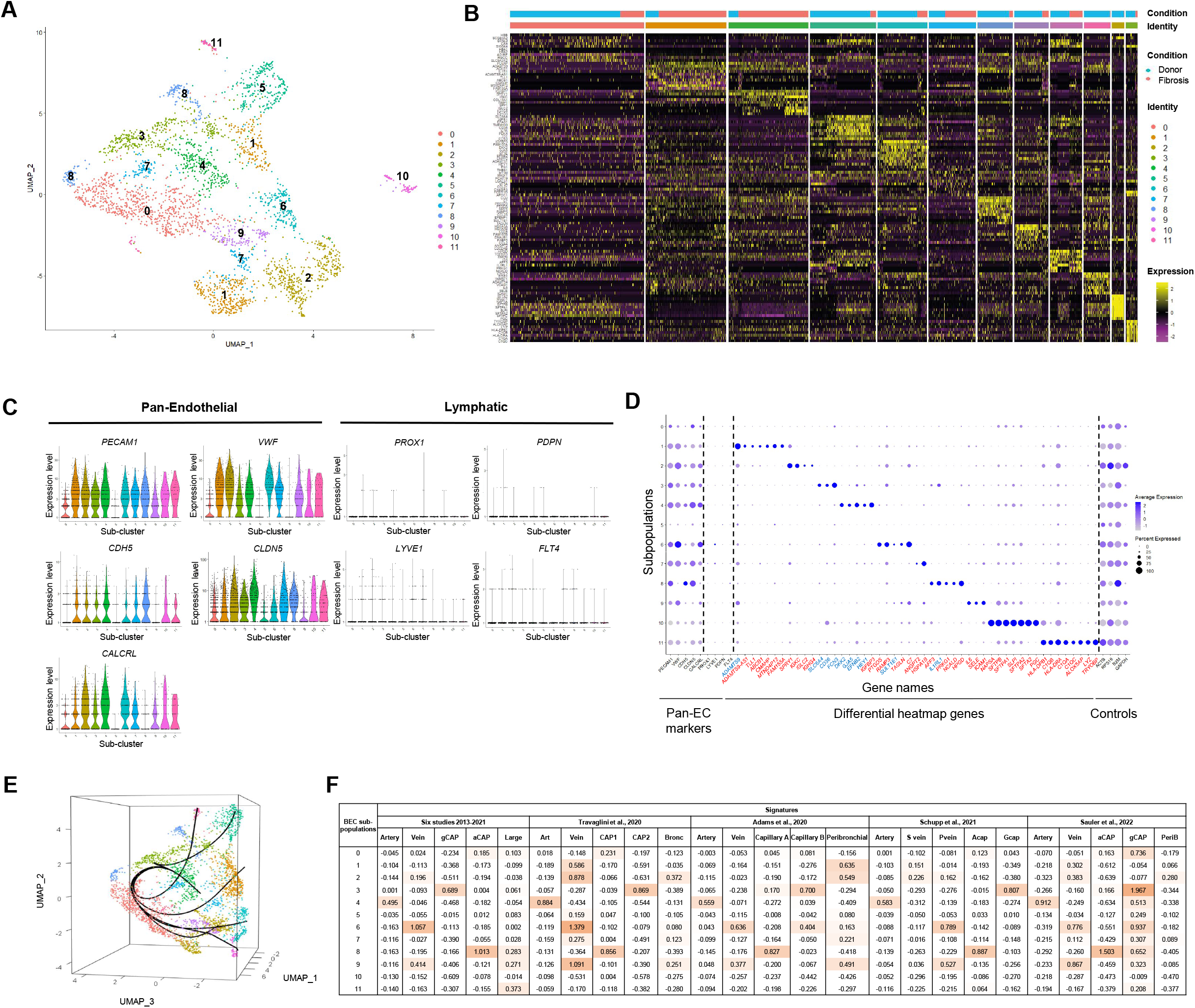
Cell subtype-specific EC subpopulation composition of ageing human lung blood vessel endothelial cell cluster from donors and IPF patients from two independent cohorts. To comprehensively decipher transcriptional signatures of ageing human lung blood vessel endothelial cell (BEC) in IPF, we subtracted scRNAseq data for BEC (n=3192) from the total lung cell populations from integrated dataset (two cohorts and both conditions, see Figure 1A), and sub-clustered them. **(A)** UMAP representation of all cells and cell subpopulations from BEC cluster (from all 18 pooled samples) which are presented in the Figure 1. Subpopulations were labelled 0-11 according to their signatures (Additional file 2: Figure S3). Note that, although subpopulations 1, 7 and 8 visually present with two potential subgroups each on this UMAP, each of two subgroups are not significantly different from each other in the described clustering parameters (Additional file 1: Detailed methods). In subpopulation 1, this is likely to be due to the limitations of visualisation of 2D UMAP compared to 3D UMAP, which shows as an individual cluster (see also Figure 2E, Additional file 4: Video S1). In subpopulation 7 and 8, 3D UMAP reveals the shift of one subgroup towards subpopulation zero (see also Figure 2E; Additional file 4: Video S1). **(B)** Heatmap of top 10 differentially expressed genes (DEG) by subpopulation. Each column represents the average expression for a cell, hierarchically grouped by condition (donor or fibrosis) and cell type. Gene expression values are scaled from 2 (yellow) to -2 (purple) across rows within each cell type. Specific details about each cell subpopulation can be found in Additional file 2: Figures S3 - 5, and Additional file 3: Table S4. All cell clusters can be separated with high confidence (Additional file 3: Table S5). **(C, D)** The analysis of the top 10 DEG for each subpopulation (120 in total) was performed alongside tests for expression of pan-EC and LEC markers and using violin plots to test their subpopulation-specific expression for determining their selection for inclusion (or not) in 10 signatures of ageing lung BEC subpopulations. **(C)** Violin plots of expression of pan-endothelial and LEC markers in 12 identified BEC subpopulations. **(D)** Dot plots of 49 genes identified as differentially expressed by the heatmap in B, plotted alongside pan-EC and LEC markers and control genes for comparison, and used in annotation of 12 identified BEC subpopulations (see also annotation summary presented in Figure 4). Note that 39 (marked in red) of 49 identified subpopulation-specific DEG have not been previously assigned as markers of individual generic subpopulations of ageing human lung BEC. **(E)** Representative lineage map in a form of a three-dimensional (3D) UMAP generated by performing trajectory analysis using the R package Slingshot. Plotted lines are trajectories representing predicted relationships between 12 identified BEC subpopulations. 3D video can be found Additional file 4: Video S1. **(F)** Table of module scores (Additional file 1: Detailed methods) grouped by references from which reference signatures were derived. Six studies from 2013-2021 include: (i) Gillich et al., 2019 (ii) Vanlandewijck et al., 2019; (iii) Iso et al., 2013; (iv) Nukala et al., 2021; (v) Ochiya et al., 2014; (vi) Herwig et al., 2016. Abbreviations: aCAP - aerocyte capillary; gCAP - general capillary; large – large vessel; art- artery; CAP1 - capillary 1; CAP2 - capillary 2; bronc – bronchial; S vein - systemic vein; P vein - pulmonary vein; Acap - aerocyte capillary; Gcap - general capillary; PeriB – Peribronchial (Note that names given by the authors are preserved). *Coral red* represents a subpopulation with positive score (above 0) for signature, colour intensity is proportional to the score positivity. Additional information is available in Additional file 1: Detailed methods, Annotation of BEC subpopulation. UMAP - Uniform Manifold Approximation and Projection.

Expression analysis of the 49 subpopulation-specific DEG (Figure 2D) in the total lung revealed that the majority of them were not EC-specific, i.e. also expressed in other cell types (Additional file 2: Figure S4A). 39 of them have not been previously assigned as markers of 8 generic subpopulations of BEC identified and characterised earlier in human or mouse lung [21, 22, 25, 26, 33], and thus are newly associated with ageing human lung endothelium (Figure 2D; Additional file 2: Figure S4A, B). A pathway analysis uncovered their belonging to specific classes of proteins, including secreted molecules, cell surface receptors, transcription factors and sub-cellular distribution (Additional file 2: Figure S4B-C).

Three-dimensional pseudo-time lineage analysis revealed close relationships between identified BEC subpopulations (Figure 2E; Additional file 4: Video S1). This included a direct link between subpopulations 5 and 3, and manifestation of subpopulation zero as a “convergence point” for seven subpopulations - 1, 2, 4, 6, 9, 10, 11 and, partially, 8. The expression analysis of markers of EC sub-types proposed in other transcriptional studies in human and murine lungs [21, 26, 33–38] was applied to 12 ageing human lung BEC subpopulations identified in our study, and revealed only few distinct “marker / ageing human lung BEC subpopulation” matches (Additional file 2: Figures S6, S7). Further exploration of identities of 12 subpopulations by using these markers and also those proposed for specific EC subpopulations in HLCA and IPF cell atlas studies [21, 22, 24–26], when coupled with module scoring (Figure 2F) and cell cycle (Additional file 2: Figure S8) analyses, confirmed these findings, whilst 4 subpopulations (zero, 5, 10 and 11) remained un-matched (Figures 2F). Finally, quantitative analysis revealed a significant increase in representation of two subpopulations (1 and 2) and overall decrease in subpopulation 7 in IPF (Figure 3A-C; Additional file 3: Tables S5, S6), with specificity by the cohort (Figure 3D).

**Figure 3.**
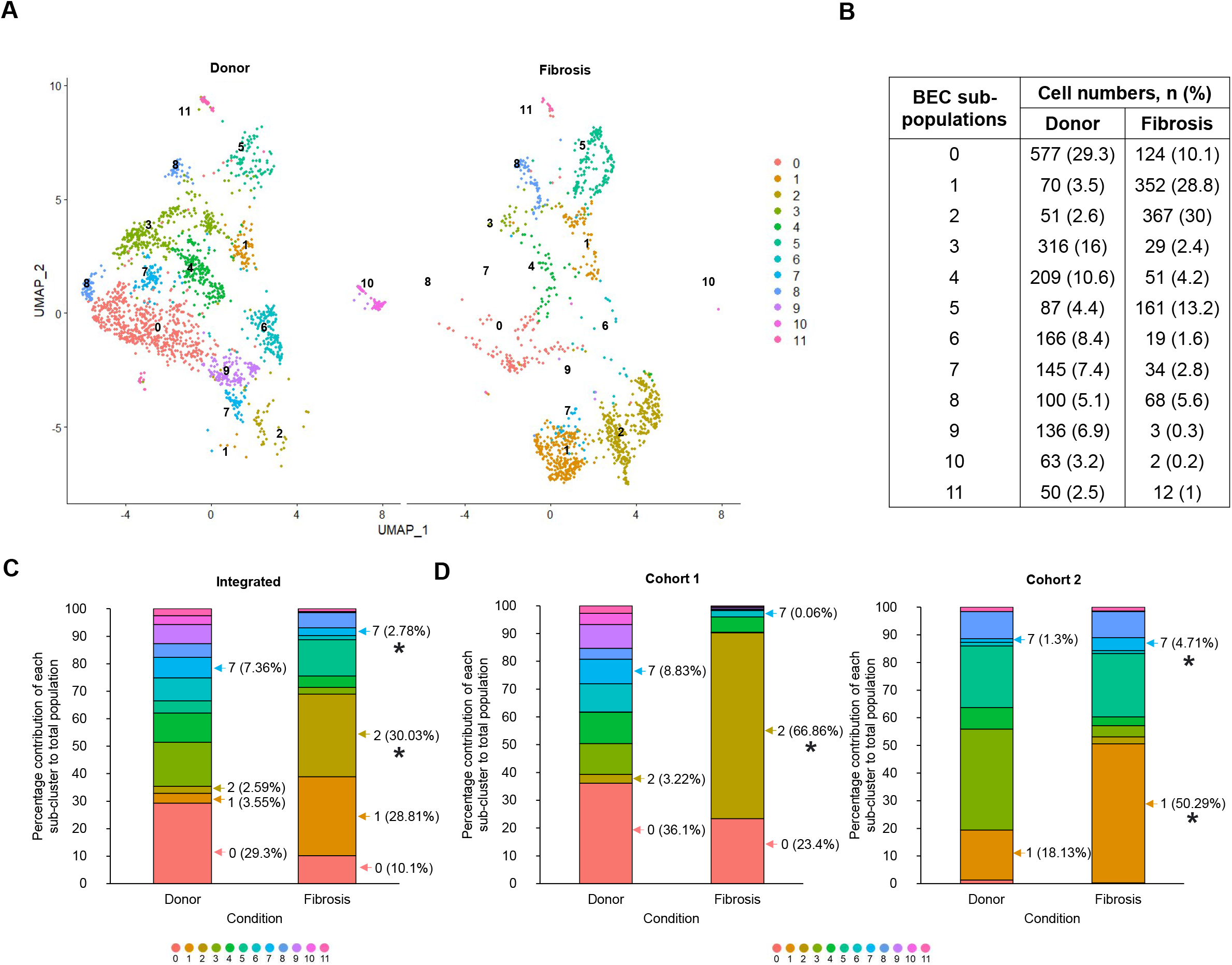
Percentage contributions of blood endothelial cell subpopulations in donors and IPF patients. Quantitative analysis of percentage contribution of cells from each subpopulation to total blood endothelial cell (BEC) population in ageing human lung from healthy donors and IPF patients was conducted using Seurat v4. **(A)** UMAP of BEC split by sample condition (donor or fibrosis). **(B)** Table detailing the number of cells per cluster, split by condition. **(C-D)** Stacked bar charts of percentage contribution of each subpopulation to total endothelial cell population split by condition for **(C)** total integrated dataset and **(D)** cohort 1 (*left)* and cohort 2 (*right)*. Cell numbers from each individual sample were analysed using Multiple t test. P < 0.05 (*) was considered significant. UMAP - Uniform Manifold Approximation and Projection.

### Annotation of Blood Vessel Endothelial Cell Subpopulations and Identification of Dedifferentiated Cells in Ageing Human Lung

Altogether, our findings formed the foundation for the annotation of 12 ageing human lung BEC subpopulations with distinct transcriptional signatures (Figure 4A; in-depth description is provided in Additional file 1: Detailed methods, Annotation of BEC subpopulations). Integrative analysis revealed that ageing human lung BEC belong to three and 8 subpopulations of bronchial and pulmonary circulation networks respectively (with one, inflammatory, being “intermediary” as also identified in other reports [24]), and include two previously undescribed subpopulations - designated as “de-differentiated” endothelium - in pulmonary circulation (Figures 4A, 2F, Additional file 2: Figure S9). Ten subpopulations of ageing human lung BEC have distinct transcriptional signatures, whilst no obvious DEG could be identified for subpopulations zero and 5 (Figures 2B, 2D, 4A, Additional file 2: Figures S5, S11; in-depth description is provided in Additional file 1: Detailed methods, Annotation of BEC subpopulations).

**Figure 4.**
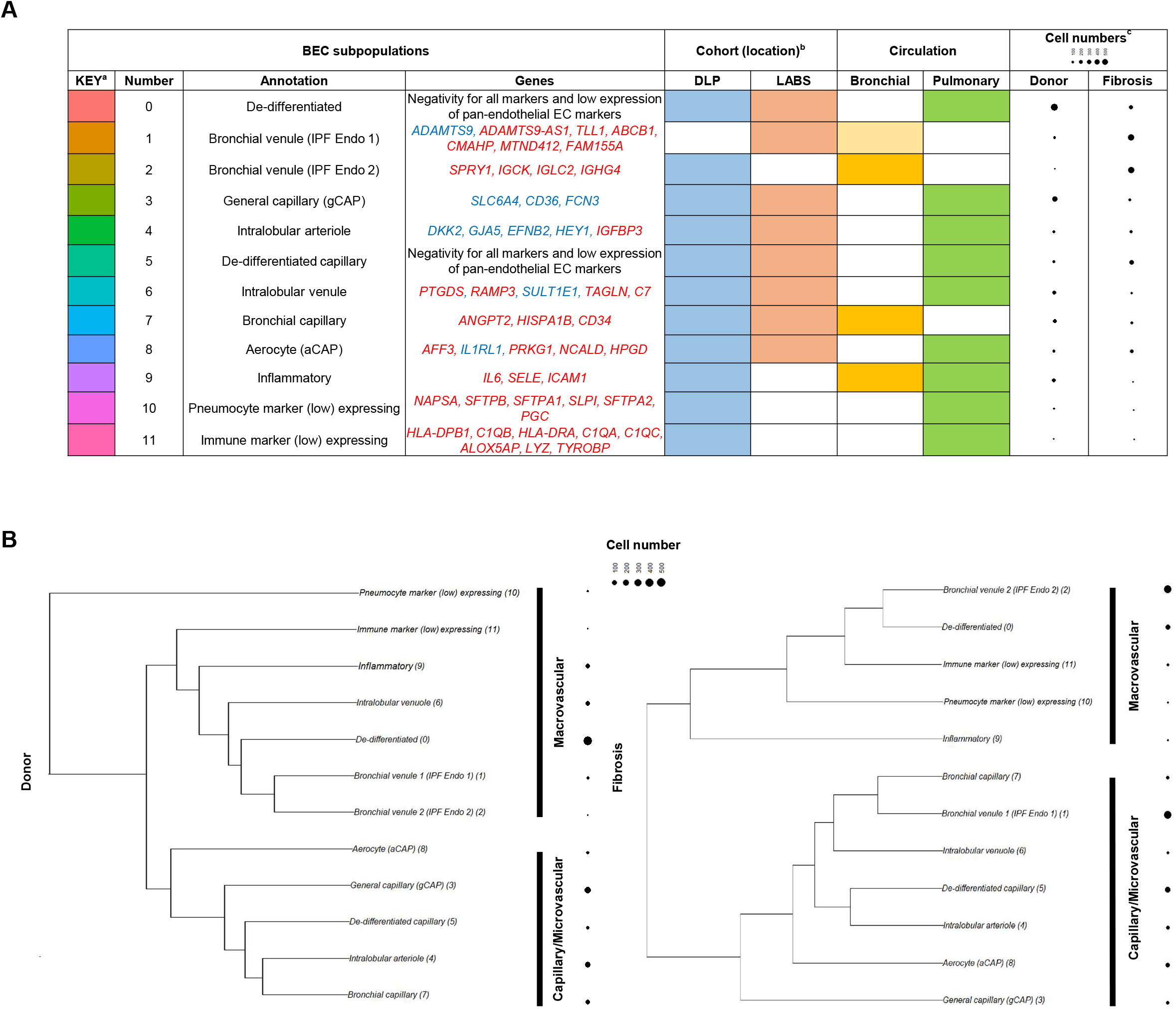
Annotation of subpopulations and relationship between them in donors and IPF patients. The profiling of markers of EC sub-types (Figure 2F, Additional file 2: Figures S6, S7, S9), when combined with the expression of pan-EC and LEC markers (Figure 2C-D) and sub-population-specific signatures identified in this study (based on expression pattern of 49 DEG; Figures 2D, Additional file 2: Figure S3), lung tissue sample collection locations (Table 1), modules scores of association with bronchial or pulmonary circulatory networks (Figure 2F, Additional file 2: Figure S9), EC differentiation status (Additional file 2: Figure S12), IPF-related alterations in cell numbers (Figure 3), high expression of *IL6*, *SELE* and *ICAM1* [21, 24]; Figure 2D), cell cycle analysis data (Additional file 2: Figure S8) and findings from three-dimensional pseudo-time lineage analysis (Figure 2E, Additional file 4: Video S1), formed the foundation for the annotation of 12 identified ageing human lung BEC subpopulations with distinct transcriptional signatures (Note that in-depth description is provided in Additional file 1: Detailed methods, Annotation of BEC subpopulations). **(A)** Table summarizing the information taken into account for naming/annotating subpopulations, including gene signatures, sample location by cohort, blood vessel circulatory networks (bronchial and pulmonary) and cell numbers (by condition - donor or IPF/fibrosis). ^a^ Key colour code for subpopulations is the same as in Figures 2 and 3. ^b^ Sample location (by cohort) information corresponds to Table 1. DLP = distal lung parenchyma, LABS = Longitudinal apical to basal segments. ^c^ Cell number dot plots corresponds to data in Figure 3. Signature genes include previously reported (blue; n=10) and *de novo* identified in our study genes (red; n=39). **(B)** Dendrogram (cluster tree) and dot plot of subpopulations, based on unsupervised hierarchical clustering of 12 subpopulations of ageing human lung BEC from integrated total dataset, which was split into two separate objects by condition (donor on the *left* and fibrosis on the *right*). Stratification into two groups termed “capillary” (or “microvascular”) and “macrovascular” is based on distinct clustering of capillary sub-clusters (3, 5, 7 and 8).

Subpopulations zero, 5, 10 and 11 had similar expression levels and ridgeplot profiles (histograms) to all other BEC subpopulations for the housekeeping gene beta-2 macroglobulin (B2M; Figure 2D; Additional file 2: Figure S12A). These subpopulations also exhibited low, but similar in ridgeplot profiles to other BEC subpopulations, expression of pan-EC genes (Figure 2C-D, Additional file 2: Figure S15A, B). Subpopulations zero and 5 contained a lower average number of genes per cell compared to other BEC subpopulations (Additional file 2: Figure S12B) and did not express markers of 23 non-EC cell clusters of human lung (Additional file 3: Table S1; Additional file 2: Figure S12C) or EC progenitors (Additional file 2: Figure S12D).

Next, we used these novel identified signatures (Figure 4A) to test the hypothesis that BEC subpopulation zero (the largest one in donor group in integrated dataset; Figure 3C) is exclusively associated with ageing in the human lung. The comparison of scRNAseq datasets of four ageing and three non-ageing/young human lungs against these signatures led to annotation of BEC subpopulations in both age groups (Additional file 2: Figures S13, S14). All signatures, except for subpopulations 1 and 2 (bronchial venules IPF Endo 1 and 2, which are noticeably associated mainly with ageing IPF lung in cohorts 2 and 1 respectively; Figure 3C-D), were matching specific subpopulations in both age groups (as expected, since these were donor lung samples, Additional file 2: Figure S14E). Cell number analysis revealed that BEC subpopulation zero (identified as subpopulation A in ageing vs non-ageing/young lung analysis), which has no obvious DEG, was present within ageing human lung only (Additional file 2: Figures S13B-C, S14E), supporting our hypothesis and its annotation as an ageing-related “de-differentiated” BEC (Figure 4A).

Finally, unsupervised hierarchical clustering of 12 subpopulations of ageing human lung BEC revealed two distinct groups - “capillary/microvascular” and “macrovascular” (Figure 4B). In IPF, the relationships between subpopulations were altered, with “bronchial venule/IPF Endo 1” and “intralobular venule” subpopulations “translocating” to “capillary/ microvascular” (Figure 4B).

### Cell Numbers for Bronchial Blood Vessel Endothelial Cell Subpopulations in Ageing Distal Lung Depend on Fibrosis Degree and Bronchi Number in IPF

To investigate the reason for differential cell numbers observed for two bronchial BEC subpopulations (2 and 7; bronchial venule and capillary BEC respectively) in IPF (Figure 3C, D), we focused on analysing scRNAseq data from the distal part of human ageing lungs (cohort 1; four donors and four IPF patients; Figure 5 A, B). This analysis exposed considerable variation in a percentage of subpopulation 2 cells between individual samples in IPF (5.48 - 91.6 %), suggesting inter-lung sample heterogeneity. The comparison of scRNAseq data with IPF lungs histology (based on haematoxylin and eosin staining) for each individual case, which has been made available by the authors in the supplementary materials section of the original study [27], revealed that the lowest percentage of subpopulation 2 cells was detected in sample IPF04, where there was a noticeable lack of bronchial structures. Based on these findings, we hypothesised that in IPF lung, significantly altered heterogeneity (in terms of differential cell numbers) of ageing bronchial (and also possibly pulmonary and lymphatic) endothelium reflects association with histological changes, i.e. tissue composition, and, in particular, with the increase in percentage of (i) areas occupied by peri-bronchial spaces undergoing fibrotic alterations and/or (ii) a number of bronchi, in particular, which both underpin inter- and intra-lung heterogeneity in IPF.

**Figure 5.**
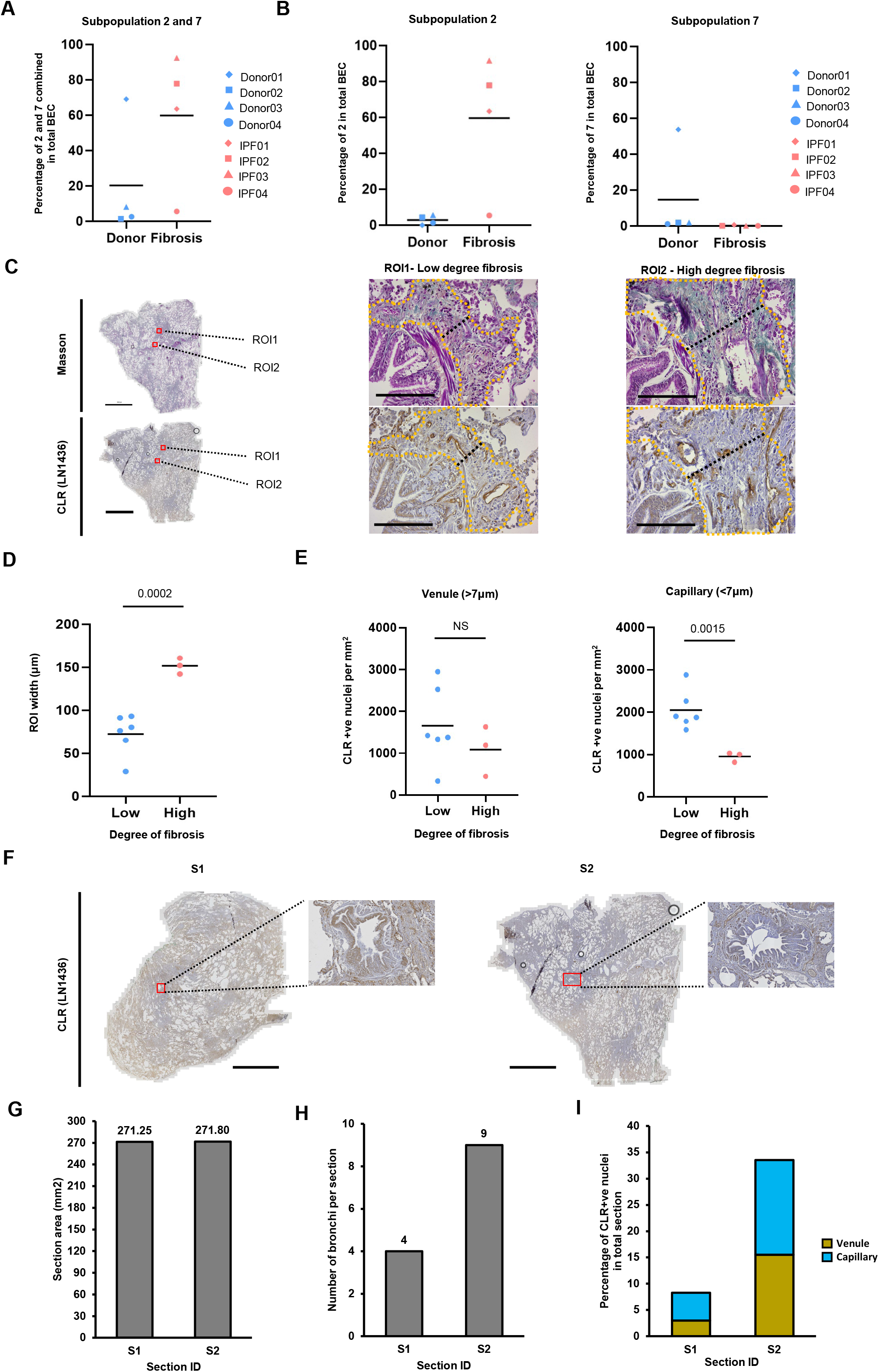
Spatial validation of bronchial blood endothelial subpopulations in distal lung by immunohistochemistry and quantitative high content image analysis. **(A-B)** Dotplots of quantitative analysis of percentage contribution of cells from bronchial subpopulation 2 and 7 to total blood endothelial cell (BEC) population in ageing human lung from healthy donors (n=4) and IPF patients (n=4) from cohort 1 (Table 1) was conducted using Seurat v4. Donor samples are shown in blue, fibrosis samples in coral red. **(C)** Masson’s trichrome and **(C, F)** immunohistochemical staining for calcitonin receptor like receptor (CLR) was conducted on formalin fixed paraffin embedded (FFPE) tissue sections of distal lung surgical biopsy from a 58-year-old male patient with IPF (with confirmed histopathological pattern of usual interstitial pneumonia) using our anti-human CLR antibody LN1436 [43]). **(C)** Images of scanned full sections (scalebars represent 5000µm) and two enlarged regions of interest (ROI) 1 and 2 (see full set in Additional file 2: Figure S16; scalebars represent 100µm). Yellow dotted line represents peri-bronchial areas (between bronchi and alveolar regions) that were analysed. Black dotted line represents example measurement of width of peri-bronchial area (see Additional file 1: Detailed methods). Dot plots of quantitative analysis of **(D)** peri- bronchial area width and **(E)** number of CLR-positive nuclei from 9 analysed ROI. **(D-E)** Low degree fibrosis areas are shown in blue, high degree fibrosis areas in red. A cut off of 90µm for average peri-bronchial area width was used to determine high (> 90) and lower (< 90) degree fibrosis. Shapiro Wilcoxon normality test and Mann-Whitney U test were used to analyse the data. P values are indicated at the top. Dotplots presenting: **(D)** quantification for peri-bronchial areas width in µm and **(E)** a number of CLR positive nuclei per mm^2^ (venule nuclei, *left*; capillary nuclei, *right*). **(F)** Two sections (S1 and S2) from the same distal lung biopsy were used for quantitative high content Image J analysis. Individual bronchi were identified within the total section under microscope and selected for downstream analysis (Additional file 1: Detailed methods). Scale bars represent 5000 µm. Indicated with red box and enlarged to the right of each scanned section are two representative examples of bronchi in individual sections (see full set in Additional file 2: Figure S17). Barcharts representing the results of quantitative image analysis of: **(G)** total section areas in mm^2^, **(H)** number of bronchi per section and **(I)** percentage of CLR-positive nuclei in total section; all done for S1 and S2 sections. Colours for bronchial venules and capillaries match colours of ageing human lung BEC subpopulations 2 and 7 presented throughout paper (see Figure 4A as an example).

To test this hypothesis, we analysed a distal lung surgical diagnostic lung biopsy from a 58-year-old male patient with IPF, confirmed by a histopathological pattern of UIP. First, we identified 8 BEC subpopulations, including 2 and 7, by spatial position/localisation in ageing human lung tissue of EC by immunostaining using two pan-endothelial (for various human tissues, including lung) markers - platelet endothelial cell adhesion molecule (*PECAM-1*), also known as cluster of differentiation 31 (CD31; [41]) and calcitonin receptor-like receptor (CLR, encoded by *CALCRL* gene; [42–44]; see also Figure 1D) (Additional file 2: Figure S15). Next, image analysis allowed quantification of IPF-associated changes in cell numbers for subpopulations 2 and 7 (Figure 5C-I). The comparative analysis of low vs high degree fibrotic areas within peri-bronchial regions in distal lung parenchyma identified no differences in the number of CLR-positive nuclei in venules (vessel diameter >7µm) and a significant decrease in capillaries (vessel diameter <7µm; Figure 5C-E; Additional file 2: Figure S16), reflecting BEC from subpopulations 2 and 7 respectively, between groups.

Furthermore, comparative analysis of sections from two different areas of a distal lung parenchyma from the same IPF patient revealed the association of percentage of CLR- positive nuclei for both capillaries and venules with the number of bronchi (Figure 5F-I; Additional file 2: Figure S17).

Altogether, these findings from immunohistochemistry corroborated the scRNAseq analysis of IPF-associated changes in cell numbers of bronchial BEC subpopulations 2 and 7 in distal lung parenchyma in four IPF patients from cohort 1. Importantly, it revealed the association of changes in cell numbers of ageing human BEC subpopulations 2 (venules) and 7 (capillaries) in peri-bronchial areas with the fibrosis degree and bronchi number.

### scRNAseq and Immunohistochemistry Reveal Fibrotic Alveolar Regions as Potential Niches for Dedifferentiated Blood Vessel Endothelial Cell Subpopulations in IPF

To investigate the possible location of BEC subpopulation zero in IPF, we focused on analysing scRNAseq data from the distal part of human ageing lungs (cohort 1; four donors and four IPF patients; Figure 6A). This exposed a noticeable variation in a percentage of cells between individual samples in IPF (3.79 - 73 %; Figure 6A), suggesting their inter-lung heterogeneity (similar to findings for BEC subpopulation 2; Figure 5B), and revealing a negative correlation between subpopulations zero and two (Figure 6B). Interestingly, the highest percentage of subpopulation zero cells was detected in the sample IPF04 (Figure 6C), which according to histology of individual IPF lungs (based on haematoxylin and eosin staining) for each individual case, which has been made available by the authors in the supplementary materials section of the original study ([27]), was associated with the lack of bronchial structures. Based on these findings and association of subpopulation zero with pulmonary circulation (4A; based on negative score for bronchial circulation signature, Figure 2F), we hypothesised that BEC subpopulation zero resides mainly in the alveolar regions or “niches” of the distal part of human ageing lungs in IPF, where it is likely to derive from BEC subpopulations 3, 4, 6 or 8 (Figures 2A, E, 3D and 4A).

**Figure 6.**
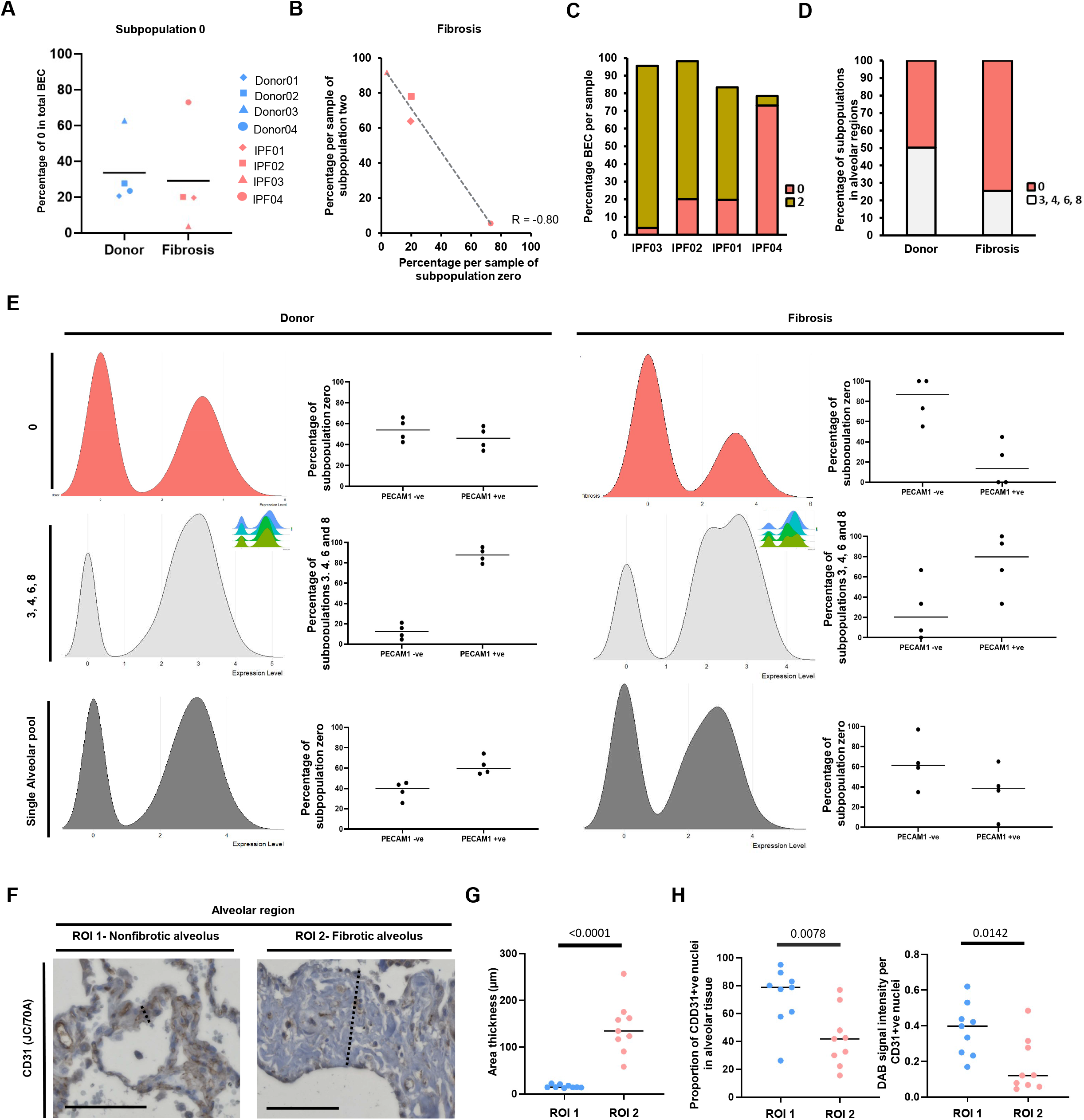
Comparative analysis of numbers of blood endothelial subpopulations in alveolar regions of distal lung in IPF by scRNAseq, immunohistochemistry and quantitative high content imaging. (A) Dotplot, **(B)** scatterplot and **(C-D)** stacked bar charts of quantitative analysis of percentage contribution of cells from subpopulations 0, 2, 3, 4, 6 and 8 to total blood endothelial cell (BEC) population in ageing human lung from healthy donors (n=4) and IPF patients (n=4) from cohort 1 (Table 1) was conducted using Seurat v4. **(A-B)** Donor samples are shown in blue, fibrosis samples - in coral red. **(B)** Pearson correlation analysis for percentage contribution of subpopulations zero vs subpopulation two in fibrosis samples. Line of best fit shown by grey dashed line. R - Pearson correlation coefficient. **(C)** Contribution of subpopulation zero and 2 to total BEC per sample coloured by subpopulation in fibrosis samples. Colours of ageing human lung BEC subpopulations match those presented throughout paper (see Figure 4A as an example). **(D)** Stacked bar chart showing percentage contribution of BEC subpopulation zero and combined alveolar BEC subpopulations 3, 4, 6 and 8, when analysed together as a “single alveolar pool”. Subpopulation zero is coloured by coral red, Subpopulations 3, 4, 6, 8 are coloured white. Data includes both donor and fibrosis samples. **(E)** Paired ridgeplots/histograms and dotplots of *PECAM1* expression in donor (left) and fibrosis (right) samples from cohort 1 (Table 1) was conducted using Seurat v4. *Top row,* (coral ridgeplot) - subpopulation zero *Middle row,* (light grey ridgeplot) - subpopulations 3, 4, 6 and 8 combined, with insert reflecting data for individual subpopulations; with colours of ageing human lung BEC subpopulations matching those presented throughout paper (see Figure 4A as an example). *Bottom row,* (grey ridgeplot) - subpopulations zero, 3, 4, 6 and 8 combined into a “single alveolar pool”. Note that the ridgeplot and dotplot for this pool in IPF patients (right) representing the *PECAM1* mRNA expression profile in alveolar region in the lung tissue, with increased number of CLR-negative BEC when compared to donors (left). **(F-H)** Immunohistochemical staining for CD31 was conducted on formalin fixed paraffin embedded (FFPE) tissue sections of distal lung surgical biopsy from a 58-year old male patient with IPF (with confirmed histopathological pattern of usual interstitial pneumonia) using mouse monoclonal antibody clone JC/70A. **(F)** Two representative regions of interest (ROI; 1 and 2) of alveolar regions of distal lung that were used for quantification are presented (see full set in Additional file 2: Figure S18). *Left,* low degree fibrosis, *right,* high degree fibrosis. Scalebar represents 100µm. Black dashed line represents example measurement of width of alveolar area analysed (for full details, see Additional file 1: Detailed methods). Dot plots of quantitative analysis of **(G)** alveolar area thickness in µm and **(H)** proportion of CD31 positive nuclei per mm2 (left) and average DAB signal intensity per CD31 positive nuclei (right). Nonfibrotic alveoli samples are shown in blue and fibrotic alveoli samples - in coral red. Nine ROIs were analysed for both nonfibrotic and fibrotic alveoli. Statistical analysis was done using Shapiro Wilcoxon test and Mann-Whitney U test. P values are indicated at the top. Note that immunohistochemistry and quantitative high content imaging profiles **(H)** confirm those from scRNAseq data analysis **(E).**

To test this hypothesis, scRNAseq data analysis was focused on distal ageing human lung samples (cohort 1), revealing that the increase in percentage of subpopulation zero cells in IPF lungs was associated with a decreased in percentage of alveolar subpopulations 3, 4, 6 or 8 BEC, when they are analysed together, as a “single alveolar pool” in ageing human lung (Figure 6D). Interestingly, subpopulation zero has expression profiles for housekeeping gene B2M (Figure 1B, Additional file 2: Figure S12A) and pan-EC markers (*PECAM1*/CD31 and *CALCLR*/CLR) (Additional file 2: Figures S15A, B) comparable to all other ageing human lung BEC subpopulations, whilst having a considerably larger proportion of cells with lower expression of the latter two genes. Furthermore, quantitative analysis of *PECAM1* expression in donor and fibrosis samples confirmed similarities in ridgeplot profiles between subpopulation zero and alveolar BEC subpopulations 3, 4, 6 and 8 (in terms of presence of both *PECAM1* positive and negative cells, as well as a trend for increased number of *PECAM1* negative cells in fibrosis; Figure 6E, top and middle panels). Integrative analysis of alveolar BEC subpopulations (3, 4, 6 and 8) and subpopulation zero taken together, as a “single alveolar pool” in ageing human lung, revealed the trend for a reduction of *PECAM1* expression (reflected by the decrease in percentage of positive cells) in fibrosis, also linked to an increased variability between individual samples (Figure 6E, bottom panel). Immunohistochemistry and image analysis of CD31 expression in distal alveoli in IPF lung revealed a statistically significant decrease in proportion of CD31-positive nuclei and CD31 signal intensity in fibrotic compared to non-fibrotic alveoli (Figure 6 F-H; Additional file 2: Figure S18). Combining analysis of *PECAM1/*CD31 expression by scRNAseq and immunohistochemistry revealed similar decreases in expression (compare last graphs in Figures 6E and H), thus supporting our hypothesis that BEC subpopulation zero resides mainly in the alveolar regions/niches of the distal part of human ageing lungs in IPF.

### Expression of Genes Linked to Disease-Specific Signalling Pathways and Endothelial Cell Processes is Altered in Ageing Lung Blood Vessel Endothelium in IPF

Comparison of gene expression between donor and IPF lungs led to identification of 3596 significantly regulated (1600 down- and 1996 up-) DEG across 12 BEC subpopulations (Figures 7A-B, Additional file 2: Figure S19A; Additional file 3: Table S7). There were significant associations with specific signalling pathways in 7 (two bronchial and 5 pulmonary, including two “de-differentiated”) from 12 subpopulations (Figure 7C). DEG in subpopulations zero, 8 and 11 were directly linked to IPF-associated pathways, and in four others (1, 3, 5 and 11) - with fibrotic or inflammatory pathways (Figures 7C, Additional file 2: Figure S19B). Next, the utilisation of published gene set enrichment analysis (GSEA) libraries for the assessment of expression of marker genes which are associated with ten EC- relevant processes revealed positive module scores in 5-12 of the BEC subpopulations for individual processes on average (Additional file 2: Figure S20) and significant changes in several (3 bronchial and 6 pulmonary, including two “de-differentiated”) BEC subpopulations in IPF (Figure 7D-E). This reflected subpopulation-specific pattern of endothelium response to the fibrotic lung environment in all processes, ranging from two (for senescence) to 8 (for vasodilation) subpopulations amongst 9 analysed (Figures 7E).

**Figure 7.**
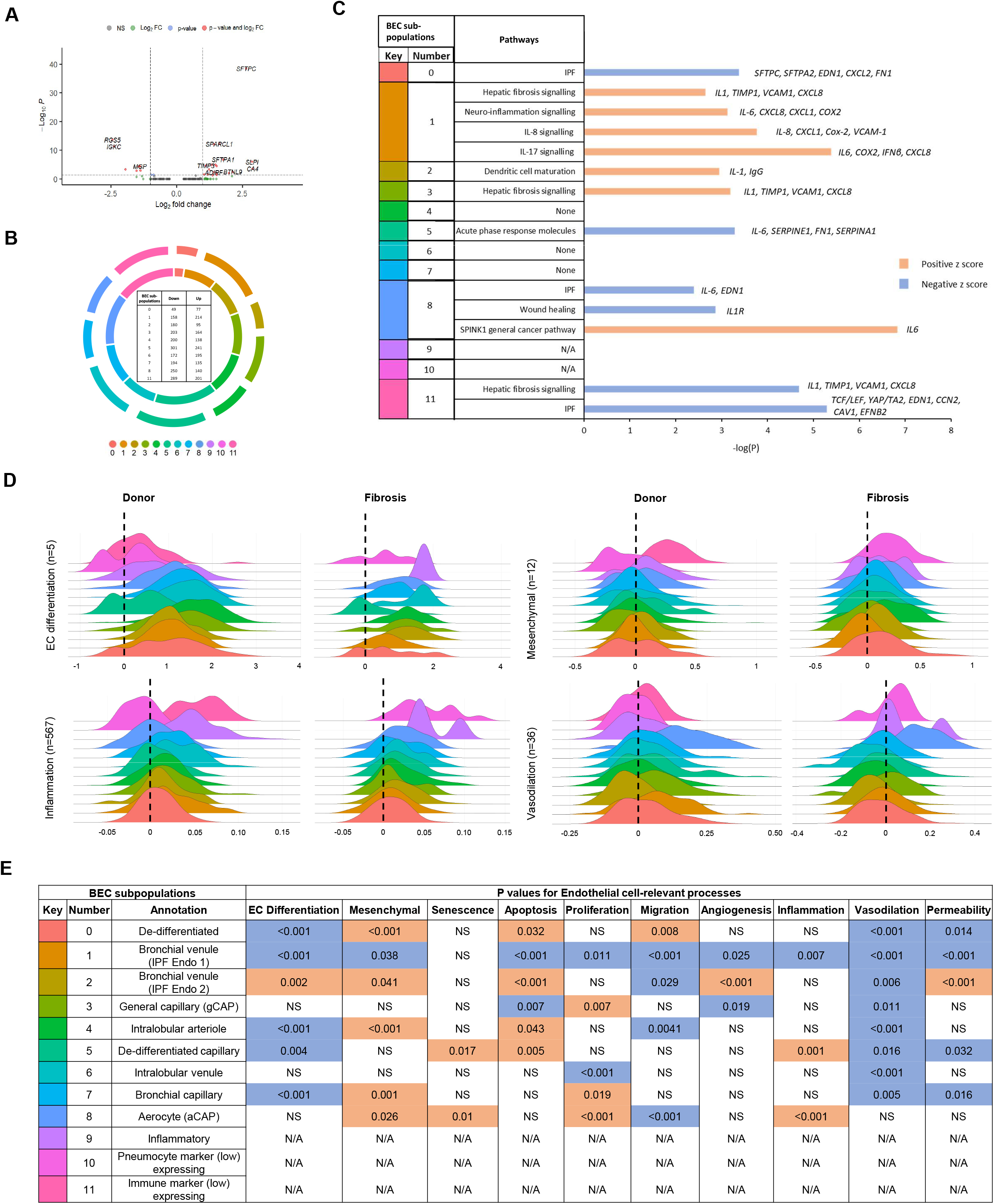
Association of differentially expressed genes in ageing human lung blood vessel endothelial cell with specific signalling pathways and endothelial cell-relevant processes in IPF. (A) An example of volcano plot of differentially expressed genes (DEG) (donor vs IPF/fibrosis) in blood endothelial cell (BEC) subpopulation zero. Volcano plots for other subpopulations can be found in the Additional file 2: Figure S19A. Statistical analysis was done using Shapiro Wilcoxon test and Wilcoxon rank sum test, with P value <0.05 (dotted line indicates cut-off) considered significant. **(B)** Doughnut plot of DEG between donor and IPF/fibrosis for each subpopulation. Outer ring shows up-regulated genes and inner ring represents down-regulated genes, with table in the centre showing numbers of DEG per subpopulation. Clusters 9 and 10 were excluded from the analysis, as they contained too few cells to analyse between conditions. **(C)** Bar chart detailing the results of the Ingenuity Pathway Analysis (IPA) using Core expression analysis function [77] of statistically significant altered pathways using DEG from B. *Blue colour* represents a negative Z score (down), and *orange* represents a positive Z score (up). DEG from three subpopulations (4, 6 and 7) did not significantly associate with any pathways and from two others (9 and 10) had too few cells in the IPF group for analysis. IPA was performed as described in Additional file 1: Detailed methods. **(D, E)** Key colour code for subpopulations is the same as in panel C and in Figures 2-4. **(D)** Histograms (ridgeplot) of endothelial cells differentiation (n= 5), mesenchymal transition (n= 12), inflammation (n= 567), and vasodilation (n= 36) scores in all blood endothelial subpopulations in donors (*donor*) and IPF (*fibrosis*). Ridgeplots for senescence (n= 79), apoptosis (n= 161), proliferation (n= 54), migration (n= 175), angiogenesis (n= 48), and permeability (n= 40) can be found in Additional file 2: Figure S20. Number of genes in sets are indicated in brackets. Black dotted line is added to annotate zero (cut-off point for differences). **(E)** Table detailing the P values for difference in module scores between donor and fibrosis. *Blue* colour represents down- regulation and *orange* - up-regulation in IPF. Statistical analysis was done using Shapiro Wilcoxon test and Mann-Whitney U test. UMAP - Uniform Manifold Approximation and Projection.

### Integrated Single Cell Map and Immunohistochemistry of Ageing Human Lung Lymphatic Endothelium Reveal Its Heterogeneity and Contribution in IPF

Unsupervised clustering of subtracted ageing human lung LEC alone in integrated dataset (689 cells) identified two main populations consisting of five distinct, and to our knowledge previously uncharacterised, subpopulations in total (Figures 1A, 8A), as based on their DEG patterns (top 10 DEG for each, 50 in total, with 20 not associated with LEC formerly [22, 26] (Figures 8B-D; Additional file 2: Figure S21, 22F) and cell cycle analysis (Additional file 2: Figure S23). All signatures were specific in recognising individual LEC subpopulations (Additional file 2: Figure S21A-B). Intriguingly, ageing human lung LEC subpopulations 1 and 2 were present only in cohort 1 and subpopulations zero, three and 4 – exclusively in cohort 2 (Additional file 2: Figure S24). Quantification of cell numbers revealed a trend for a decrease (for subpopulations 1 and 2) but overall a lack of non-significant changes for all subpopulations between donor and IPF lungs in integrated (Additional file 2: Figure S24A-B; Additional file 3: Table S9) or percentage contribution when split by cohort (Additional file 2: Figure 24C-E; Additional file 3: Table S9) datasets, in contrast to cell number data on BEC (Figure 3C, D). Altogether, our findings formed the foundation for detailed transcriptional characterisation of five distinct (informed by DEG, our transcriptional signatures, sample collection site and cell cycle information) LEC subpopulations in ageing human lung (Figure 8D; in-depth description is provided in Additional file 1: Detailed methods, Annotation of LEC Subpopulations).

**Figure 8.**
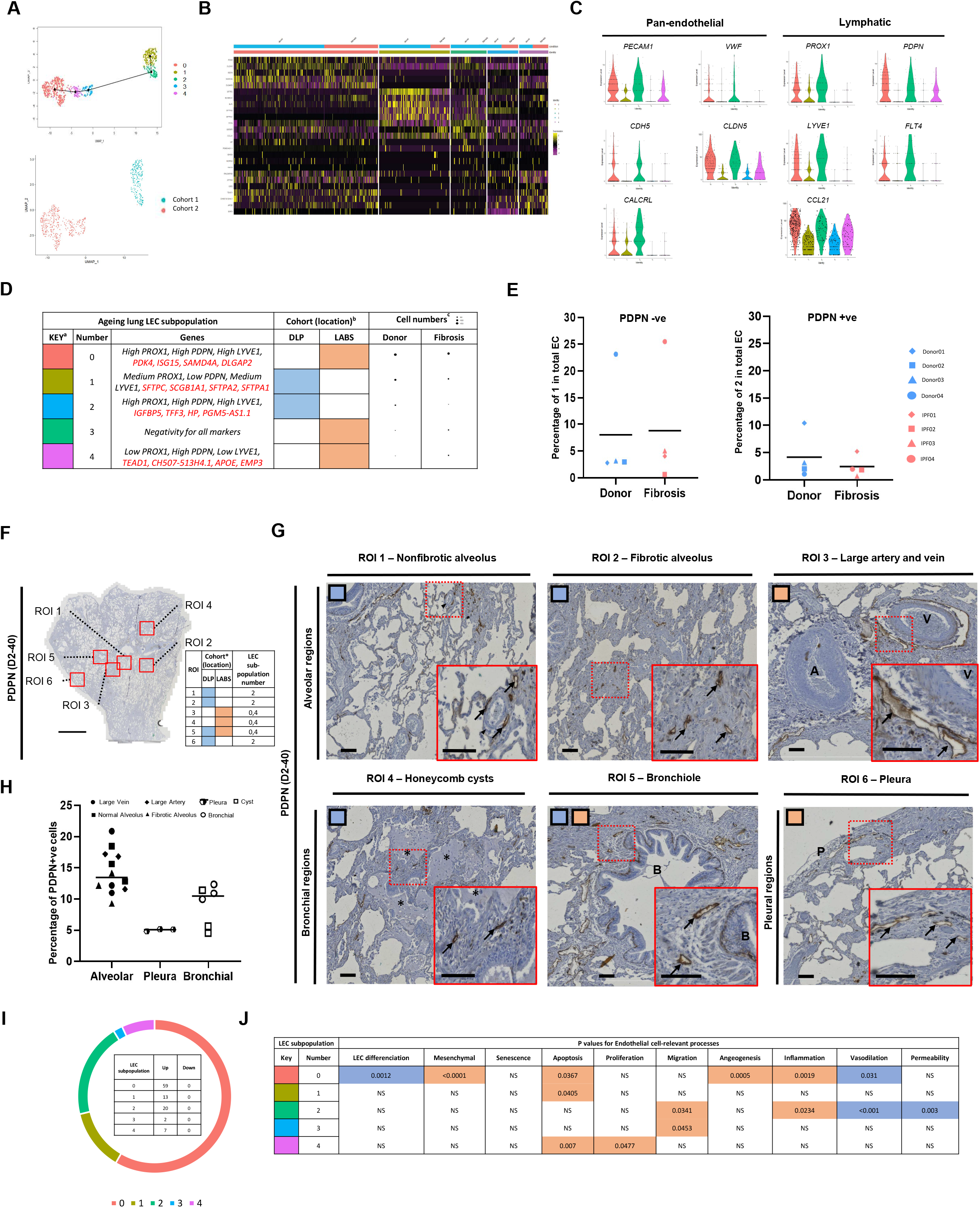
Cell subtype-specific composition of ageing human lung lymphatic endothelial cell cluster from donors and IPF patients from two independent cohorts. To comprehensively decipher transcriptional signatures of ageing human lung lymphatic vessel endothelium in IPF, we subtracted scRNAseq data for ageing human lung lymphatic vessel endothelial cell (LEC) (n=659) from the total lung cell populations from integrated dataset (two cohorts and both conditions, see Figure 1A), and sub-clustered them. (A) *Top,* UMAP representation of all cells from total LEC cluster/population (from all 18 pooled samples; Figure 1) with pseudotime lineage analysis of 5 resulting subclusters. *Bottom*, UMAP of LEC clusters coloured by cohort. Clusters were labelled 0-4 according to their signatures, which are presented as **(B)** heatmap of top 5 differentially expressed genes by cluster. Full details about each cell sub-population can be found in Additional file 2: Figure S21-23. **(C)** Violin plots demonstrating expression of pan-EC markers and LEC-specific genes (*PROX1*, *PDPN*, *LYVE-1* and *FLT4*) and *CCL21* [78] was done to confirm the identities of all cell subpopulations as LEC. **(D)** Table summarizing the information for each subpopulation, including gene signature (including previously reported, in *blue*, and *de novo* identified in our study, in *red*, genes), sample location by cohort and cell numbers. ^a^ Key colour code for subpopulations is the same as in Figure 8A. ^b^ Sample location (by cohort) information corresponds to Table 1. DLP = distal lung parenchyma, LABS = Longitudinal apical to basal segments. ^c^ Cell number dot plots correspond to data in Additional file 2: Figure S24 A, B. **(D)** Dotplot of quantification for percentage contribution of subpopulation 1 and 2 to total LEC per sample. *Left,* subpopulation 1, *right,* subpopulation 2. Donor samples are shown in blue, fibrosis samples - in coral red. **(F-H)** Immunohistochemical staining for PDPN was conducted on formalin fixed paraffin embedded (FFPE) tissue sections of distal lung surgical biopsy from a 58-year-old male patient with IPF (with confirmed histopathological pattern of usual interstitial pneumonia) using mouse monoclonal antibody clone D2-40. **(F)** Image of scanned full section (scalebars represent 5000µm), with six regions of interest (ROI) 1-6 labelled with red boxes and shown enlarged in (G) (scalebars represent 100µm). Table insert to the right is the summary of information for each ROI on whether observed structure could be identified in original publications for cohorts 1 and 2 (* - [25, 27]), i.e. DLP and LABS areas of the lung respectively (see D), and which LEC subpopulations were detected in our analysis (see A). **(G)** |Representative images for each ROI (magnified from F; scalebars represent 100µm), including inserts (red dotted boxes) of the areas containing PDPN-positive lymphatic vessels (*bottom right;* scalebars represent 100µm). Coloured squares in the top left corner correspond to mapping conducted using table presented in **(F)**. Arrows - lymphatic vessel; A - artery; V – vein; Arrowhead - small vessel; * - honeycomb cysts; B – bronchioles; P - pleura. (H) Dot plots of quantitative analysis of percentage of PDPN-positive nuclei from all identifiable nuclei per mm2 within individual ROI (n=3 per ROI), grouped by location within distal lung diagnostic biopsy of IPF (for full details, see Additional file 1: Detailed methods; images of 21 ROI used for high content image analysis are presented in Additional file 2: Figure S25). **(I)** Doughnut plot of DEG between donor and IPF/fibrosis for each subpopulation. Ring shows up-regulated genes, with table in the centre showing numbers of DEG per subpopulation (see also Additional file 3: Table S10). Note that no downregulated genes were found for LEC subpopulations. **(J)** Table detailing the P values for difference in module scores between donor and fibrosis. *Blue* colour represents down-regulation and *orange* – up-regulation in IPF. Statistical analysis was done using Shapiro Wilcoxon test and Mann-Whitney U test. UMAP - Uniform Manifold Approximation and Projection.

To investigate the reason for non-differential cell numbers observed for ageing human LEC in IPF (Additional file 2: Figure S24), compared to differential cell numbers observed for two bronchial BEC subpopulations (2 and 7), we focused scRNAseq data analysis on two LEC subpopulations (1 and 2) detected in the distal part of human ageing lungs from four donors and four IPF patients (cohort 1; Figure 8E). This exposed very small variation in a percentage of subpopulation 2 (PDPN-positive) cells between individual samples in IPF (0.71 - 5.20%; Figure 8E), contrasting findings for BEC subpopulations zero, two and 7 (Figures 5B and 6A, C). Based on this observation and on histology from 4 previous reports, jointly covering 5 different structures in ageing lung in donors or IPF patients [9, 11, 26, 46], we hypothesised that PDPN-positive LEC are distributed uniformly in all tissue regions (bronchial and alveolar) within ageing human distal lung parenchyma in IPF.

For testing this hypothesis, we immunostained human IPF lung tissue using anti-PDPN antibody D2-40 (Figure 8 F, G). The quantification analysis of immunostaining in this distal lung parenchyma sample uncovered signs of developing fibrosis and substantial intra-lung heterogeneity throughout the section areas/regions and revealed the distribution of PDPN-positive vessels (Additional file 2: Figure S25). Interestingly, the comparison to inter-lung heterogeneity, which was detected across four samples within cohort 1 (as demonstrated by histological analysis using haematoxylin and eosin staining; [27]) and cohort 2 (as demonstrated by immunohistochemistry images presented by the authors; [25]) detected the presence of all identifiable structures from both cohorts (non-fibrotic and fibrotic alveolae, bronchiole and honeycomb cyst in cohort 1; and large artery and veins, bronchiole and pleura in cohort 2) within the surgical distal lung biopsy. It also revealed uniform distribution of PDPN-positive LEC nuclei in all detected tissue regions (bronchial, alveolar and pleural) within ageing human distal lung parenchyma and honeycomb cyst regions (Figure 8 F-G; Additional file 2: Figure S25). Together with finding from scRNAseq data analysis, which showed that in ageing human lung PDPN-positive LEC subpopulation 2 was present only in cohort 1, whilst PDPN-positive LEC subpopulations zero and 4 - exclusively in cohort 2 (Figure 8C), these data suggest that our immunostaining detected all three subpopulations of PDPN-positive LEC subpopulations (zero, two and 4) in ageing human lung in IPF within our single diagnostic biopsy (Figure 8 F-G).

Next, high content image quantification of cell numbers/percentage of cells revealed uniform percentages of PDPN-positive LEC across analysed bronchial, alveolar and subpleural regions, including both fibrotic and non-fibrotic regions (ranging from 4.5% to 20.9%; Figure 8 H; Additional file 2; Figure S25). These findings support scRNAseq analysis data, which revealed lack of significant differences for these subpopulations in IPF lungs in the integrated dataset (Additional file 2: Figure S24A-B; Additional file 3: Table S9), thus supporting our hypothesis.

Finally, gene expression comparison between donor and IPF lungs led to identification of 101 significantly up-regulated DEG across 5 LEC subpopulations (Figures 8I), compared to 3596 DEG for ageing human ling BEC (Figure 7B) and significant linkage to IPF-associated pathways in subpopulations zero and one (Additional file 2: Figure S26). Finally, the utilisation of GSEA libraries identified significant alterations in ageing human lung LEC, in IPF (Figure 8J; Additional file 2: Figure S26 C-L).

## Discussion

The role of the lung circulatory system, including both blood and lymphatic vessel networks, has been implicated in IPF pathophysiology, but subpopulations of endothelium in ageing human lung and their disease-specific properties remained insufficiently characterized. Previous studies aimed at generating cellular atlases have predominantly used diverse datasets from healthy donors and patients with different diseases covering a wide range of ages, ethnicities and other characteristics [22, 25–27, 30, 32]. This complicated the deconvolution of a highly heterogeneous population of specialized cells in human lung and detection of changes triggered by the ageing process itself, as well as additional alterations caused by individual age-associated diseases. We generated a first integrated (multi-cohort) map of individual cell populations in ageing human lung in IPF and identified the contribution of blood and lymphatic vessels endothelium. This resource can be used as a foundation for identifying and studying ageing endothelium in other age-associated chronic lung diseases.

The identification and annotation of 17 subpopulations of endothelium (12 BEC and 5 LEC), with distinct molecular signatures and percentage contribution in ageing human lung in donors and IPF patients, is of particular significance. Integrated single-cell maps of the ageing human lung endothelium advanced resolution of BEC and LEC populations, increasing the number of formerly identified in ageing human lung EC subtypes [21, 22] from 9 to 17 and revealing previously underappreciated extend of their diversity and heterogeneity, especially in distal lung. Apart from confirming the presence of three bronchial and 8 pulmonary, as well as one “intermediary” (inflammatory), subpopulations, our integrative analysis detected four to our knowledge previously uncharacterised subpopulations of BEC in ageing human lung, when compared to previous scRNAseq reports [21, 22, 24]. Spatial validation based on immunohistochemistry using pan-EC markers confirmed the presence of 8 of these 12 BEC subpopulations in the distal ageing human lung. Our study also revealed the existence of 5 subpopulations of ageing LEC by transcriptional analysis. DEG signatures for 17 subpopulations of ageing human lung endothelium contain 59 novel subpopulation-specific candidate genes (39 for BEC, 20 for LEC). The pathway analysis of these DEG provides a springboard for exploring their roles and potential as biomarkers for diagnostics and/or prognostics in IPF.

Our study identified two BEC subpopulations (10 and 11, “Pneumocyte marker (low) expressing” and “Immune marker (low) expressing” respectively) in both the young and the ageing human lung. These subpopulations have been previously characterised in murine and porcine models [47–49], but to our knowledge have not been yet identified in the human lung. The pneumocyte marker (low) expressing subpopulation had been found in the context of pulmonary hypertension or obesity in mice [47, 48] and surfactant proteins have been detected in the circulation in IPF [50, 51]. The immune marker (low) expressing subpopulation had been found in porcine vasculature via generation of a multiple-organ single-cell transcriptomic map [49]. In human lung, these cells express markers of EC progenitors, including hematopoietic cells and hemogenic endothelium [52, 53]. These findings and abundance of both BEC subpopulations 10 and 11 in distal human lung warrant further studies to dissect their functions/roles in physiological conditions and chronic lung diseases.

To our knowledge, the identification of ageing human lung BEC subpopulations belonging to bronchial or pulmonary circulation or specific lymphatic networks [19, 20], including two subpopulations of “de-differentiated” cells (zero and 5), has not been conducted before our study. These “de-differentiated” BEC subpopulations were present in all nine analysed ageing human lungs from 49-72 years old subjects and belonged exclusively to pulmonary circulation. Findings from our scRNAseq analysis in IPF are in accordance with reports about the plasticity of endothelium and loss of EC identity, frequently associated with Endo- MT in vascular disorders [7, 54–56]. In our study, decreases in EC differentiation scores in three BEC subpopulations (zero, 4 and 7) and one LEC subpopulation (zero) in IPF co-occurred with increases in mesenchymal score. These findings suggest that Endo-MT, which has been previously proposed in IPF [56, 57], is also one of the subpopulation-specific features of the ageing human lung endothelium in this disease.

Lower average number of genes per cell associated with ageing human lung BEC de- differentiation in subpopulations zero and 5 has been also observed in few subpopulations of other cell types within ageing human lung in our study, concordant to findings for individual cell subpopulations identified in scRNAseq studies of ageing tissues from other organs [58, 59]. Importantly, subpopulations zero and 5 have comparable levels of the housekeeping gene *B2M* and pan-EC marker expression profiles (ridgeplots) to other ageing human lung BEC subpopulations, whilst having a considerably larger proportion of cells with lower expression of *PECAM1* and *CALCRL*. Together with distinct clustering of BEC population, including these two subpopulations, away from the rest of cell populations within the integrated single cell map of ageing human lungs (Figure 1B), these data support the identity of subpopulations zero and 5 as EC. The age-associated de-differentiation of human lung endothelium subpopulations, especially from alveolar regions (BEC sub-populations 3, 4, 6 and 8), is the most likely explanation for their “contribution” to subpopulation zero, which contained ∼30% of BEC in ageing human lungs. Together with pseudotime analysis data demonstrating that subcluster zero potentially act as a convergence “hub” for 7 subpopulations of ageing BEC, these findings could serve as an explanation for immunohistochemistry-based findings in our study and other reports using pan-EC markers and suggesting a loss of vasculature in IPF lung [10, 60].

Cell cycle analysis revealed that the majority (over 50%) of the cells in subpopulations zero, 4-7 and 9 in ageing human lung were in S phase. Cell cycle arrest and increase in S phase proportion have been linked to cell senescence in studies using *in vivo* or *in vitro* models, including murine lung EC [59, 61]. All nine analysed ageing human lung BEC subpopulations had positive senescence scores, which increased only in two pulmonary subpopulations, “de-differentiated capillary” and “aerocyte”, in IPF. Senescence has been classified as a hallmark of ageing [4], but remains challenging to analyse in human tissues and EC within any organ. To date, the presence of senescent endothelium has been associated with hypertension and increased cardiovascular disease risk, but not chronic lung diseases or lung circulatory system pathology, in elderly subjects [62–64]. In our study, we report the presence and location of senescent EC in ageing blood vessels within distal lung parenchyma diagnostic biopsy of IPF, warranting further investigation.

The age-associated decline in properties of highly specialized EC subpopulations in human lung (de-differentiation due to Endo MT, global reduction in gene expression, and senescence) is likely to contribute to alterations of function of individual vessel sub-types comprising respective blood and lymphatic systems. Further exploration is needed into dissecting whether further de-differentiation or senescence [65] in individual subpopulations of IPF endothelium contribute to disease pathophysiology and/or progression and may present further avenues for therapy using drugs targeting these pathways in elderly. Furthermore, although rare, IPF can occur in younger adult populations (11.3 per 100LJ000 individuals; [66]), with a significant paucity of studies investigating the mechanisms of lung fibrosis pathogenesis. The contribution of revealed in our study ageing human lung BEC- related processes, and “de-differentiation” in particular, to IPF pathophysiology and susceptibility in young population requires further investigation.

Disease-specific heterogeneity of ageing human lung endothelium in IPF patients involves changes in the percentage contribution and transcriptional profiles of individual EC subpopulations. In IPF, observed significant increase in cell numbers in two (bronchial venules IPF Endo 1 and 2) and decrease in one (bronchial capillary) BEC subpopulations is associated with fibrosis, bronchi number and tissue sample location (distal parenchyma or other regions within lung), but accompanied by the lack of significant changes in lymphatic endothelium. These findings for ageing BEC are in accordance with IPF-associated changes for “VE peribronchial” subpopulation, which was identified by conducting analysis of age-unmatched lung tissue samples [25]. This subpopulation shares a degree of similarity with two subpopulations of ageing human lung BEC identified in our study - IPF Endo 1 and 2, which are transcriptionally distinct and derived from two independent cohorts (Figures 2F, 3D), with only one being from the same scRNAseq dataset as “VE peribronchial” subpopulation. Observed in our study decrease in percentage contribution of total LEC (Figure 1G) or LEC sub-population 2 in cohort 1 (distal lung samples) in IPF was non-significant, but concurs with decline observed by others in lymphatic vessel density in distal lung parenchyma in IPF [9]. These findings warrant further investigation of the specific bronchial BEC and LEC subpopulations as potential cellular targets for therapeutic management in IPF.

Changes in transcriptional profiles of individual subpopulations of ageing human lung EC related to key EC-relevant processes suggest their subpopulation-specific functional responses within the fibrotic lung microenvironment and roles in IPF pathophysiology. Alterations in three pulmonary BEC and one LEC subpopulation in IPF lung included DEG which significantly mapped to IPF pathways. This finding suggests previously unrecognized direct involvement of “de-differentiated”, “aerocyte” and “immune marker (low) expressing” BEC (subpopulations zero, 8 and 11 respectively) and LEC subpopulation 1 from ageing human lung in IPF and their potential utility as targets for therapy. Moreover, the association of DEG in one bronchial and three pulmonary BEC subpopulations with fibrotic or inflammatory pathways implies their active roles in IPF progression. Finally, the translocation of IPF Endo 1 subpopulation from the macrovascular to the capillary/microvascular group in IPF suggests a disease-specific shift in its transcriptional phenotype to a capillary/microvascular-like endothelium, which co-occurs with changes to its vascular tone and permeability.

Previous studies of lung vasculature in IPF suggest a contribution of increased permeability [15, 16], aberrant angiogenesis [67] and endothelial dysfunction [17] in blood vessels to the fibrotic lung microenvironment and disease progression. Our data show that eight BEC and two LEC subpopulations exhibit decreases in vasodilation scores, with five of these subpopulations (including two bronchial and two pulmonary “de-differentiated” BEC and LEC subpopulation one) also showing reduction in permeability scores in IPF. Two common co-morbidities in IPF are cardiovascular disease and pulmonary hypertension [68, 69], suggesting that systemic as well as lung-specific endothelial dysfunction could be contributing to IPF. Secondary endpoints in a recent clinical trial of the prostacyclin analogue treprostinil via non-systemic delivery (inhalation) to treat IPF patients with pulmonary hypertension, included improved lung function and a decrease in fibrosis progression events compared to placebo [70, 71]. These data are intriguing in the context of our findings indicating that pulmonary “de-differentiated” endothelium (subpopulations zero and 5) and the BEC from the “singular alveolar pool” (3, 4, 6 and 8) in IPF could be primary cellular targets for inhalation-based therapies affecting expression of genes involved in regulation of EC identity and vascular tone. Avenues for targeting these and other BEC or LEC subpopulations, which show changes in vasodilation and permeability scores in IPF, for therapy should be therefore explored/investigated.

To our knowledge, a limited number of studies characterizing individual lymphatic system networks (and vessel sub-types or LEC composing them) in physiologically ageing or ageing-associated IPF lungs have been conducted to date with main focus on intralobular and sub-pleural lymphatics [9, 11, 20, 26, 46]. Our study revealed 5 distinct sub-populations (based on 20 novel DEG and including three PDPN-positive subpopulations) of LEC, all with statistically unchanged cell numbers and modest changes in gene expression in IPF (101 vs 3596 DEG, when compared to BEC). The study also uncovered uniform distribution of PDPN-positive vessels in all tissue regions (bronchial, alveolar and pleural) within ageing human distal lung parenchyma detected using a single diagnostic biopsy of IPF. These findings bridge and substantially expands knowledge available from previous studies [9, 11, 26, 39, 40, 45, 46] to include honeycomb cysts. At the same time, this warrants further annotation and spatial validation of these subpopulation to facilitate dissection of their roles in pathophysiology of IPF and other chronic lung diseases.

IPF-associated changes in transcriptional profiles of LEC suggest subpopulation-specific pattern of responses to the fibrotic lung environment (with only 101 DEG in LEC subpopulations compared to 3596 identified for BEC), affecting EC-relevant processes predominantly in LEC from sub-populations zero and two. Transcriptional changes in LEC from ageing lung are likely to account for the alterations in their properties and reported “disappearance” of subpleural and interlobular lymphatics in IPF by others [9]. Furthermore, the network of subpleural lymphatic capillaries drains lymph from the surface of the lung [20] and presents potential route for drug delivery to distal lung tissue affected by fibrotic changes in IPF [72]. Lymphatics have been considered an attractive option for targeted therapy in cancer and should be also exploited for the management of IPF and other chronic lung diseases [73]. Therefore, further investigations of the roles of ageing human LEC in the fibrotic lung environment is warranted.

Current therapy for IPF patients aims to slow disease progression and maintain lung function [74, 75]. Two medications, nintedanib and pirfenidone, have been approved for the treatment of IPF on the basis of reductions in the rate of physiologic decline, but the disease still progresses in treated patients and there remains no cure [76]. Promising clinical trials such as the recent trial of treprostinil [70, 71] continue to improve the outlook for future therapy. Understanding the properties of individual circulation networks, blood and lymphatic EC subpopulations and their altered functions within pre-fibrotic and fibrotic microenvironments in IPF lung presents possible avenues for management (halting or reversing progression) of this debilitating and life-threatening disease. IPF endothelium and its individual subpopulations present potential cellular targets for therapy repurposing appropriate anti-fibrotic, -inflammatory or -senescence drugs in IPF, including the possibilities for personalised medicine.

## Conclusions

In summary, our study discovered the phenomenon of the distinct “IPF endothelium” phenotype/state in ageing human lung, which contributes to UIP pattern in IPF. We propose that disease-specific heterogeneity of ageing human lung endothelium (from blood and lymphatic vessels) should be considered as a hallmark of IPF. In the future, the exploitation of the knowledge about EC heterogeneity, including properties and functions of individual EC subpopulations, in ageing human lung has the potential in IPF to protect lung function, improving patients’ quality of life and survival. Our molecular findings should help with building a conceptual framework for appreciating the endothelium in ageing human lung as a significant contributor and a probable target for biomarker development and/or therapy in IPF, other chronic lung diseases and diseases associated with fibrosis in other organs, including lymphoedema and cancer.

## Methods

To characterise ageing human lung endothelium in IPF and to account for its intra- and inter-lung heterogeneity (within and between individuals; [29]), we conducted the comparative analysis of lung tissues from elderly IPF patients and healthy donors (49-72 years old for both groups) by integrating scRNAseq datasets from two independent cohorts, each composed of age-matched healthy and IPF lung samples [25, 27]. We generated integrated single-cell maps of ageing human lung endothelium, identified EC subpopulations and characterised their transcriptional profiles, differences in percentage contribution and gene expression signatures, and also conducted spatial analysis of findings by performing immunohistochemistry and high content microscopy using distal lung diagnostic biopsy of IPF (with confirmed histopathological pattern of usual interstitial pneumonia). Full details of the methods are provided in Additional file 1: Detailed methods.

## Supporting information

Additional file 1: Detailed methods

## Data Availability

All data produced in the present study are available upon reasonable request to the authors

https://www.ncbi.nlm.nih.gov/geo/query/acc.cgi?acc=GSE122960

https://www.ncbi.nlm.nih.gov/geo/query/acc.cgi?acc=GSE136831

## Declarations

### Ethics approval and consent to participate

This study utilised publicly available data. Detailed information can be found in the original articles [25; 27]. Analysis of surplus human lung biopsy tissue by immunohistochemistry was approved by the Hull and East Riding Research Ethics Committee (reference 08/H1304/54).

### Consent for publication

Not applicable.

### Availability of data and materials

The datasets analysed during the current study are available in gene expression omnibus (GEO). Data from cohort 1 [27] is available at GSE122960_RAW: https://www.ncbi.nlm.nih.gov/geo/query/acc.cgi?acc=GSE122960 and in cohort 2 [25] - from GSE136831_RAW: https://www.ncbi.nlm.nih.gov/geo/query/acc.cgi?acc=GSE136831.

### Competing interests

The authors declare that they have no competing interests.

### Funding

Kate Garthwaite Pulmonary Fibrosis Research Fund; University of Hull Endothelial Cell Research Fund; University of Hull PhD Scholarships Fund for “Health Global Data Pipeline (Health*GDP) for Biomedical Research and Clinical Applications” cluster; The Anatomical Society.

### Author contributions

ECF, SPH and LLN contributed to study conception and design. ECF, AAM and LLN performed bioinformatic analysis. ECF and LLN performed immunohistochemistry staining and image analysis. ECF and LLN analysed and interpreted data. SPH and LLN supervised study execution. ECF and LLN drafted the manuscript. All authors reviewed the manuscript.

## Acknowledgements

We thank all transplant donors and IPF patients who participated in the several studies analysed in this manuscript and the authors who made scRNAseq datasets publicly available. Mr Collins and Viper HPC Team, Drs Peter O’Toole, Joanne Marrison (both from The University of York Biotechnology Facility Team), Professors Adrian Harris, Anthony Maraveyas, Tatsuo Shimosawa and John Greenman, Drs Sarah De Val, Stephen Henderson, Camille Ettelaie, Markus Queisser, Veronica Carroll, Vicky Green, Mr Matthew Morfitt, Mr Dimitrios Manolis, Mrs Shirin Hasan and Mrs Nicola Briggs for their support.

## Notes

### Competing Interest Statement

The authors have declared no competing interest.

### Funding Statement

The work is funded by Kate Garthwaite Pulmonary Fibrosis Research Fund, the University of Hull Endothelial Cell Research Fund, PhD Scholarships Fund for 'Health Global Data Pipeline (Health*GDP) for biomedical research and clinical applications' cluster and The Anatomical Society.

### Author Declarations

IPF samples and age-matched donor samples in cohort 1 were selected from the gene expression omnibus (GEO) GSE122960_RAW and in cohort 2 - from GSE136831_RAW

### Summary of Updates

1). Previously presented figures have been updated. More specifically, in Figure 2D, novel genes in the signatures have been coloured in red. Furthermore, in Figure 3C and D, subpopulations with statistically significant differences in cell numbers between donor and fibrosis groups have been marked with an asterisk. Also, in Figure 4, the signature specificity ridgeplot and table have been moved to the Supplementary Material. Finally, Figure 5 has been renumbered as Figure 7, due to introduction of three new figures (see below). 2). New figures have been introduced to validate the scRNAseq findings by using immunohistochemistry and IPF biopsy. This includes Figures 5, 6 and 8. More specifically, Figures 5 and 6 contain new data on scRNA seq data analysis, immunostaining and high content imaging quantification for CLR and CD31. Next, Figure 8 includes lymphatic endothelial cell data which has been relocated from the Supplementary Data and also new data on immunostaining and high content imaging quantification for podoplanin. 3). Supplemental files have been updated by providing additional graphs for 5 of the previous figures and including 5 new figures and one table. 4). The main text has been updated to reflect the above mentioned changes, improve clarity of expression, update section titles (e.g. Integrated Single-Cell RNA Sequencing Analysis Reveals Endothelial Cell Contribution to Ageing Human Lung Tissue Microenvironment in IPF has been updated to Integrated Single Cell Map of Ageing Human Lungs Displays Endothelial Cell Contribution in Disease). Also, two new sections have been added to the text to describe new data presented in Figures 5 and 6 (see above). Furthermore, additional details have been provided to the description of findings on lymphatic endothelium - to reflect new data and relevant relocations from the Supplementary Material (see above). Finally, new sub-sections have been included in Methods section to reflect new experimental procedures which were used for generating data presented in Figures 5, 6 and 8. 5). Author affiliations have been updated to reflect recent changes.

## References

1. Zaman T, Lee JS. Risk factors for the development of idiopathic pulmonary fibrosis: A review. Curr Pulmonol Rep 2018; 7: 118–125.

2. Martinez FJ, Collard HR, Pardo A, Raghu G, Richeldi L, Selman M, Swigris JJ, Taniguchi H, Wells AU. Idiopathic pulmonary fibrosis. Nat Rev Dis Primers 2017; 3: 17074.

3. Maher TM, Bendstrup E, Dron L, Langley J, Smith G, Khalid JM, Patel H, Kreuter M. Global incidence and prevalence of idiopathic pulmonary fibrosis. Respiratory research 2021; 22: 1–10.

4. López-Otín C, Blasco MA, Partridge L, Serrano M, Kroemer G. The hallmarks of ageing. Cell 2013; 153: 1194–1217.

5. Strongman H, Kausar I, Maher TM. Incidence, prevalence, and survival of patients with idiopathic pulmonary fibrosis in the UK. Advances in therapy 2018; 35: 724–736.

6. Wolters PJ, Collard HR, Jones KD. Pathogenesis of idiopathic pulmonary fibrosis. Annu Rev Pathol 2014; 9: 157–179.

7. Gaikwad AV, Lu W, Dey S, Bhattarai P, Chia C, Larby J, Haug G, Myers S, Jaffar J, Westall G, Singhera GK, Hackett TL, Markos J, Eapen MS, Sohal SS. Vascular remodelling in idiopathic pulmonary fibrosis patients and its detrimental effect on lung physiology: potential role of endothelial-to-mesenchymal transition. ERJ Open Res 2022; 8.

8. Aspelund A, Robciuc MR, Karaman S, Makinen T, Alitalo K. Lymphatic System in Cardiovascular Medicine. Circ Res 2016; 118: 515–530.

9. Ebina M, Shibata N, Ohta H, Hisata S, Tamada T, Ono M, Okaya K, Kondo T, Nukiwa T. The disappearance of subpleural and interlobular lymphatics in idiopathic pulmonary fibrosis. Lymphatic research and biology 2010; 8: 199–207.

10. Ebina M, Shimizukawa M, Shibata N, Kimura Y, Suzuki T, Endo M, Sasano H, Kondo T, Nukiwa T. Heterogeneous increase in CD34-positive alveolar capillaries in idiopathic pulmonary fibrosis. American journal of respiratory and critical care medicine 2004; 169: 1203–1208.

11. El-Chemaly S, Malide D, Zudaire E, Ikeda Y, Weinberg BA, Pacheco-Rodriguez G, Rosas IO, Aparicio M, Ren P, MacDonald SD, Wu HP, Nathan SD, Cuttitta F, McCoy JP, Gochuico BR, Moss J. Abnormal lymphangiogenesis in idiopathic pulmonary fibrosis with insights into cellular and molecular mechanisms. Proc Natl Acad Sci U S A 2009; 106: 3958–3963.

12. Gaje PN, Stoia-Djeska I, Cimpean AM, Ceausu RA, Tudorache V, Raica M. Lymphangiogenesis as a Prerequisite in the Pathogenesis of Lung Fibrosis. In vivo 2014; 28: 367–373.

13. Glasgow CG, El-Chemaly S, Moss J. Lymphatics in lymphangioleiomyomatosis and idiopathic pulmonary fibrosis. European Respiratory Review 2012; 21: 196–206.

14. Lara AR, Cosgrove GP, Janssen WJ, Huie TJ, Burnham EL, Heinz DE, Curran-Everett D, Sahin H, Schwarz MI, Cool CD. Increased lymphatic vessel length is associated with the fibroblast reticulum and disease severity in usual interstitial pneumonia and nonspecific interstitial pneumonia. Chest 2012; 142: 1569–1576.

15. Montesi SB, Rao R, Liang LL, Goulart HE, Sharma A, Digumarthy SR, Shea BS, Seethamraju RT, Caravan P, Tager AM. Gadofosveset-enhanced lung magnetic resonance imageing to detect ongoing vascular leak in pulmonary fibrosis. European Respiratory Journal 2018; 51.

16. Montesi SB, Zhou IY, Liang LL, Digumarthy SR, Mercaldo S, Mercaldo N, Seethamraju RT, Rosen BR, Caravan P. Dynamic contrast-enhanced magnetic resonance imageing of the lung reveals important pathobiology in idiopathic pulmonary fibrosis. ERJ open research 2021; 7; 00907–2020

17. Probst CK, Montesi SB, Medoff BD, Shea BS, Knipe RS. Vascular permeability in the fibrotic lung. European Respiratory Journal 2020; 56; 1900100

18. Sgalla G, Iovene B, Calvello M, Ori M, Varone F, Richeldi L. Idiopathic pulmonary fibrosis: pathogenesis and management. Respiratory research 2018; 19: 1–18.

19. Sharara RS, Hattab Y, Patel K, DiSilvio B, Singh AC, Malik K. Introduction to the anatomy and physiology of pulmonary circulation. Critical Care Nursing Quarterly 2017; 40: 181–190.

20. Stump B, Cui Y, Kidambi P, Lamattina AM, El-Chemaly S. Lymphatic Changes in Respiratory Diseases: More than Just Remodeling of the Lung? Am J Respir Cell Mol Biol 2017; 57: 272–279.

21. Sauler M, McDonough JE, Adams TS, Kothapalli N, Barnthaler T, Werder RB, Schupp JC, Nouws J, Robertson MJ, Coarfa C. Characterization of the COPD alveolar niche using single-cell RNA sequencing. Nature Communications 2022; 13: 1–17.

22. Travaglini KJ, Nabhan AN, Penland L, Sinha R, Gillich A, Sit RV, Chang S, Conley SD, Mori Y, Seita J, Berry GJ, Shrager JB, Metzger RJ, Kuo CS, Neff N, Weissman IL, Quake SR, Krasnow MA. A molecular cell atlas of the human lung from single-cell RNA sequencing. Nature 2020; 587: 619–625.

23. Stuart T, Satija R. Integrative single-cell analysis. Nat Rev Genet 2019; 20: 257–272.

24. Neumark N, Cosme Jr C, Rose K-A, Kaminski N. The idiopathic pulmonary fibrosis cell atlas. American Physiological Society Bethesda, MD; 2020. p. L887–L892.

25. Adams TS, Schupp JC, Poli S, Ayaub EA, Neumark N, Ahangari F, Chu SG, Raby BA, DeIuliis G, Januszyk M. Single-cell RNA-seq reveals ectopic and aberrant lung- resident cell populations in idiopathic pulmonary fibrosis. Science advances 2020; 6: eaba1983.

26. Schupp JC, Adams TS, Cosme Jr C, Raredon MSB, Yuan Y, Omote N, Poli S, Chioccioli M, Rose K-A, Manning EP. Integrated single-cell atlas of endothelial cells of the human lung. Circulation 2021; 144: 286–302.

27. Reyfman PA, Walter JM, Joshi N, Anekalla KR, McQuattie-Pimentel AC, Chiu S, Fernandez R, Akbarpour M, Chen C-I, Ren Z. Single-cell transcriptomic analysis of human lung provides insights into the pathobiology of pulmonary fibrosis. American journal of respiratory and critical care medicine 2019; 199: 1517–1536.

28. Morse C, Tabib T, Sembrat J, Buschur KL, Bittar HT, Valenzi E, Jiang Y, Kass DJ, Gibson K, Chen W. Proliferating SPP1/MERTK-expressing macrophages in idiopathic pulmonary fibrosis. European Respiratory Journal 2019; 54(2):1802441

29. He D, Wang D, Lu P, Yang N, Xue Z, Zhu X, Zhang P, Fan G. Single-cell RNA sequencing reveals heterogeneous tumor and immune cell populations in early-stage lung adenocarcinomas harboring EGFR mutations. Oncogene 2021; 40: 355–368.

30. Habermann AC, Gutierrez AJ, Bui LT, Yahn SL, Winters NI, Calvi CL, Peter L, Chung M-I, Taylor CJ, Jetter C. Single-cell RNA sequencing reveals profibrotic roles of distinct epithelial and mesenchymal lineages in pulmonary fibrosis. Science advances 2020; 6: eaba1972.

31. Luecken MD, Zaragosi LE, Madissoon E, Sikkema L, Firsova AB, De Domenico E, et al. The discovAIR project: a roadmap towards the Human Lung Cell Atlas. Eur Respir J. 2022;60(2). 2102057

32. Vieira Braga FA, Kar G, Berg M, Carpaij OA, Polanski K, Simon LM, Brouwer S, Gomes T, Hesse L, Jiang J, Fasouli ES, Efremova M, Vento-Tormo R, Talavera- López C, Jonker MR, Affleck K, Palit S, Strzelecka PM, Firth HV, Mahbubani KT, Cvejic A, Meyer KB, Saeb-Parsy K, Luinge M, Brandsma CA, Timens W, Angelidis I, Strunz M, Koppelman GH, van Oosterhout AJ, Schiller HB, Theis FJ, van den Berge M, Nawijn MC, Teichmann SA. A cellular census of human lungs identifies novel cell states in health and in asthma. Nat Med 2019; 25: 1153–1163.

33. Gillich A, Zhang F, Farmer CG, Travaglini KJ, Tan SY, Gu M, Zhou B, Feinstein JA, Krasnow MA, Metzger RJ. Capillary cell-type specialization in the alveolus. Nature 2020; 586: 785–789.

34. Herwig N, Belter B, Pietzsch J. Extracellular S100A4 affects endothelial cell integrity and stimulates transmigration of A375 melanoma cells. Biochemical and Biophysical Research Communications 2016; 477: 963–969.

35. Iso T, Kedes L, Hamamori Y. HES and HERP families: multiple effectors of the Notch signaling pathway. J Cell Physiol 2003; 194: 237–255.

36. Nukala SB, Tura-Ceide O, Aldini G, Smolders V, Blanco I, Peinado VI, Castellà M, Barberà JA, Altomare A, Baron G, Carini M, Cascante M, D’Amato A. Protein network analyses of pulmonary endothelial cells in chronic thromboembolic pulmonary hypertension. Sci Rep 2021; 11: 5583.

37. Ochiya T, Takenaga K, Endo H. Silencing of S100A4, a metastasis-associated protein, in endothelial cells inhibits tumor angiogenesis and growth. Angiogenesis 2014; 17: 17–26.

38. Vanlandewijck M, He L, Mäe MA, Andrae J, Ando K, Del Gaudio F, Nahar K, Lebouvier T, Laviña B, Gouveia L, Sun Y, Raschperger E, Räsänen M, Zarb Y, Mochizuki N, Keller A, Lendahl U, Betsholtz C. A molecular atlas of cell types and zonation in the brain vasculature. Nature 2018; 554: 475–480.

39. Sibler E, He Y, Ducoli L, Keller N, Fujimoto N, Dieterich LC, Detmar M. Single-cell transcriptional heterogeneity of lymphatic endothelial cells in normal and inflamed murine lymph nodes. Cells 2021; 10: 1371.

40. Takeda A, Hollmén M, Dermadi D, Pan J, Brulois KF, Kaukonen R, Lönnberg T, Boström P, Koskivuo I, Irjala H, Miyasaka M, Salmi M, Butcher EC, Jalkanen S. Single-Cell Survey of Human Lymphatics Unveils Marked Endothelial Cell Heterogeneity and Mechanisms of Homing for Neutrophils. Immunity 2019; 51: 561–572.e565.

41. Pusztaszeri MP, Seelentag W, Bosman FT. Immunohistochemical Expression of Endothelial Markers CD31, CD34, von Willebrand Factor, and Fli-1 in Normal Human Tissues. Journal of Histochemistry & Cytochemistry. 2006;54(4):385–395

42. Nikitenko LL, Smith DM, Bicknell R, Rees MC. Transcriptional regulation of the CRLR gene in human microvascular endothelial cells by hypoxia. FASEB J. 2003 Aug;17(11):1499–501.

43. Nikitenko LL, Blucher N, Fox SB, Bicknell R, Smith DM, Rees MC. Adrenomedullin and CGRP interact with endogenous calcitonin-receptor-like receptor in endothelial cells and induce its desensitisation by different mechanisms. J Cell Sci. 2006;119(Pt 5):910–22.

44. Nikitenko LL, Leek R, Henderson S, Pillay N, Turley H, Generali D, Gunningham S, Morrin HR, Pellagatti A, Rees MC, Harris AL, Fox SB. The G-protein-coupled receptor CLR is upregulated in an autocrine loop with adrenomedullin in clear cell renal cell carcinoma and associated with poor prognosis. Clin Cancer Res. 2013;19(20):5740–8.

45. Xiang M, Grosso RA, Takeda A, Pan J, Bekkhus T, Brulois K, Dermadi D, Nordling S, Vanlandewijck M, Jalkanen S, Ulvmar MH, Butcher EC. A Single-Cell Transcriptional Roadmap of the Mouse and Human Lymph Node Lymphatic Vasculature. Front Cardiovasc Med 2020; 7: 52.

46. Kambouchner M, Bernaudin J-F. Intralobular pulmonary lymphatic distribution in normal human lung using D2-40 antipodoplanin immunostaining. Journal of Histochemistry & Cytochemistry 2009; 57: 643–648.

47. Bondareva O, Rodríguez-Aguilera JR, Oliveira F, Liao L, Rose A, Gupta A, Singh K, Geier F, Schuster J, Boeckel JN, Buescher JM, Kohli S, Klöting N, Isermann B, Blüher M, Sheikh BN. Single-cell profiling of vascular endothelial cells reveals progressive organ-specific vulnerabilities during obesity. Nat Metab. 2022 Nov;4(11):1591–1610

48. Rodor J, Chen SH, Scanlon JP, Monteiro JP, Caudrillier A, Sweta S, Stewart KR, Shmakova A, Dobie R, Henderson BEP, Stewart K, Hadoke PWF, Southwood M, Moore SD, Upton PD, Morrell NW, Li Z, Chan SY, Handen A, Lafyatis R, de Rooij LPMH, Henderson NC, Carmeliet P, Spiroski AM, Brittan M, Baker AH. Single-cell RNA sequencing profiling of mouse endothelial cells in response to pulmonary arterial hypertension. Cardiovasc Res. 2022 Aug 24;118(11):2519–2534.

49. Wang F, Ding P, Liang X, Ding X, Brandt CB, Sjöstedt E, Zhu J, Bolund S, Zhang L, de Rooij LPMH, Luo L, Wei Y, Zhao W, Lv Z, Haskó J, Li R, Qin Q, Jia Y, Wu W, Yuan Y, Pu M, Wang H, Wu A, Xie L, Liu P, Chen F, Herold J, Kalucka J, Karlsson M, Zhang X, Helmig RB, Fagerberg L, Lindskog C, Pontén F, Uhlen M, Bolund L, Jessen N, Jiang H, Xu X, Yang H, Carmeliet P, Mulder J, Chen D, Lin L, Luo Y. Endothelial cell heterogeneity and microglia regulons revealed by a pig cell landscape at single-cell level. Nat Commun. 2022 Jun 24;13(1):3620.

50. Bhatti F, Ball G, Hobbs R, Linens A, Munzar S, Akram R, Barber AJ, Anderson M, Elliott M, Edwards M. Pulmonary surfactant protein a is expressed in mouse retina by Müller cells and impacts neovascularization in oxygen-induced retinopathy. Invest Ophthalmol Vis Sci 2014; 56: 232–242.

51. Colmorten KB, Nexoe AB, Sorensen GL. The Dual Role of Surfactant Protein-D in Vascular Inflammation and Development of Cardiovascular Disease. Front Immunol 2019; 10: 2264.

52. Timmermans F, Plum J, Yöder MC, Ingram DA, Vandekerckhove B, Case J. Endothelial progenitor cells: identity defined? J Cell Mol Med. 2009 Jan;13(1):87–102.

53. Abdelgawad ME, Desterke C, Uzan G, Naserian S. Single-cell transcriptomic profiling and characterization of endothelial progenitor cells: new approach for finding novel markers. Stem Cell Res Ther. 2021 Feb 24;12(1):145.

54. Dejana E, Hirschi KK, Simons M. The molecular basis of endothelial cell plasticity. Nat Commun 2017; 8: 14361.

55. Gaikwad AV, Eapen MS, McAlinden KD, Chia C, Larby J, Myers S, Dey S, Haug G, Markos J, Glanville AR. Endothelial to mesenchymal transition (EndMT) and vascular remodeling in pulmonary hypertension and idiopathic pulmonary fibrosis. Expert Review of Respiratory Medicine 2020; 14: 1027–1043.

56. Piera-Velazquez S, Mendoza FA, Jimenez SA. Endothelial to mesenchymal transition (EndoMT) in the pathogenesis of human fibrotic diseases. Journal of clinical medicine 2016; 5: 45.

57. Jia W, Wang Z, Gao C, Wu J, Wu Q. Trajectory modeling of endothelial-to-mesenchymal transition reveals galectin-3 as a mediator in pulmonary fibrosis. Cell Death Dis. 2021 Mar 26;12(4):327

58. Durante MA, Kurtenbach S, Sargi ZB, Harbour JW, Choi R, Kurtenbach S, Goss GM, Matsunami H, Goldstein BJ. Single-cell analysis of olfactory neurogenesis and differentiation in adult humans. Nat Neurosci 2020; 23: 323–326.

59. Kumari R, Jat P. Mechanisms of cellular senescence: cell cycle arrest and senescence associated secretory phenotype. Frontiers in cell and developmental biology 2021; 9: 485.

60. Barratt S, Millar A. Vascular remodelling in the pathogenesis of idiopathic pulmonary fibrosis. QJM: An International Journal of Medicine 2014; 107: 515–519.

61. van Deursen JM. The role of senescent cells in ageing. Nature 2014; 509: 439–446.

62. Minamino T, Miyauchi H, Yoshida T, Ishida Y, Yoshida H, Komuro I. Endothelial cell senescence in human atherosclerosis: role of telomere in endothelial dysfunction. Circulation 2002; 105: 1541–1544.

63. Seals DR, Jablonski KL, Donato AJ. Ageing and vascular endothelial function in humans. Clin Sci (Lond) 2011; 120: 357–375.

64. Versari D, Daghini E, Virdis A, Ghiadoni L, Taddei S. The ageing endothelium, cardiovascular risk and disease in man. Exp Physiol 2009; 94: 317–321.

65. Ting KK, Coleman P, Zhao Y, Vadas MA, Gamble JR. The ageing endothelium. Vasc Biol 2021; 3: R35–R47.

66. Raghu G, Chen SY, Hou Q, Yeh WS, Collard HR. Incidence and prevalence of idiopathic pulmonary fibrosis in US adults 18-64 years old. Eur Respir J. 2016;48(1):179–86.

67. Barratt SL, Creamer A, Hayton C, Chaudhuri N. Idiopathic pulmonary fibrosis (IPF): an overview. Journal of clinical medicine 2018; 7: 201.

68. Farkas L, Gauldie J, Voelkel NF, Kolb M. Pulmonary hypertension and idiopathic pulmonary fibrosis: a tale of angiogenesis, apoptosis, and growth factors. Am J Respir Cell Mol Biol 2011; 45: 1–15.

69. Hubbard RB, Smith C, Le Jeune I, Gribbin J, Fogarty AW. The association between idiopathic pulmonary fibrosis and vascular disease: a population-based study. Am J Respir Crit Care Med 2008; 178: 1257–1261.

70. Nathan SD, Tapson VF, Elwing J, Rischard F, Mehta J, Shapiro S, Shen E, Deng C, Smith P, Waxman A. Efficacy of Inhaled Treprostinil on Multiple Disease Progression Events in Patients with Pulmonary Hypertension due to Parenchymal Lung Disease in the INCREASE Trial. Am J Respir Crit Care Med 2022; 205: 198–207.

71. Nathan SD, Waxman A, Rajagopal S, Case A, Johri S, DuBrock H, De La Zerda DJ, Sahay S, King C, Melendres-Groves L, Smith P, Shen E, Edwards LD, Nelsen A, Tapson VF. Inhaled treprostinil and forced vital capacity in patients with interstitial lung disease and associated pulmonary hypertension: a post-hoc analysis of the INCREASE study. Lancet Respir Med 2021; 9: 1266–1274.

72. Solari E, Marcozzi C, Ottaviani C, Negrini D, Moriondo A. Draining the Pleural Space: Lymphatic Vessels Facing the Most Challenging Task. Biology (Basel) 2022; 11. 419

73. Zhang XY, Lu WY. Recent advances in lymphatic targeted drug delivery system for tumor metastasis. Cancer Biol Med 2014; 11: 247–254.

74. Maher TM, Strek ME. Antifibrotic therapy for idiopathic pulmonary fibrosis: time to treat. Respir Res 2019; 20: 205.

75. Raghu G, Rochwerg B, Zhang Y, Garcia CA, Azuma A, Behr J, Brozek JL, Collard HR, Cunningham W, Homma S, Johkoh T, Martinez FJ, Myers J, Protzko SL, Richeldi L, Rind D, Selman M, Theodore A, Wells AU, Hoogsteden H, Schünemann HJ. An Official ATS/ERS/JRS/ALAT Clinical Practice Guideline: Treatment of Idiopathic Pulmonary Fibrosis. An Update of the 2011 Clinical Practice Guideline. Am J Respir Crit Care Med 2015; 192: e3-19.

76. Brownell R, Kaminski N, Woodruff PG, Bradford WZ, Richeldi L, Martinez FJ, Collard HR. Precision Medicine: The New Frontier in Idiopathic Pulmonary Fibrosis. Am J Respir Crit Care Med 2016; 193: 1213–1218.

77. Krämer A, Green J, Pollard Jr J, Tugendreich S. Causal analysis approaches in ingenuity pathway analysis. Bioinformatics 2014; 30: 523–530.

78. González-Loyola A, Petrova TV. Development and aging of the lymphatic vascular system. Adv Drug Deliv Rev. 2021;169: 63–78.

