## Additional file 1: Detailed methods for "Altered Heterogeneity of Ageing Lung Endothelium is a Hallmark of Idiopathic Pulmonary Fibrosis"

**Hallmark of Idiopathic Pulmonary Fibrosis**

Additional file 1: Detailed methods

**Detailed Methods**

**Dataset Selection**

For integrative analysis, single cell RNA sequencing (scRNAseq) data for lung tissue samples from ageing people were selected only from age-matched (49-72 years old) transplant donors and recipient IPF patients, and only from those studies that contained samples for both conditions. From all publicly available scRNAseq datasets of human IPF lungs (4 in total), only two (to our knowledge), fulfilled our selection criteria and were used for generating a single dataset consisting of cohorts for integrative analysis [1, 2]. IPF samples and age-matched donor samples in cohort 1 were selected from the gene expression omnibus (GEO) GSE122960_RAW [2] and in cohort 2 - from GSE136831_RAW [1]. Four age-matched pairs (scRNAseq data from lung tissue samples from transplantation stage IPF patients and healthy donors) from the first database [2] matched our selection criteria and formed cohort 1 (Table 1, Figure 1A). Five age-matched pairs from the second database (32 IPF and 28 donor samples ranging from 21 to 80 years old, [1], matching our selection criteria and forming cohort 2, were randomly selected for the integrated analysis in order to correspond to the number of cells analysed from cohort 1 (Table 1, Figure 1A). The number of cells analysed per cohort was matched as close as possible. More samples were selected in cohort 2 as they contained less cells per sample than cohort 1. In these two cohorts, the overall protocol for cell extraction and processing for scRNAseq was similar, but with three differences. Firstly, the samples were collected from peripheral distal lung tissue (parenchyma; 1-2cm) in cohort 1 and from longitudinal sections from apical to basal segments of the lungs in cohort 2. Secondly, cohort 2 samples were processed to derive single cells and then frozen in liquid nitrogen until processed in batches. Samples in cohort 1 were processed as soon as single cells were obtained from tissues. Thirdly, there were variations in reagents for the sample collection processes. The summary of the information on cohort-specific patient characteristics and lung tissue collection sites is outlined in Table 1. The collection from different regions across the lung presented the ideal platform to account for heterogeneity of tissue composition within and between the individual lungs included in our integrated analysis.

For the analysis of young vs old lung, three samples of young lung (aged 21-29; GSE122960_RAW; [2]) and the healthy elderly donor samples from cohorts 1 and 2 (n=9, aged 49-66, Table 1; GSE122960_RAW, [2]; GSE136831_RAW, [1]) were used. The individual samples were given the value ‘age’ and were labelled as ‘young’ and ‘old’ respectively.

**Ethics approval and consent to participate.**

This study utilised publicly available data. Detailed information can be found in the original articles [25; 27]. In brief, the Ethical approval and consent to participate is summarised below.

In cohort 1 [27], all procedures using human tissues were approved by the Northwestern University Institutional Review Board (STU00056197, STU00201137, and STU00202458) and by the University of Chicago Institutional Review Board (NCT00515567 and NCT00470327) and received further approval from The United States Army Medical Research and Materiel Command`s Office of Research Protections, Human Research Protection Office. All patients provided written informed consent.

In cohort 2 [25], IPF lungs were obtained from patients undergoing transplant while healthy lungs were from rejected donor lung organs that underwent lung transplantation at the Brigham and Women’s Hospital or donor organs provided by the National Disease Research Interchange (NDRI) approved by the Partners Healthcare Institutional Board Review (IRB Protocol # 2011P002419).

Analysis of surplus human lung biopsy tissue by immunohistochemistry (see section Immunohistochemistry below) was approved by the Hull and East Riding Research Ethics Committee (reference 08/H1304/54).

**Individual Sample Processing and Quality Control**

Seurat version 4 (v4) was used to perform dimensionality reduction, clustering, and visualization for the scRNAseq data [3]. Each individual selected sample was loaded from GEO, and the percentage of mitochondrial genes were calculated for each cell. Cells containing >5000, <200 identified genes or >20% of mitochondrial genes were removed. SCTransform with default parameters was used to normalize and scale the data, and dimensionality reduction was performed using PCA on the top 2000 most variable genes. Each individual sample was processed, and endothelial cells were identified before being saved as an R object before merging all samples into a total Seurat object, performing integration and clustering, as indicated below.

**Dataset Integration**

Unique molecular identifiers (UMI) for the remaining after filtering cell barcodes were analysed using the Seurat v4 considered one of the best integration tools [4]. The UMI counts were normalized using a scale factor of 10,000 UMIs per cell and then natural log transformed using a pseudo count of 1. Integration, embedding and clustering were performed as recommended by Seurat v4 [5]. Briefly, the top variable genes within each dataset were selected using the FindVariableFeatures function using the “vst” parameter. Shared patterns of variance in these genes within each dataset where then used to integrate the datasets using Seurat’s FindIntegrationAnchors and IntegrateData, then the resulting integrated expression matrix was scaled using Seurat’s ScaleData function.

**Dimension Reduction, Graph Embedding, Clustering and Visualization**

Scaled values from the integrated assay were subject to principal component analysis (PCA). Principal components (PCs) were then subject to feature selection based on ranked contribution to variance. Selected PCs were used to estimate Euclidean distances between cells in feature-space, and cells are graph embedded with edges connecting the nearest neighbours for each cell using the FindNeighbors function. This network of connected cells was then subject to Louvain clustering. The range of possible resolutions to identify these subpopulations was visualized using clustree R package. Resolution 0.5 was utilized for this analysis and throughput the study. This is comparable to widely used 0.6-0.8 range, which enables avoiding over-clustering [6]. For visualization, cell distances and their graph embedding were subject to uniform manifold approximation and projection (UMAP), where cells are plotted in two-dimensional feature-space, such that cells with similar transcriptional profiles will appear closer to one another. Further classification of the clusters was preformed using the FindAllMarkers function to identify differentially expressed genes (DEG) between clusters. These DEG were then additionally screened using violin plots for visualising their expression across all clusters.

**Analysis of Individual Cohorts Before Integration**

Clustering analysis of aging human lung tissue in two independent cohorts identified two closely related clusters of aging blood and lymphatic vessel EC (BEC and LEC respectively) in both (Additional file 2: Figure S1 A-B). Unsupervised sub-clustering analysis of the total aging human lung endothelium (BEC and LEC pooled together) revealed 5 sub-populations of EC in cohort 2, closely matching COPD cell atlas data that compared aging-matched datasets of human lung and identified 6 sub-populations of EC [7] (Additional file 2: Figure S1C-E). In contrast, unexpectedly 12 sub-populations of aging human lung EC were identified in cohort 1 (Additional file 2: Figure S1C-E), potentially reflecting the differences in sample collection sites (Table 1).

**Endothelial Cell Identification**

Following clustering and visualization of the human aging lung clusters using UMAP, we categorized cell types based on expression profile of classical lineage markers reported by others in the IPF cell atlas covering lung from various ages (21-80) [8]. Endothelial cell (EC) cluster was identified using classical (platelet endothelial cell adhesion molecule-1, PECAM1; vascular endothelial cadherin, CDH5; claudin 5, CLDN5) and novel (calcitonin-receptor-like receptor, CLR, encoded by *CALCRL* gene) pan-endothelial markers [9-13]. These were further classified into two sub-sets - blood vessel ECs (BEC; based on von Willebrand Factor, vWF, expression) and lymphatic vessel ECs (LEC; based on expression of prospero homeobox 1, PROX1; podoplanin, PDPN; lymphatic vessel endothelial hyaluronan receptor 1, LYVE1 and fms related receptor tyrosine kinase 4, FLT4). Two subsets consisting of only BEC or LEC were created and used for downstream analyses of physiologically aging lungs and IPF lungs from elderly patients. Essential quality controls (filtering data, normalization and endothelial identification; [14]) were conducted for the datasets from individual cohorts (Figure 1A) to confirm comparable quality (uniform reads and presence of clusters of lung cell types in integrated dataset; Figures 1B, Additional file 2: Figure S1) despite differences in used protocols.

**Cluster and Subpopulation Screening and Profiling**

The data was screened to assess the presence of clusters and subpopulation containing a low number of genes per cell [14]. The number of genes per cell for each cluster in the total lung data was plotted in a violin plot. All of the 25 identified in total aging human lung clusters were sub-set into their own SeuratObject and were re-clustered to identify subpopulation. All subpopulation of the aging lung were analysed for the number of genes per cell for each cluster. This information was used to confirm if the low number of genes per cell was cell type specific. The expression of a panel of endothelial progenitor markers (*CD34, PTPRC, KIT, POU5F1, MYC, PROM1, CXCR4, TEK, GYPA, NT5E* and *SPN*; [15]) in BEC subpopulation was also analysed using violin plots.

**Cell Trajectory Analysis**

We performed a pseudo-time–based cell trajectory analysis using the R package Slingshot [16]. The slingshot wrapper function was performed with the UMAP dimensionality reduction and cluster labels as in Seurat objects to identify the trajectory. The subclustered ECs were prepared for processing using the slingshot function as SingleCellExperiment function. The slingshot trajectories are calculated such that each trajectory has a single start and end point. For clarity and brevity, multiple trajectories were plotted on the same graph.

**Cell-Cycle Scoring**

A list of cell cycle markers was loaded from Seurat V4 and segregated into markers of G2/M or S phase [3, 39]. The Cell-Cycle Scoring function was utilised to assign each cell within the SeuratObject a ‘score’, based on its expression of G2/M and S phase markers. Cells expressing neither G2/M phase nor S phase markers are predicted to be not cycling and in G1 phase. Based on results of cell-cycle scoring, each cell was assigned a “cell-cycle score” (either G2/M, S or G1). The number of cells within each subpopulation with each of the three cell-cycle scores, i.e. in each of the three stages of the cell cycle, were then quantified.

**Cell Scoring Assay**

The Seurat object for the sub-set BEC and LEC were split using the Seurat SplitObject function by sample condition (donor and fibrosis). The objects were then queried using publicly available transcriptional signatures (GOBP cellular senescence (senescence), Hallmark apoptosis (apoptosis), GOBP endothelial cell proliferation (proliferation), GOBP blood vessel endothelial cell migration (migration), angiogenesis (angiogenesis), GOBP inflammation (inflammation), GOBP vasodilation (vasodilation) and GOBP regulation of vascular permeability (permeability)) from the GSEA website ([17]; <http://www.broad.mit.edu/gsea/>) developed by the BROAD institute. The libraries for EC differentiation, LEC differentiation and Endo MT were developed from literature. GSEA results should be interpreted as alterations to these processes, not simple increases or decreases as the values may suggest. Therefore, a change in expression of genes outlined in GSEA libraries is termed as a process “score” (s), e.g., EC differentiation or Endo-MT scores etc. The “scoring” was calculated based on the difference between the average expression levels of the gene set in a particular subpopulation compared to the total population of cells. “Positive” or “negative” (above or below 0 respectively) score would suggest that this group (module) of genes is expressed in a particular cell at a higher or lower level, than expected, respectively, given the average expression across the cell population. Individual cells were scored using the Seurat function AddModualScore. These scores were then visualized using the Seurat RidgePlot function. A full list of the genes used are summarized in Additional file 3: Table S7.

**Differential Gene Expression Analysis and Qiagen Ingenuity Pathway Analysis (IPA)**

The Seurat object for the sub-set BEC and LEC were further sub-set so that each cluster became a separate Seurat Object. The active identity of each cluster was then set to sample condition (donor or IPF groups). The Seurat function FindMarkers function was used to identify the differentially expressed genes (DEG) between donor and IPF groups. These genes were then exported into an .xlxs file, along with p-values, logFC and adjusted p-values. This was performed for every subpopulation. The lists of DEG were uploaded separately into Qiagen IPA software (QIAGEN Inc., <https://digitalinsights.qiagen.com/IPA>). A Core expression analysis function was run for each list, taking into account only genes with Wilcoxon rank sum test P < 0.05 and querying only human processes. The pathways which had a definitive z score were then summarized in Figure 7 C for BEC and Additional file 2: Figures S26 A, B for LEC. IPA's z-score indicates a predicted activation or inhibition of a pathway/gene, where a negative z value annotates an overall pathway's inhibition, and a positive z value annotates an overall pathway's activation.

**Statistical Analyses**

Analysis of cell type specific marker genes was performed using the FindAllMarkers function in Seurat v4. This utilizes the Wilcoxon rank sum test, with p values adjusted for multiple comparisons using the Bonferroni method. Adjusted p-values of <0.05 were considered significant. Significance of cell cycle analysis data between sample conditions was tested using a chi-square test. P < 0.05 was considered significant. Analysis of the cell scoring assay was performed using a Shapiro Wilcoxon test to test for normality and Mann-Whitney U test. P < 0.05 was considered significant. Analysis of cell numbers between donor and fibrosis was tested using multiple t tests. P < 0.05 was considered significant. Statistical analysis was performed in R or in Graph pad prism.

**Annotation of BEC Subpopulation**

Subclusters were annotated based on the specific transcriptional signature derived from heatmap analysis (Figure 2 B-D) and other information. Subcluster 0 was annotated as “de-differentiated” BEC due to its distinct clustering, as a part of BEC population, away from the rest of cell populations within the integrated single cell map of ageing human lungs (Figure 1B), either low (*PECAM*, *vWF,* *CLDN5* and *CALCRL*) or absent (*CDH5*) expression levels for general EC markers (Figures 2C, D), comparable to other subpopulations levels of the housekeeping gene *B2M* (Additional file 2: Figures S12 A) and pan-EC marker expression profiles (ridgeplots), whilst having a considerably larger proportion of cells with lower expression of *PECAM1* and *CALCRL* (Additional file 2: Figures S15 A, B), lower average number of genes per cell (Additional file 2: Figures S12 B), lack of expression of specific markers from heatmap (Figure 2 B, Additional file 2: Figure S5) and a close relationship to all other clusters, as revealed by pseudo-time lineage analysis (Figure 2E). Subpopulations 1 and 2 were named “bronchial venule 1 (IPF Endo 1)” and “bronchial venule 2 (IPF Endo 2)” BEC. Both clusters had distinct gene expression signatures revealed by heatmaps (Figure 2 B, D) and significant “bronchial” and “vein” identity scores (Figures 2F, Additional file 2: Figure S9). “Bronchial venule 1 (IPF Endo 1)” BEC sub-cluster partly, yet non-significantly, matched the “vascular endothelium (VE) peribronchial” sub-cluster signature (Figures 2F, Additional file 2: Figure S6) which was identified by others through analysis of age-unmatched datasets of lung tissues from donors and IPF patients [1]. Both subpopulations were annotated “IPF Endo” (1 or 2) due to the increased number of these EC sub-populations in IPF lung (Figure 3). These subpopulations were differentially present in two analysed cohorts (Figure 3A, D), thus intimately linking them to different parts of the lung (the cross-organ region and distal parenchyma in cohorts 2 and 1 respectively, Table 1). Sub-cluster 3 was annotated “general capillary” or “gCap” BEC due to its expression of capillary, and presence of gCap-specific markers *CD36*, *FCN3* and *IL7R* [18]; Additional file 2: Figure S6). Sub-cluster 4 was labelled as “intralobular arteriole” BEC due to its expression of classical arterial differentiation genes (e.g., *EFNB2*, *HEY1*, *GJA5*; [19, 20]; Figure 2F, Additional file 2: Figures S6). Sub-cluster 5 was labelled as “de-differentiated capillary” BEC due to either extremely low (*PECAM*, *vWF* and *CLDN5*) or absent (*CDH5* and *CALCRL*) expression levels for general EC markers (Figures 2C, D), lower average number of genes per cell and lack of expression of specific markers (Additional file 2: Figures S9), and a close relationship to general capillary as revealed by pseudo-time lineage analysis (Figure 2E). Sub-cluster 6 was annotated as “intralobular venule” BEC based on expression of venous differentiation genes (e.g., *NR2F2*, *SOX18* and *TAGLN*; [19, 20]; Figure 2F, Additional file 2: Figures S6). Sub-cluster 7 was labelled as “bronchial capillary” BEC due to its positive scoring for “bronchial” genes signature ([21]; Figures 2F, Additional file 2: S9), similarity to the “general capillary” BEC sub-cluster (Figures 2F, Additional file 2: Figures S6) and high expression of *CD34,* known to be high in lung alveolar capillaries ([12]; Additional file 2: Figures S7). Cluster 8 was labelled as “aerocyte” BEC based on its expression of aerocyte-specific markers (*IL1RL1*, *AFF3*, *CA4* and *NCALD*; [18]; Figure 2F, Additional file 2: Figures S6). Sub-cluster 9 was annotated as “inflammatory” BEC due to high expression of *IL6*, *SELE* and *ICAM1* ([22]; Figure 2D), as also identified in other reports [25]).

The sub-cluster 10 BEC subpopulation expresses pan-EC at similar levels to other BEC subpopulations (Figure 2C, D). It is present in both the young and the ageing human lung (as sub-cluster L; Additional file 2: Figures S13, S14E). The cluster was predominantly present in the distal lung (cohort 1; Figure 3D), at approximately 2-7% in the donor and 0-1% in the IPF (Additional file 3: Table S5). This subpopulation has a distinct transcriptional signature including *SFTPA1, SFTPB, NAPSA* and *SLIP* genes (Figures 2B, D; Additional file 2: Figure S3) expressed at lower levels compared to any clusters in the total lung map (Additional file 2: Figure S4A). EC subpopulation expressing *SFTPA1, SFTPC, SFTPD, KRT7* and *AGER* genes was recently identified in murine lung and termed “sftp+” EC or “EC-pneumocyte” [26, 27]. Analysis of expression of these genes in ageing human lung BEC sub-populations revealed their predominant expression in subpopulation 10 (Additional file 2: Figure S10A). Pseudo-time lineage analysis showed that BEC subpopulation 10 had closest proximity to intralobular venule cluster 6 and inflammatory cluster 9 (Figure 2E). Based on all these findings, we annotated this BEC subpopulation in ageing human lung as “Pneumocyte marker (low) expressing” (Figure 4A).

The sub-cluster 11 BEC subpopulation expresses pan-EC at similar levels to other BEC subpopulations (Figure 2C, D). It is present in both the young and the ageing human lung (as sub-cluster M; Additional file 2: Figures S13, S14E). The subpopulation was mainly present in the distal lung (cohort 1; Figure 3D), at approximately 1-6% in the donor and 0-1.4% in the IPF (Additional file 3: Table S5). This subpopulation has a distinct transcriptional signature including *HLA-DPB1, C1QA* and *LYZ* genes (Figures 2B, D; Additional file 2: Figure S3) expressed at lower levels than other clusters in the total lung map (Additional file 2: Figure S4A). EC subpopulation expressing *C1QA, CTSS, CTSD, CTSB* and *CTSZ* genes was recently identified in porcine endothelium and termed “immune-active EC” [28]. Analysis of expression of these genes in ageing human lung BEC sub-populations revealed their predominant expression in subpopulation 11 (Additional file 2: Figure S10B). Pseudo-time lineage analysis showed that the BEC subpopulation also has a close proximity to intralobular arteriole sub-cluster 4 (Figure 2E). The subpopulation had higher expression levels of large vessel markers *NOTCH1,* *HEXB* and *S100A4* ([23, 24]; Figure 2F, Additional file 2: Figure S7B), with the latter being also associated with murine and human EC de-differentiation/Endo-MT in *in vitro* models [25, 26]. Subpopulation 11 also had the highest average EndoMT module score of all BEC sub-populations (Additional file 2: Figure S20 K). Finally, the BEC subpopulation 11 expresses EC progenitor marker genes *CD34*, *RUNX1*, *CXCR4*, *PTPRC, SPN, PPARG* and others ([29, 30]; Additional file 2: Figure S12D). Based on these findings, we annotated the BEC Subpopulation 11 as “Immune marker (low) - expressing”.

Also, subpopulations 10 and 11 clustered with “macrovascular” BEC in the dendrogram (unsupervised hierarchical cluster tree) for both donor and IPF groups (Figure 4B). Subpopulations 10 and 11 were mainly present in cohort 1 (Figures 3 C-E), thus intimately linking their location, and hence origin, to distal parenchyma (Table 1).

**Annotation of LEC Subpopulations**

To comprehensively decipher the transcriptional signature of aging human lung lymphatic vessel endothelium in IPF, we extracted scRNAseq data for LEC from the total lung from two cohorts in both conditions (689 cells) and performed unsupervised re-clustering (Figure 1A), which revealed two main populations consisting of five distinct subpopulations in total (Figures 1A, 8A). Expression analysis of pan-EC markers and LEC-specific genes (*PROX1*, *PDPN*, *LYVE-1* and *FLT4*) and *CCL21* was done to confirm the identities of all cell subpopulations as LEC (Figure 8C). Combined with expression profiles of five genes known to be expressed in LEC (*ACKR2, GJA1, NFACT1, CD34* and *NR2F2*; [9, 27]) and 22 DEG from the heat map (Figure 8B, D, Additional file 2: Figures S21, S22), cell cycle analysis data (Additional file 2: Figures S23) and tissue sample collection locations (Table 1), these data were deemed insufficient basis for the annotation for each of five subpopulation of LEC beyond their numbering and detailed transcriptional characterisation (Figure 8D).

**Formalin Fixed Paraffin Embedded Tissue Section Selection and Sectioning**

A formalin fixed paraffin embedded (FFPE) tissue section of distal lung diagnostic biopsy of IPF (with confirmed histopathological pattern of usual interstitial pneumonia) from 58 years old subject was used for the spatial validation of scRNAseq findings in distal lung in this study by immunohistochemistry and quantitative high content image analysis. Analysis of surplus human lung biopsy tissue was approved by the Hull and East Riding Research Ethics Committee (reference 08/H1304/54). Two FFPE blocks of tissue from the same patient, both distal lung parenchyma, were used for downstream analysis. Sections were cut at 4µm thickness and transferred to a 42°C water bath where the sections were caught on Vectabond coated, Starfrost slides. The slides were dried overnight at 56 °C in an oven.

**Mason’s Trichrome staining**

Mason’s Trichrome staining was performed using a modified method [31]. The FFPE sections were deparaffinised in histo-clear and rehydrated through an ethanol dilution series (100, 95, 75, 0% ethanol in ddH2O) before processing. The sections were stained with Weigerts haematoxylin by adding equal quantities of solution A and solution B to a test tube and mixing well then adding to the section and staining for 30 min. The nuclei were differentiated with 1% acid alcohol (IDA99). Followed by staining with 0.5% acid fuchsin in 0.5% acetic acid for 1 min. This was also differentiated with 1% phosphomolybdic acid until collagen was decolourised (with muscle and red blood cells (RBC’s) remaining red) for up to 5 mins depending on section thickness. Finally, the sections were counterstained with 0.2% light green in 2% acetic acid for 3 minutes. This was differentiated with 1% acetic acid to fix and differentiate the light green for 1 min. The sections were then dehydrated through an ethanol series and cleared using histo-clear. Finally, the sections were mounted using DPX mounting medium (Agar, R1340) and a 50mm glass cover slip before being imaged.

**Immunohistochemistry**

Immunohistochemistry was performed using a method based on manufacturer recommendations, with minor modifications. The sections were stained using the Dako EnVision+ Dual Link System-HRP (DAB+) kit (Agilent, #K4065). Briefly the FFPE sections were deparaffinised in histo-clear and rehydrated through an ethanol dilution series (100, 95, 75, 0% ethanol in ddH2O) before processing. The sections then underwent appropriate epitope retrieval of microwaving in either 10 mM Citrate buffer (pH 6) or Tris (10 nM) /EDTA (1 mM) (pH 9) buffer for 3x 3 minutes with a 20 min cooling step in between microwaving. The sections were then pre-blocked using the Dako Dual endogenous enzyme block solution for ten minutes. The sections were treated at a respective appropriate concentration and overnight for each primary antibody (rabbit anti-human CLR, 1:1500, LN1436 [32]; mouse anti-human CD31, 1:100, JC/70A, Novus biological; mouse anti-human PDPN, 1:100, D2-40, Agilent) and were incubated with the labelled polymer-HRP secondary for 30 minutes. The colour was developed with DAB+ chromogen and DAB+ substrate buffer solution mixture, with development monitored under a microscope. Sections were counterstained with haematoxylin solution according to Delafield (Sigma-Aldrich, #03971) for 2 seconds. The sections were then dehydrated through an ethanol series and cleared using histo-clear. Finally, the sections were mounted using DPX mounting medium before being imaged. Controls included omission of the primary or secondary antibody. In all cases, control sections showed no staining.

**Sudan Black B staining**

Sudan Black B (SBB) staining was performed using a modified method [33]. In brief, the procedure was as follows.

*Preparation of SBB solution.* 0.7 gram of SBB (Hopkins and Williams, 26150) was dissolved in 100ml of 70% ethanol, covered with parafilm and thoroughly stirred overnight at room temperature. The solution was then filtered through filter paper and then filtered again through a frittered glass filter of medium porosity with suction. Throughout the process, it was important to avoid ethanol evaporation, which results in precipitation of the stain, so the solution was stored in an airtight container.

*Preparation of Nuclear Fast Red (NFR) solution.* 0.1 gram of NFR (Gurr’s) and 5gram of Aluminium Sulphate was dissolved in 100ml of ddH2O ethanol thoroughly stirred. The solution was slowly brought to the boil before being allowed to cool to room temperature. The solution was then filtered through filter paper.

*Staining Procedure.* The FFPE sections were deparaffinised in histo-clear and rehydrated through an ethanol dilution series (100, 95, 75% ethanol in ddH2O) before processing. A drop from freshly prepared SBB was dropped on a clean slide between two towers of three coverslips at either end of the slide. The tissue section was placed facing down on the drop of SBB on the slide and incubated at room temperature for 8 minutes until the desirable staining with no precipitation was accomplished. The staining was observed under the microscope. The slide, was carefully lifted and the SBB on the edges of the slide was wiped out manually from the back and along the edges of the slide with the help of a soft paper. The slide was then placed into 50% ethanol, transferred and washed in distilled water, counterstained with NFR for 10 min., and mounted into 40% Glycerol/TBS mounting medium. Lipofuscin staining was considered positive when perinuclear and cytoplasmic aggregates of blue-black granules were evident inside the cells.

**High Content Image J Quantification of Vessel Nuclei in Immunostained Tissues**

Stained tissue sections were scanned using the Zeiss Axioscan Z1 Slide Scanner at the University of York Biotechnical Facility. Regions of interest (ROI) were selected from either the scanned section using the QuPath analysis software [34] or by light microscopy using a Zeiss Axio Lab A1. The ROI were saved as TIF files and transferred to Fiji/ImageJ for further analysis. The images were then analysed using high content image quantification pipeline [35]. The images were opened in Image J and areas around selected ROI removed, thus leaving only the selected ROI for downstream analysis. The ROI were split into blue or brown colour components using the colour deconvolution function with the HDAB subfunction for the analysis of hematoxylin (blue) or DAB (brown) signals. The deconvoluted image was then subjected to a threshold analysis using the Otsu function to generate maps for both hematoxylin or DAB signals. An overlap map was created using the image composite function and XOR subfunction. The generated composite map was analysed using the “Analyse particals” function for particles between 10 and 1000 with a circularity of 0.5. The ROI width was determined by taking 7 measurements of width selected at random from across the ROI and the average was calculated. The vessel diameter measurement to determine capillary (<7µm) or venule (>7µm) were taken following published examples [36-38]. Briefly, a line was drawn using Image J, across the vessel lumen and the image J measure function was used to report the length in µm of the line. Finally, individual bronchi were identified within the total section under microscope and selected for downstream analysis to quantify the total section areas in mm^2^ and total number of bronchi per section, and percentage of CLR-positive nuclei in total section.

**References**

1.	Adams TS, Schupp JC, Poli S, Ayaub EA, Neumark N, Ahangari F, et al. Single-cell RNA-seq reveals ectopic and aberrant lung-resident cell populations in idiopathic pulmonary fibrosis. Science advances. 2020;6(28):eaba1983.

2.	Reyfman PA, Walter JM, Joshi N, Anekalla KR, McQuattie-Pimentel AC, Chiu S, et al. Single-cell transcriptomic analysis of human lung provides insights into the pathobiology of pulmonary fibrosis. American journal of respiratory and critical care medicine. 2019;199(12):1517-36.

3.	Hao Y, Hao S, Andersen-Nissen E, Mauck W, Zheng S, Butler A, et al. Integrated analysis of multimodal single-cell data. bioRxiv; 2020.

4.	Tran HTN, Ang KS, Chevrier M, Zhang X, Lee NYS, Goh M, Chen J. A benchmark of batch-effect correction methods for single-cell RNA sequencing data. *Genome Biol.* 2020;21(1):12.

5.	Stuart T, Satija R. Integrative single-cell analysis. Nat Rev Genet. 2019;20(5):257-72.

6.	Luecken MD, Theis FJ. Current best practices in single-cell RNA-seq analysis: a tutorial. Mol Syst Biol. 2019;15(6):e8746.

7.	Sauler M, McDonough JE, Adams TS, Kothapalli N, Barnthaler T, Werder RB, et al. Characterization of the COPD alveolar niche using single-cell RNA sequencing. Nature Communications. 2022;13(1):1-17.

8.	Neumark N, Cosme Jr C, Rose K-A, Kaminski N. The idiopathic pulmonary fibrosis cell atlas. American Physiological Society Bethesda, MD; 2020. p. L887-L92.

9.	Johnson NC, Dillard ME, Baluk P, McDonald DM, Harvey NL, Frase SL, et al. Lymphatic endothelial cell identity is reversible and its maintenance requires Prox1 activity. Genes & development. 2008;22(23):3282-91.

10.	Müller AM, Hermanns MI, Skrzynski C, Nesslinger M, Müller KM, Kirkpatrick CJ. Expression of the endothelial markers PECAM-1, vWf, and CD34 in vivo and in vitro. Exp Mol Pathol. 2002;72(3):221-9.

11.	Nikitenko LL, Smith DM, Bicknell R, Rees MC. Transcriptional regulation of the CRLR gene in human microvascular endothelial cells by hypoxia. The FASEB journal. 2003;17(11):1-25.

12.	Pusztaszeri MP, Seelentag W, Bosman FT. Immunohistochemical expression of endothelial markers CD31, CD34, von Willebrand factor, and Fli-1 in normal human tissues. J Histochem Cytochem. 2006;54(4):385-95.

13.	Wigle JT, Harvey N, Detmar M, Lagutina I, Grosveld G, Gunn MD, et al. An essential role for Prox1 in the induction of the lymphatic endothelial cell phenotype. Embo j. 2002;21(7):1505-13.

14.	Durante MA, Kurtenbach S, Sargi ZB, Harbour JW, Choi R, Kurtenbach S, et al. Single-cell analysis of olfactory neurogenesis and differentiation in adult humans. Nat Neurosci. 2020;23(3):323-6.

15.	Hristov M, Erl W, Weber PC. Endothelial progenitor cells: mobilization, differentiation, and homing. Arterioscler Thromb Vasc Biol. 2003;23(7):1185-9.

16.	Street K, Risso D, Fletcher RB, Das D, Ngai J, Yosef N, et al. Slingshot: cell lineage and pseudotime inference for single-cell transcriptomics. BMC genomics. 2018;19(1):1-16.

17.	Subramanian A, Tamayo P, Mootha VK, Mukherjee S, Ebert BL, Gillette MA, et al. Gene set enrichment analysis: a knowledge-based approach for interpreting genome-wide expression profiles. Proc Natl Acad Sci U S A. 2005;102(43):15545-50.

18.	Gillich A, Zhang F, Farmer CG, Travaglini KJ, Tan SY, Gu M, et al. Capillary cell-type specialization in the alveolus. Nature. 2020;586(7831):785-9.

19.	Adams RH. Molecular control of arterial–venous blood vessel identity. Journal of Anatomy. 2003;202(1):105-12.

20.	Swift MR, Weinstein BM. Arterial-venous specification during development. Circ Res. 2009;104(5):576-88.

21.	Travaglini KJ, Nabhan AN, Penland L, Sinha R, Gillich A, Sit RV, et al. A molecular cell atlas of the human lung from single-cell RNA sequencing. Nature. 2020;587(7835):619-25.

22.	Pober JS, Sessa WC. Evolving functions of endothelial cells in inflammation. Nature Reviews Immunology. 2007;7(10):803-15.

23.	Iso T, Kedes L, Hamamori Y. HES and HERP families: multiple effectors of the Notch signaling pathway. J Cell Physiol. 2003;194(3):237-55.

24.	Nukala SB, Tura-Ceide O, Aldini G, Smolders V, Blanco I, Peinado VI, et al. Protein network analyses of pulmonary endothelial cells in chronic thromboembolic pulmonary hypertension. Sci Rep. 2021;11(1):5583.

25.	Herwig N, Belter B, Pietzsch J. Extracellular S100A4 affects endothelial cell integrity and stimulates transmigration of A375 melanoma cells. Biochemical and Biophysical Research Communications. 2016;477(4):963-9.

26.	Bondareva O, Rodríguez-Aguilera JR, Oliveira F, Liao L, Rose A, Gupta A, Singh K, Geier F, Schuster J, Boeckel JN, Buescher JM, Kohli S, Klöting N, Isermann B, Blüher M, Sheikh BN. Single-cell profiling of vascular endothelial cells reveals progressive organ-specific vulnerabilities during obesity. Nat Metab. 2022 Nov;4(11):1591-1610

27.	Rodor J, Chen SH, Scanlon JP, Monteiro JP, Caudrillier A, Sweta S, Stewart KR, Shmakova A, Dobie R, Henderson BEP, Stewart K, Hadoke PWF, Southwood M, Moore SD, Upton PD, Morrell NW, Li Z, Chan SY, Handen A, Lafyatis R, de Rooij LPMH, Henderson NC, Carmeliet P, Spiroski AM, Brittan M, Baker AH. Single-cell RNA sequencing profiling of mouse endothelial cells in response to pulmonary arterial hypertension. Cardiovasc Res. 2022 Aug 24;118(11):2519-2534.

28.	Wang F, Ding P, Liang X, Ding X, Brandt CB, Sjöstedt E, Zhu J, Bolund S, Zhang L, de Rooij LPMH, Luo L, Wei Y, Zhao W, Lv Z, Haskó J, Li R, Qin Q, Jia Y, Wu W, Yuan Y, Pu M, Wang H, Wu A, Xie L, Liu P, Chen F, Herold J, Kalucka J, Karlsson M, Zhang X, Helmig RB, Fagerberg L, Lindskog C, Pontén F, Uhlen M, Bolund L, Jessen N, Jiang H, Xu X, Yang H, Carmeliet P, Mulder J, Chen D, Lin L, Luo Y. Endothelial cell heterogeneity and microglia regulons revealed by a pig cell landscape at single-cell level. Nat Commun. 2022 Jun 24;13(1):3620.

29.	Timmermans F, Plum J, Yöder MC, Ingram DA, Vandekerckhove B, Case J. Endothelial progenitor cells: identity defined? J Cell Mol Med. 2009 Jan;13(1):87-102.

30.	Abdelgawad ME, Desterke C, Uzan G, Naserian S. Single-cell transcriptomic profiling and characterization of endothelial progenitor cells: new approach for finding novel markers. Stem Cell Res Ther. 2021 Feb 24;12(1):145.

31.	Bancroft JDaG, M. (Ed.). Theory and Practice of Histological Techniques: Churchill Livingstone; 2008.

32.	Nikitenko LL, Blucher N, Fox SB, Bicknell R, Smith DM, Rees MC. Adrenomedullin and CGRP interact with endogenous calcitonin-receptor-like receptor in endothelial cells and induce its desensitisation by different mechanisms. J Cell Sci. 2006;119(Pt 5):910-22.

33.	Georgakopoulou EA, Tsimaratou K, Evangelou K, Fernandez Marcos PJ, Zoumpourlis V, Trougakos IP, et al. Specific lipofuscin staining as a novel biomarker to detect replicative and stress-induced senescence. A method applicable in cryo-preserved and archival tissues. Aging (Albany NY). 2013;5(1):37-50.

34.	Bankhead P, Loughrey MB, Fernández JA, Dombrowski Y, McArt DG, Dunne PD, et al. QuPath: Open source software for digital pathology image analysis. Scientific Reports. 2017;7(1):16878.

35.	Crowe AR, Yue W. Semi-quantitative Determination of Protein Expression using Immunohistochemistry Staining and Analysis: An Integrated Protocol. Bio Protoc. 2019;9(24).

36.	Gaikwad AV, Lu W, Dey S, Bhattarai P, Chia C, Larby J, et al. Vascular remodelling in idiopathic pulmonary fibrosis patients and its detrimental effect on lung physiology: potential role of endothelial-to-mesenchymal transition. ERJ Open Res. 2022;8(1).

37.	Hall SM, Hislop AA, Haworth SG. Origin, differentiation, and maturation of human pulmonary veins. Am J Respir Cell Mol Biol. 2002;26(3):333-40.

38.	Townsley MI. Structure and composition of pulmonary arteries, capillaries, and veins. Compr Physiol. 2012;2(1):675-709.

39.	Tirosh I, Izar B, Prakadan SM, Wadsworth MH 2nd, Treacy D, Trombetta JJ, Rotem A, Rodman C, Lian C, Murphy G, Fallahi-Sichani M, Dutton-Regester K, Lin JR, Cohen O, Shah P, Lu D, Genshaft AS, Hughes TK, Ziegler CG, Kazer SW, Gaillard A, Kolb KE, Villani AC, Johannessen CM, Andreev AY, Van Allen EM, Bertagnolli M, Sorger PK, Sullivan RJ, Flaherty KT, Frederick DT, Jané-Valbuena J, Yoon CH, Rozenblatt-Rosen O, Shalek AK, Regev A, Garraway LA. Dissecting the multicellular ecosystem of metastatic melanoma by single-cell RNA-seq. Science. 2016 ;352(6282):189-96.
