## Additional file 2: Supplementary figures for "Altered Heterogeneity of Ageing Lung Endothelium is a Hallmark of Idiopathic Pulmonary Fibrosis"

### FIGURE S1

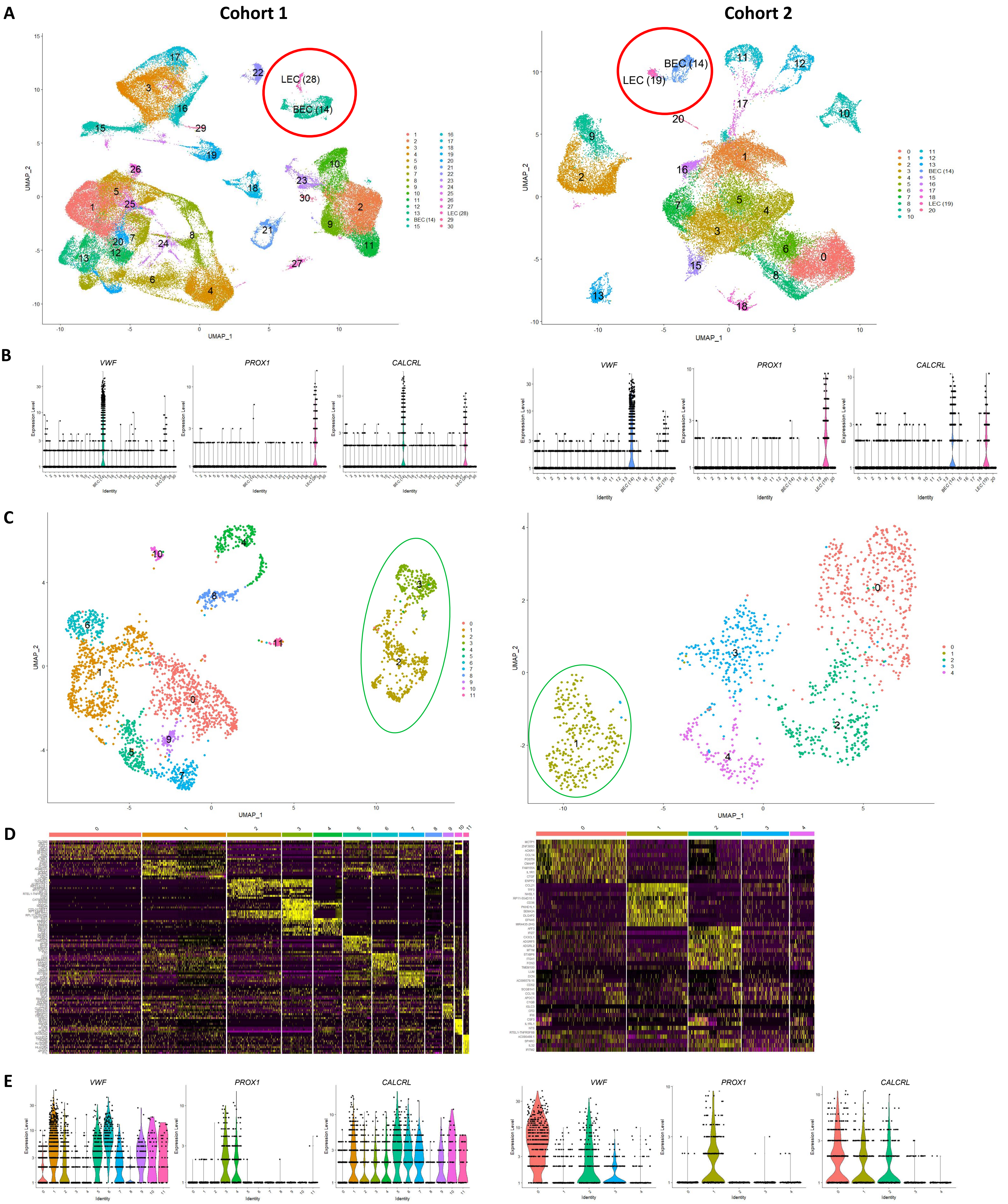
A
Cohort 1
Cohort 2
LEC (28)
BEC (14)
BEC (14)
LEC (19)
UMAP_1
UMAP_2
B
VWF
PROX1
CALCRL
Expression Level
Identity
C
D
E

### FIGURE S2 (to be continued on next page)

**A**

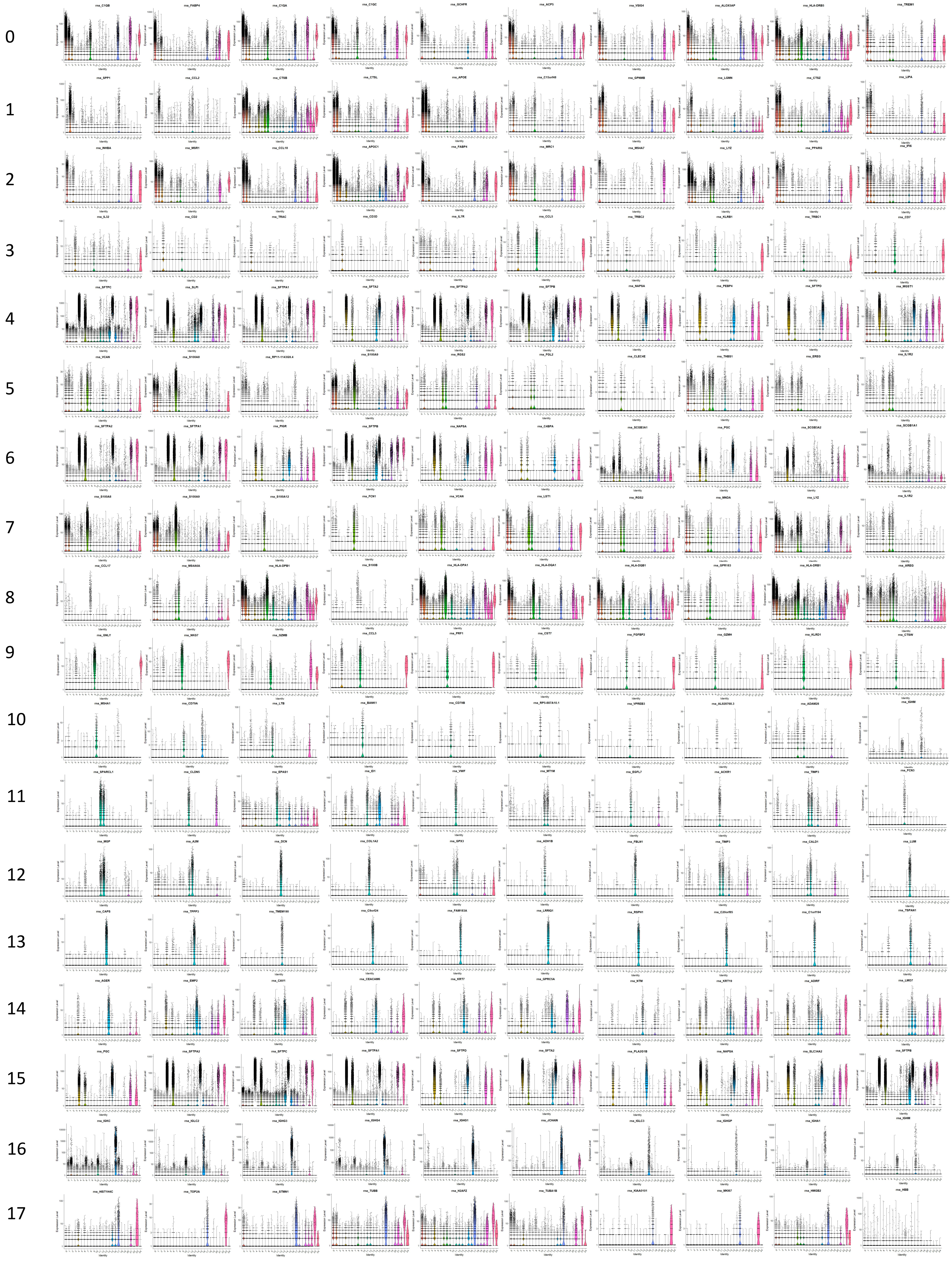

### FIGURE S2 (continued from previous page)

18
19
20
21
22
23
24

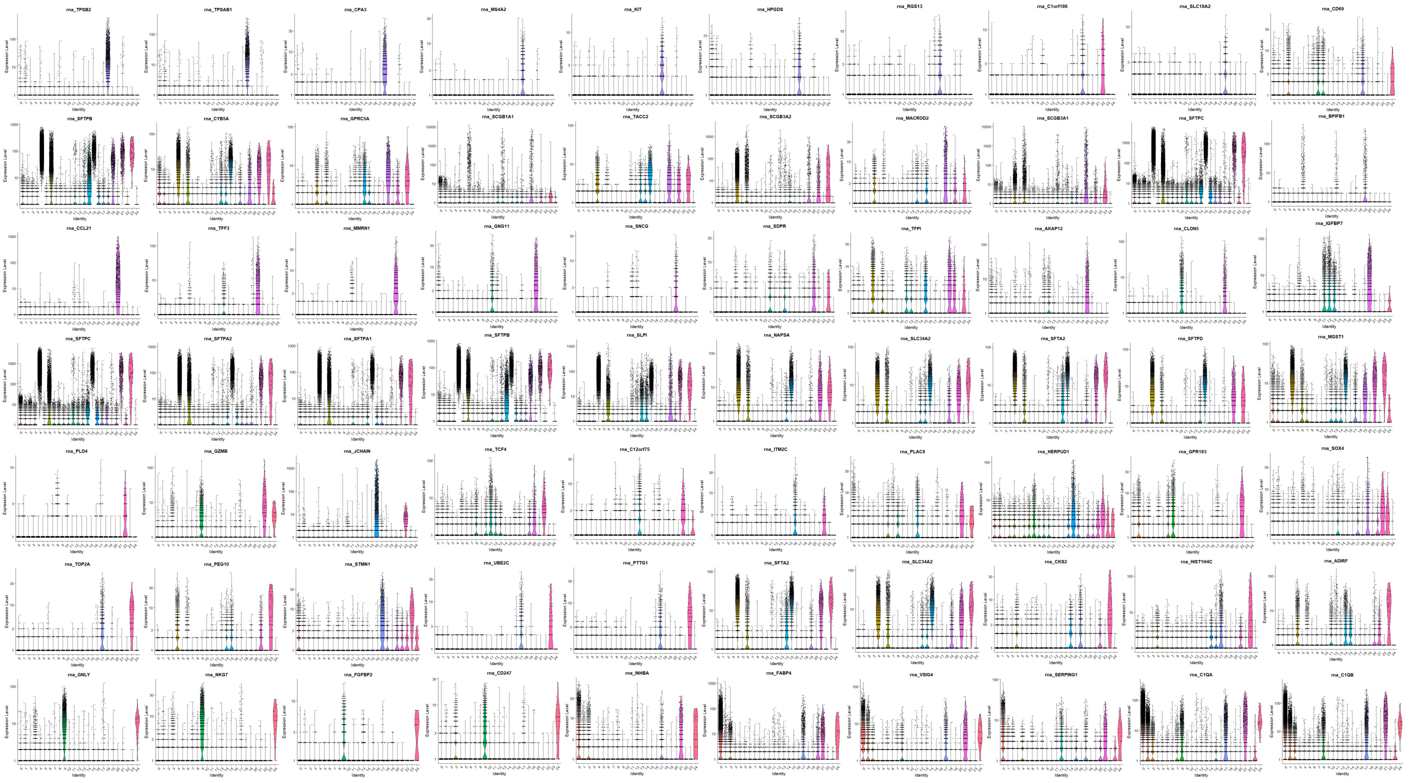


**B**

Total lung

Genes per cell

Molecules per cell

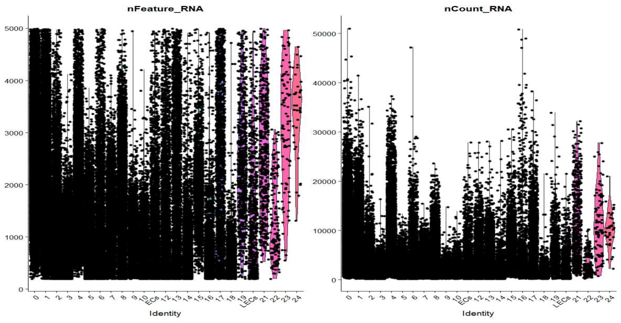

### FIGURE S3

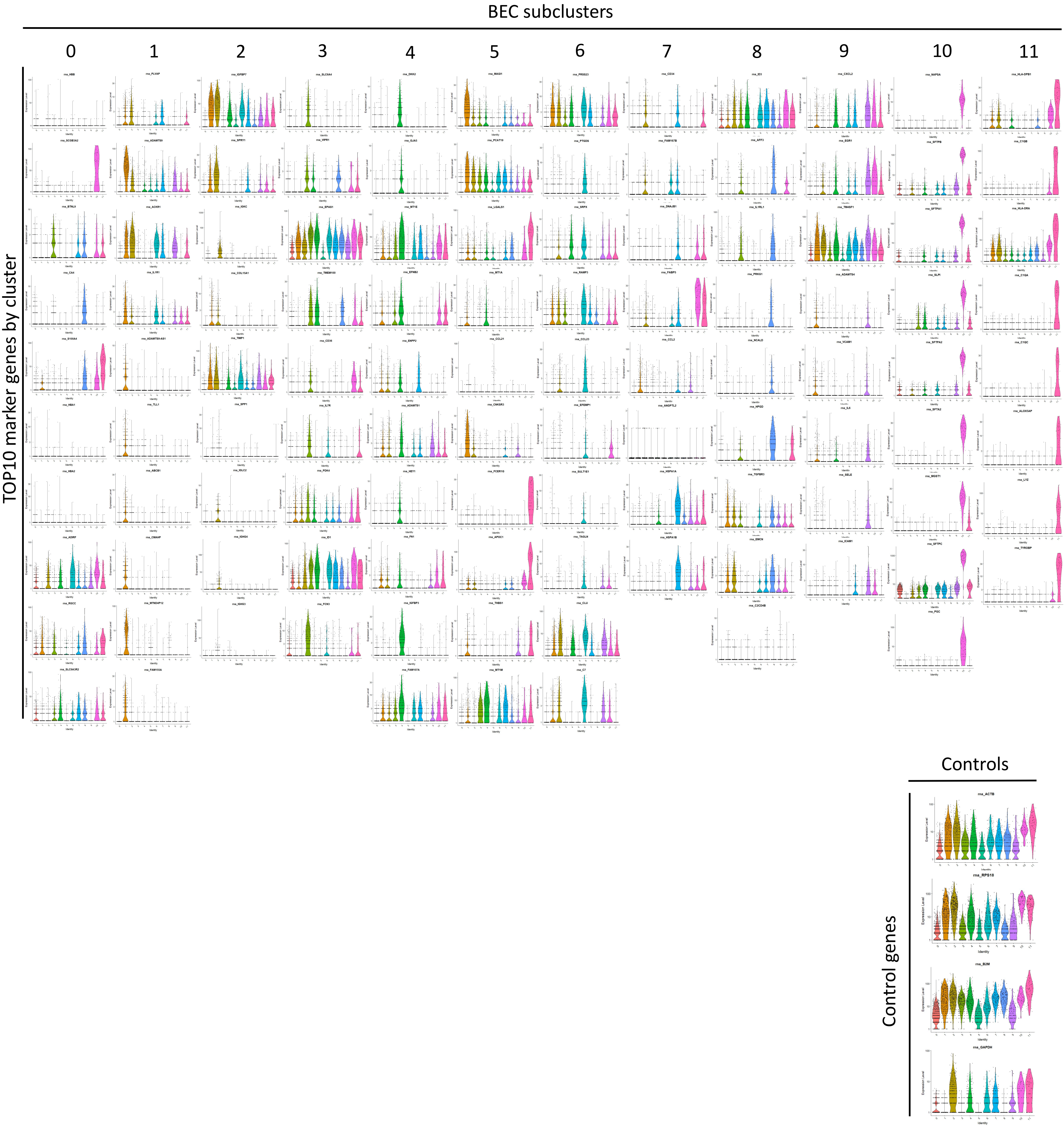
BEC subclusters
0
1
2
3
4
5
6
7
8
9
10
11
TOP10 marker genes by cluster
Controls
Control genes
rna_ACTB
rna_RPS18
rna_B2M
rna_GAPDH

### FIGURE S4

**A**

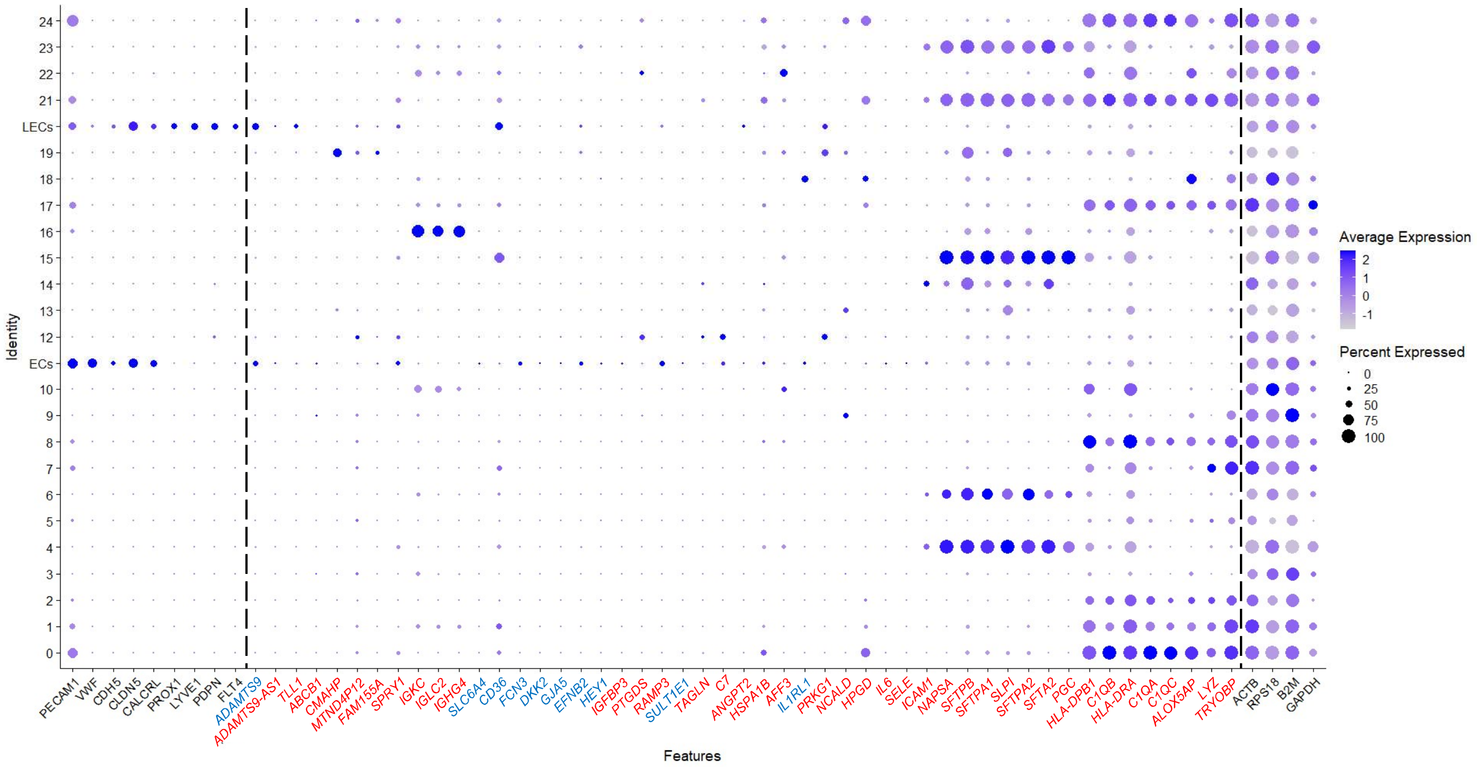


**B**

| Key | BEC sub-cluster number | Gene ID | Name | Gene Cards Classification | IPA Classification | IPA Location |
|---|---|---|---|---|---|---|
| | 1 | *ADAMTS9* | ADAM Metallopeptidase With Thrombospondin Type 1 Motif 9 | Secreted mammalian metalloprotease | Peptidase | Extracellular space |
| | | *ADAMTS9-AS1* | ADAMTS9 Antisense RNA 1 | Long non-coding RNA | Other | Other |
| | | *TLL1* | Tolloid Like 1 | Astacin-lke, zinc-dependent, metalloprotease | Peptidase | Extracellular space |
| | | *ABCB1* | ATP Binding Cassette Subfamily B Member 1 | Membrane-associated protein | Transporter | plasma membrane |
| | | *CMAHP* | Cytidine Monophospho-N-Acetylneuraminic Acid Hydroxylase, Pseudogene | Acid Hydroxylase | Other | Cytoplasm |
| | | *MTND4P12* | MT-ND4 Pseudogene 12 | Oxidoreductase enzyme | Other | Other |
| | | *FAM155A* | NALCN Channel Auxiliary Factor 1 | Transmembrane protein | Other | Other |
| | 2 | *SPRY1* | Sprouty RTK Signaling Antagonist 1 | Signalling antagonist | Other | Cytoplasm |
| | | *IGKC* | Immunoglobulin Kappa Constant | Immunoglobulins | Other | Extracellular space |
| | | *IGLC2* | Immunoglobulin Lambda Constant 2 | Immunoglobulins | Other | Extracellular space |
| | | *IGHG4* | Immunoglobulin Heavy Constant Gamma 4 (G4m Marker) | Immunoglobulins | Other | Extracellular space |
| | 3 | *SLC6A4* | Solute Carrier Family 6 Member 4 | Integral membrane protein | Transporter | Plasma membrane |
| | | *CD36* | CD36 Molecule | Receptor for thrombospondin | Transmembrane receptor | Plasma membrane |
| | | *FCN3* | Ficolin 3 | Serum protein | Other | Extracellular space |
| | 4 | *DKK2* | Dickkopf WNT Signaling Pathway Inhibitor 2 | Secreted protein | Other | Extracellular space |
| | | *GJA5* | Gap Junction Protein Alpha 5 | Component of gap junctions | Transmembrane receptor | Plasma membrane |
| | | *EFNB2* | Ephrin B2 | EPH-related receptor | Other | Plasma membrane |
| | | *HEY1* | Hes Related Family BHLH Transcription Factor With YRPW Motif 1 | Transcription regulator | Transcription regulator | Nucleus |
| | | *IGFBP3* | Insulin Like Growth Factor Binding Protein 3 | Insulin-like growth factor binding protein | Growth factor | Extracellular space |
| | 6 | *PTGDS* | Prostaglandin D2 Synthase | Enzyme | Enzyme | Cytoplasm |
| | | *RAMP3* | Receptor Activity Modifying Protein 3 | Single-transmembrane-domain proteins | G-protien coupled receptor | Plasma membrane |
| | | *SULT1E1* | Sulfotransferase Family 1E Member 1 | Enzyme | Other | Cytoplasm |
| | | *TAGLN* | Transgelin | Actin-binding protein | Other | Cytoplasm |
| | | *C7* | Complement C7 | Glycoprotein in MHC | Other | Extracellular space |
| | 7 | *ANGPT2* | Angiopoietin 2 | Antagonist of angiopoietin 1 | Growth factor | Extracellular space |
| | | *HSPA1B* | Heat Shock Protein Family A (Hsp70) Member 1B | Heat shock protein | Enzyme | Cytoplasm |
| | | *CD34* | CD34 | Pass membrane protein | Other | Plasma membrane |
| | 8 | *AFF3* | AF4/FMR2 Family Member 3 | Nuclear transcriptional activator | Transcription regulator | Nucleous |
| | | *IL1RL1* | Interleukin 1 Receptor Like 1 | Receptor | Transcription regulator | Plasma membrane |
| | | *PRKG1* | Protein Kinase CGMP-Dependent 1 | Protein kinase | Kinase | Cytoplasm |
| | | *NCALD* | Neurocalcin Delta | Calcium-binding protein | Other | Plasma membrane |
| | | *HPGD* | 15-Hydroxyprostaglandin Dehydrogenase | Dehydrogenase enzyme | Enzyme | Cytoplasm |
| | 9 | *IL6* | Interleukin 6 | Cytokine | Cytokine | Extracellular space |
| | | *SELE* | Selectin E | Cell adhesion molecule | Transmembrane receptor | Plasma membrane |
| | | *ICAM1* | Intercellular Adhesion Molecule 1 | Cell adhesion molecule | Transmembrane receptor | Plasma membrane |
| | 10 | *NAPSA* | Napsin A Aspartic Peptidase | Peptidase enzyme | Peptidase | Extracellular space |
| | | *SFTPA1* | Surfactant Protein A1 | Surfactant Protein | Other | Cytoplasm |
| | | *SLPI* | Secretory Leukocyte Peptidase Inhibitor | Secreted inhibitor | Other | Cytoplasm |
| | | *SFTPA2* | Surfactant Protein A2 | Surfactant Protein | Other | Cytoplasm |
| | | *SFTA2* | Surfactant Associated 2 | Surfactant Protein | Other | Cytoplasm |
| | | *PGC* | Progastricsin | Digestive enzyme | Enzyme | Cytoplasm |
| | 11 | *HLA-DPB1* | Major Histocompatibility Complex, Class II, DP Beta 1 | Membrane molecule | Transmembrane receptor | Plasma membrane |
| | | *C1QB* | Complement C1q B Chain | Complement protien | Other | Extracellular space |
| | | *HLA-DRA* | Major Histocompatibility Complex, Class II, DR Alpha | Membrane molecule | Transmembrane receptor | Plasma membrane |
| | | *C1QA* | Complement C1q A Chain | Complement protien | Other | Extracellular space |
| | | *C1QC* | Complement C1q C Chain | Complement protien | Other | Extracellular space |
| | | *ALOX5AP* | Arachidonate 5-Lipoxygenase Activating Protein | Arachidonic acid metabolite | Enzyme | Plasma membrane |
| | | *LYZ* | Lysozyme | Human lysozyme | Enzyme | Extracellular space |
| | | *TYROBP* | Transmembrane Immune Signaling Adaptor TYROBP | Transmembrane signaling polypeptide | Transmembrane receptor | Plasma membrane |

**C**

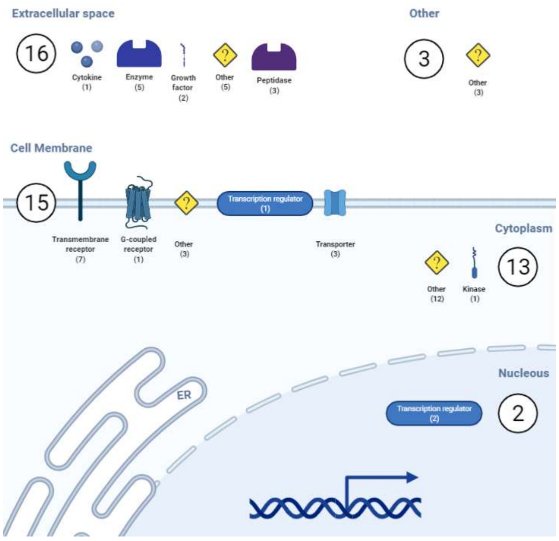


**D**

| Key | LEC sub-cluster number | Gene ID | Name | Gene Cards Classification |
|---|---|---|---|---|
| | 0 | *PDK4* | Pyruvate Dehydrogenase Kinase 4 | a mitochondrial protein |
| | | *ISG15* | Ubiquitin Like Modifier | ubiquitin-like protein |
| | | *SAMD4A* | Sterile Alpha Motif Domain Containing 4A | post-transcriptional regulator |
| | | *DLGAP2* | DLG Associated Protein 2 | membrane-associated protein |
| | 1 | *SFTPC* | Surfactant Protein C | pulmonary-associated surfactant protein C |
| | | *SCGB1A1* | Secretoglobin Family 1A Member 1 | small secreted proteins |
| | | *SFTPA2* | Surfactant Protein A2 | Surfactant Protein |
| | | *SFTPA1* | Surfactant Protein A1 | Surfactant Protein |
| | 2 | *IGFBP5* | Insulin Like Growth Factor Binding Protein 5 | Growth Factor Binding Protein |
| | | *TFF3* | Trefoil Factor 3 | secretory proteins |
| | 4 | *TEAD1* | TEA Domain Transcription Factor 1 | ubiquitous transcriptional enhancer factor |
| | | *CH507-513H4.1* | Uncharacterized CH507-145C22.1 | RNA gene |
| | | *APOE* | Apolipoprotein E | major apoprotein of the chylomicron |
| | | *EMP3* | Epithelial Membrane Protein 3 | Protien involed in cell proliferation |

### FIGURE S5

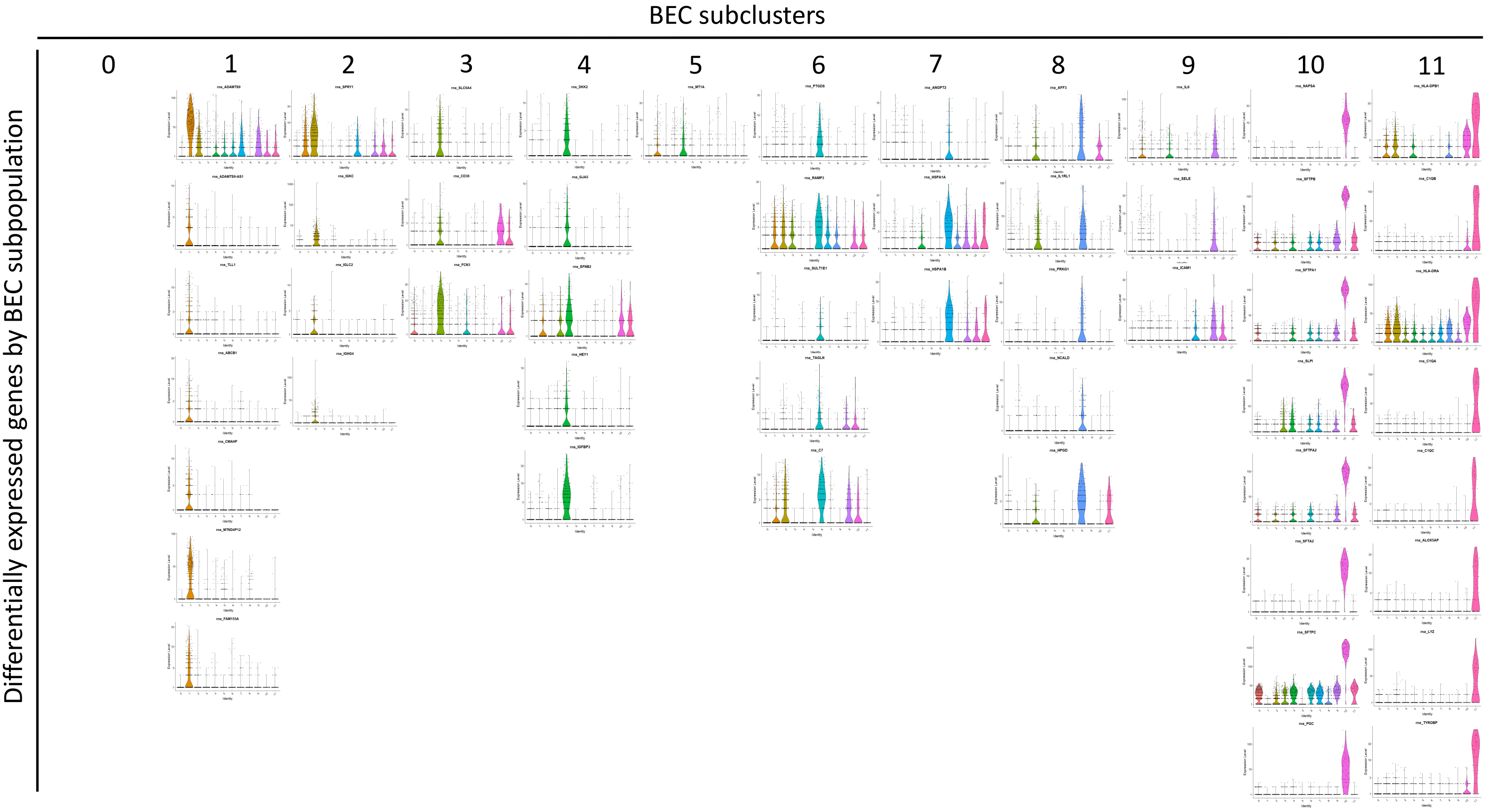
BEC subclusters
0
1
2
3
4
5
6
7
8
9
10
11
Differentially expressed genes by BEC subpopulation

### FIGURE S6

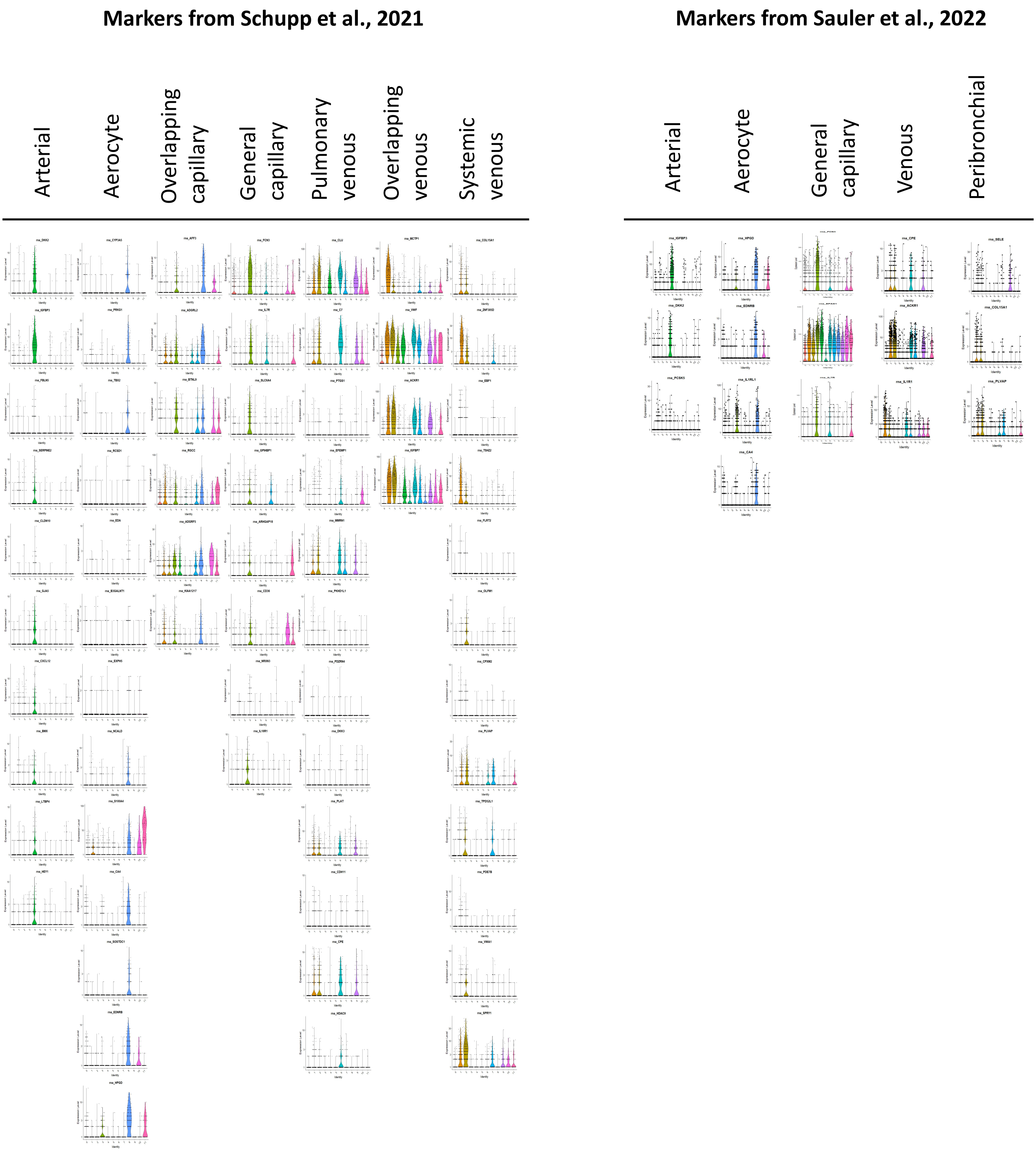
Markers from Schupp et al., 2021
Arterial
Aerocyte
Overlapping capillary
General capillary
Pulmonary venous
Overlapping venous
Systemic venous
Markers from Sauler et al., 2022
Arterial
Aerocyte
General capillary
Venous
Peribronchial

### FIGURE S7

**A**

Marker genes of EC subtypes proposed in six studies

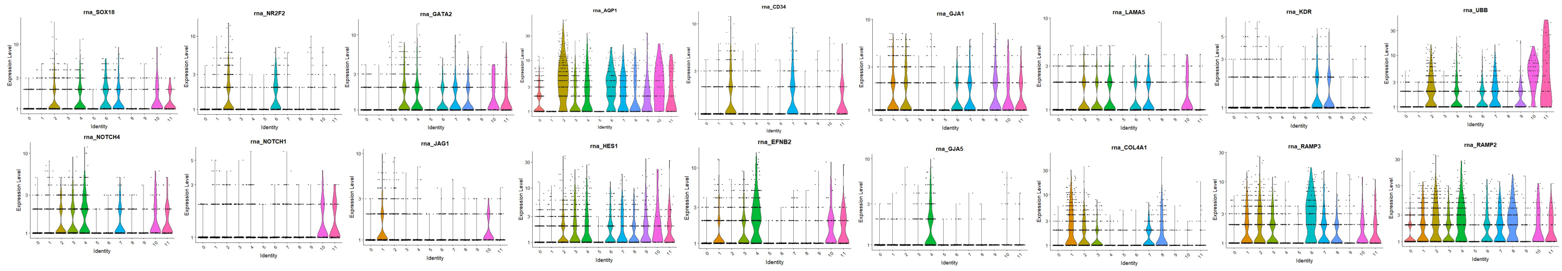


**B**

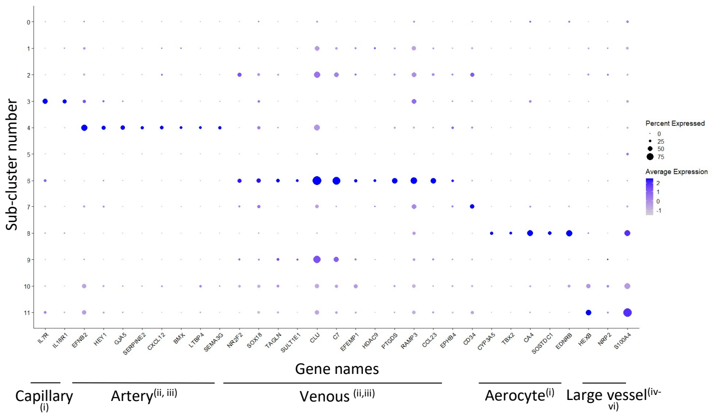

#### FIGURE S8

A

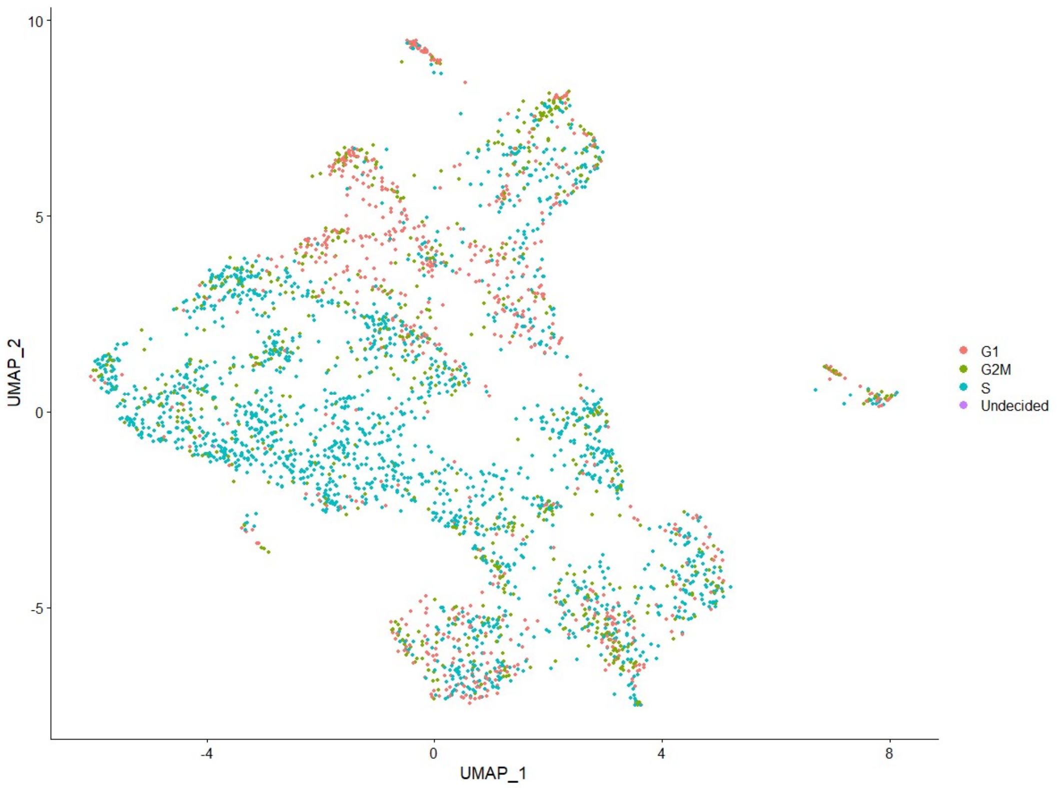


B

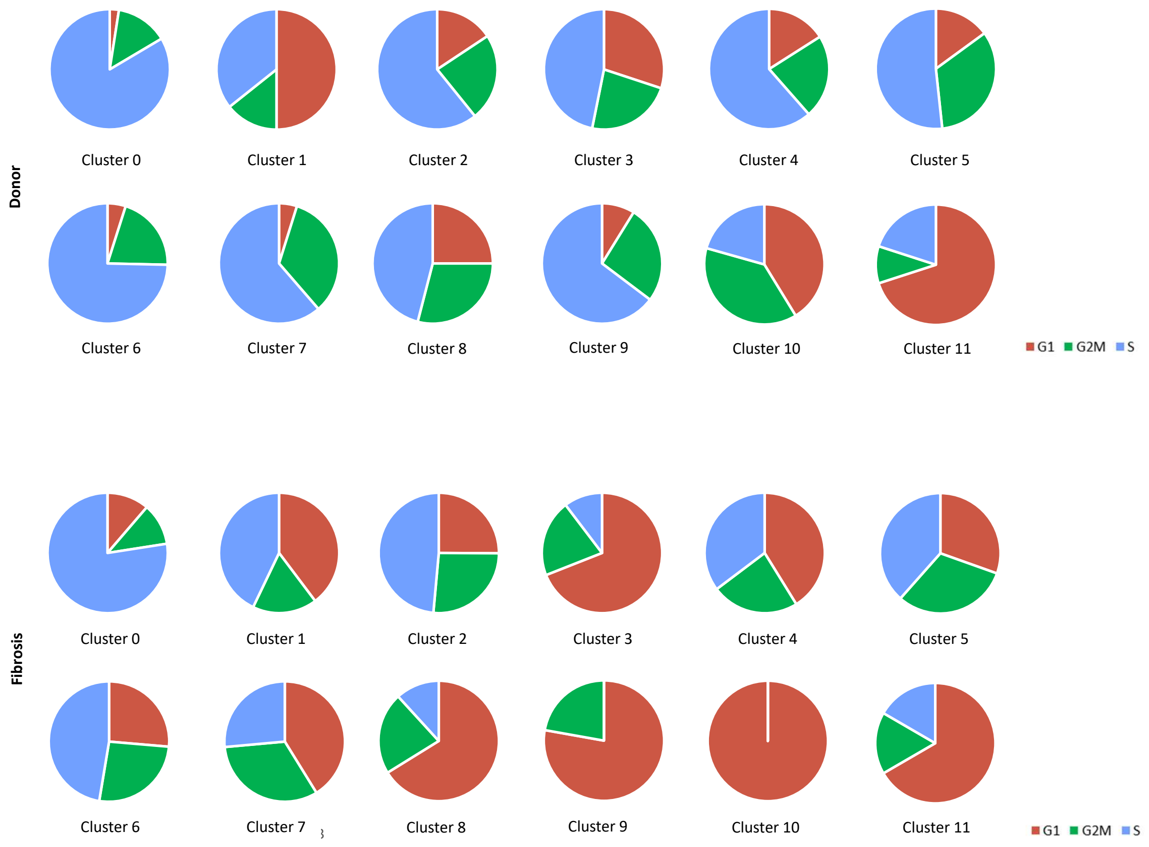


C

| BEC sub-population | Significance |
|---|---|
| 0 | <0.001 |
| 1 | ns |
| 2 | ns |
| 3 | <0.001 |
| 4 | <0.001 |
| 5 | 0.02 |
| 6 | 0.001 |
| 7 | <0.001 |
| 8 | <0.001 |
| 9 | <0.001 |
| 10 | ns |
| 11 | ns |

### FIGURE S9

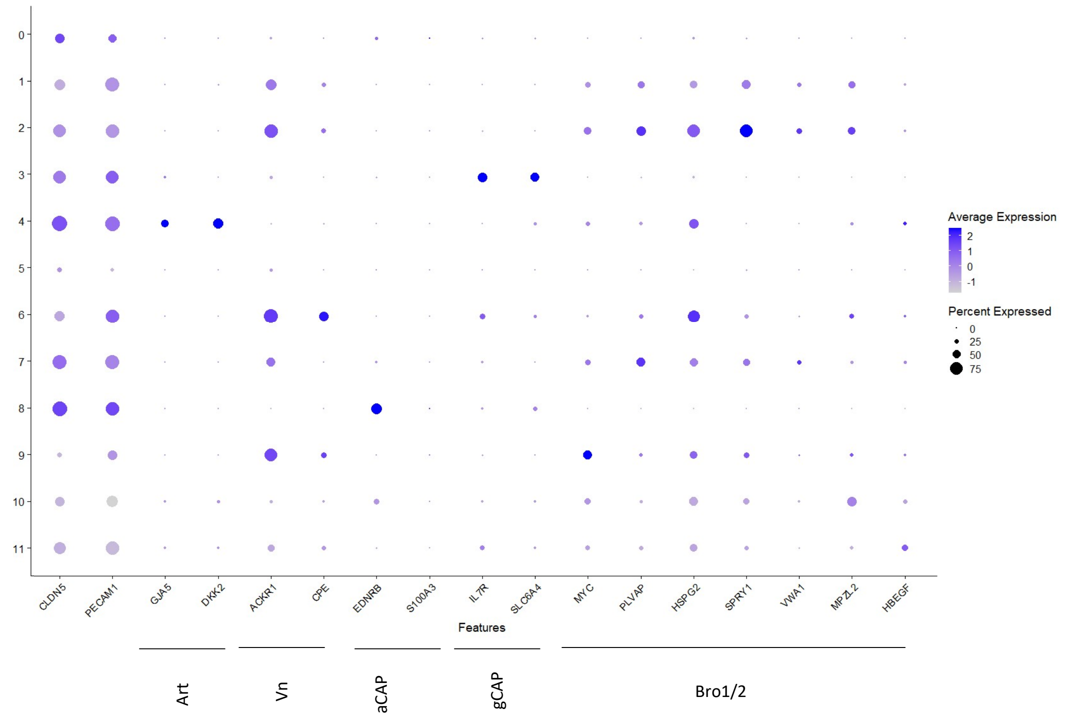

0
1
2
3
4
5
6
7
8
9
10
11
CLDN5
PECAM1
GJA5
DKK2
ACKR1
CPE
EDNRB
S100A3
IL7R
SLC6A4
MYC
PLVAP
HSPG2
SPRY1
VWA1
MPZL2
HBEGF
Features
Average Expression
2
1
0
-1
Percent Expressed
0
25
50
75
Art
Vn
aCAP
gCAP
Bro1/2

### FIGURE S10

A

**Markers from Rodor et al. 2021 and Bondareva et al., 2022**

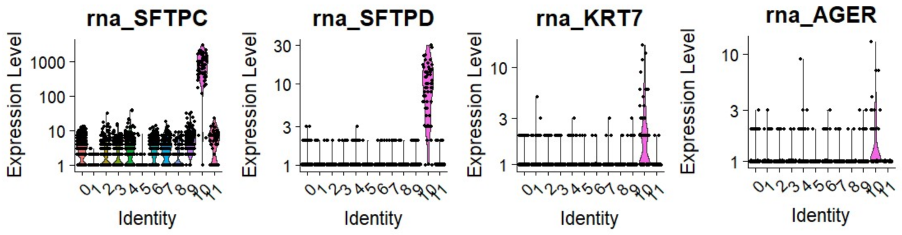


B

**Markers from Wang et al. 2022**

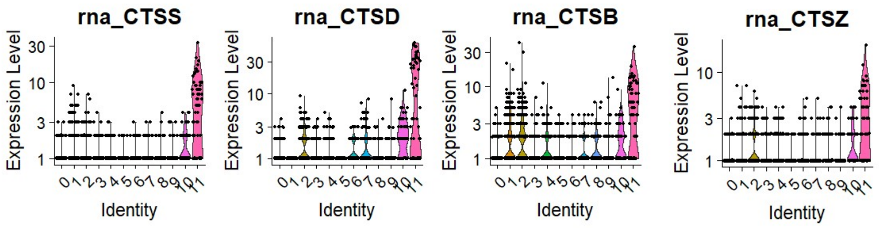

### FIGURE S11

**A**

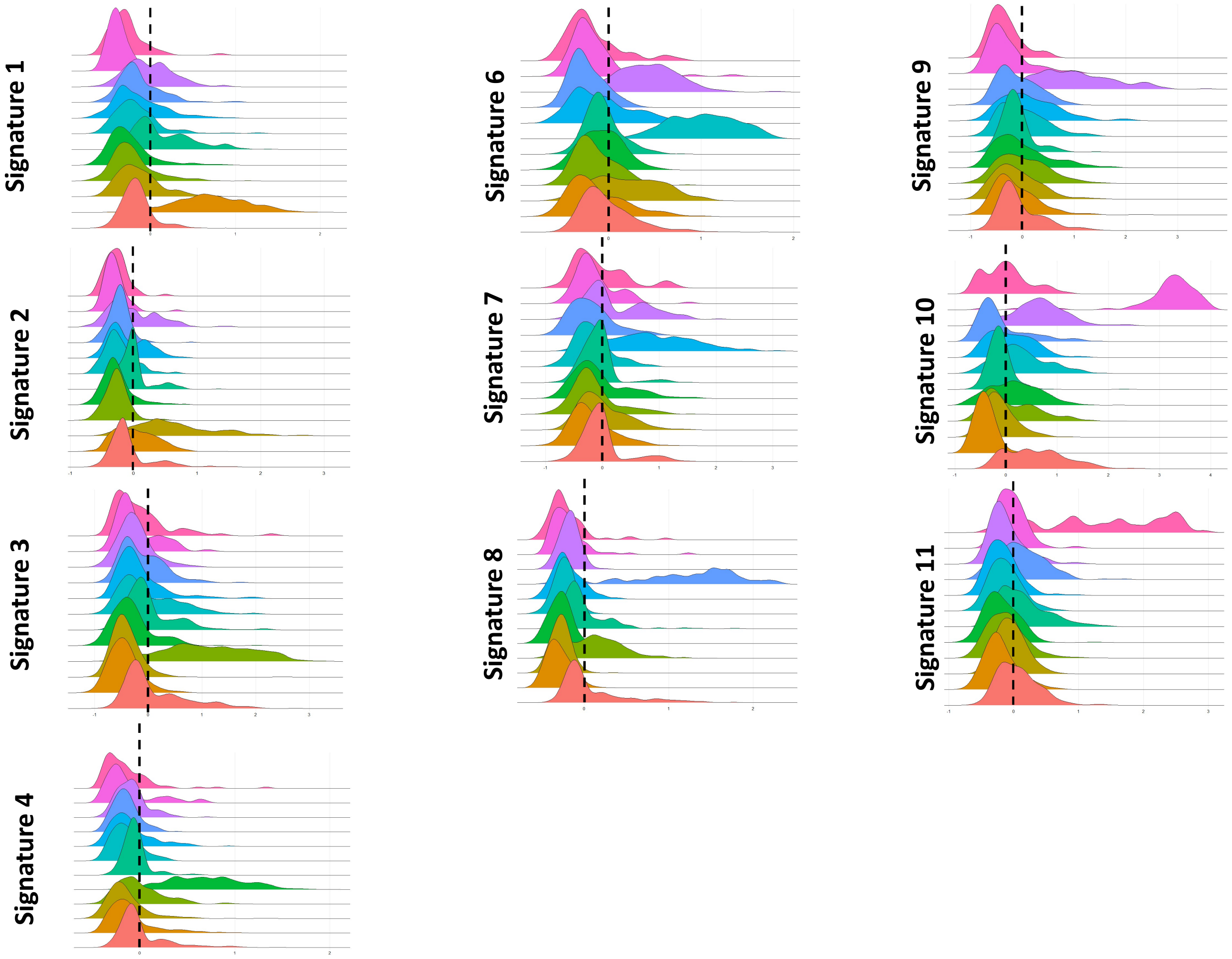


**B**

| BEC sub-cluster | Sig1 | Sig2 | Sig3 | Sig4 | Sig6 | Sig7 | Sig8 | Sig9 | Sig10 | Sig11 |
|---|---|---|---|---|---|---|---|---|---|---|
| 0 | -0.167 | -0.078 | 0.116 | 0.008 | -0.011 | -0.005 | 0.104 | -0.060 | 0.596 | 0.069 |
| 1 | 0.727 | 0.027 | -0.444 | -0.096 | -0.130 | -0.195 | -0.303 | -0.135 | -0.421 | -0.196 |
| 2 | -0.133 | 0.654 | -0.418 | -0.143 | 0.204 | -0.005 | -0.277 | -0.127 | -0.117 | -0.041 |
| 3 | -0.213 | -0.271 | 1.102 | -0.002 | -0.145 | -0.152 | 0.143 | 0.001 | 0.111 | -0.113 |
| 4 | -0.256 | -0.269 | -0.302 | 0.707 | -0.081 | -0.064 | -0.288 | -0.026 | 0.181 | -0.194 |
| 5 | 0.145 | -0.007 | 0.008 | -0.051 | -0.061 | -0.089 | -0.008 | -0.135 | -0.120 | 0.141 |
| 6 | -0.156 | -0.233 | -0.010 | -0.146 | 1.010 | -0.184 | -0.240 | -0.007 | 0.293 | -0.129 |
| 7 | -0.141 | -0.099 | -0.167 | -0.100 | -0.049 | 0.844 | -0.199 | 0.135 | 0.099 | -0.170 |
| 8 | -0.126 | -0.231 | -0.114 | -0.149 | -0.239 | -0.203 | 1.170 | -0.102 | -0.083 | 0.115 |
| 9 | -0.011 | -0.013 | -0.254 | -0.085 | 0.398 | 0.227 | -0.177 | 0.983 | 0.650 | -0.119 |
| 10 | -0.380 | -0.302 | -0.226 | -0.119 | -0.164 | -0.120 | -0.190 | -0.290 | 3.147 | -0.033 |
| 11 | -0.286 | -0.280 | -0.155 | -0.130 | -0.177 | -0.056 | -0.205 | -0.343 | -0.003 | 1.348 |

**C**

| | S1 | S2 | S3 | S4 | S6 | S7 | S8 | S9 | S10 | S11 |
|---|---|---|---|---|---|---|---|---|---|---|
| **S1** | 1 | 0.86 | -0.21 | -0.091 | 0.301 | -0.112 | -0.001 | 0.147 | -0.58 | -0.182 |
| **S2** | 0.86 | 1 | -0.322 | 0.035 | 0.517 | 0.196 | -0.182 | 0.098 | -0.58 | -0.182 |
| **S3** | -0.21 | -0.322 | 1 | 0.04 | -0.07 | -0.112 | 0.755 | 0.189 | 0.182 | 0.51 |
| **S4** | -0.091 | 0.035 | 0.04 | 1 | 0.112 | 0.322 | 0.063 | 0.266 | 0.203 | -0.231 |
| **S6** | 0.301 | 0.517 | -0.07 | 0.112 | 1 | 0.476 | -0.252 | 0.552 | 0.245 | -0.371 |
| **S7** | -0.112 | 0.196 | -0.112 | 0.322 | 0.476 | 1 | -0.049 | 0.343 | 0.294 | 0.007 |
| **S8** | -0.001 | -0.182 | 0.755 | 0.063 | -0.252 | -0.049 | 1 | 0.224 | 0.217 | 0.587 |
| **S9** | 0.147 | 0.098 | 0.189 | 0.266 | 0.552 | 0.343 | 0.224 | 1 | 0.385 | -0.453 |
| **S10** | -0.58 | -0.58 | 0.182 | 0.203 | 0.245 | 0.294 | 0.217 | 0.385 | 1 | -0.07 |
| **S11** | -0.182 | -0.182 | 0.51 | -0.231 | -0.371 | 0.007 | 0.587 | -0.453 | -0.07 | 1 |

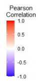

### FIGURE S12

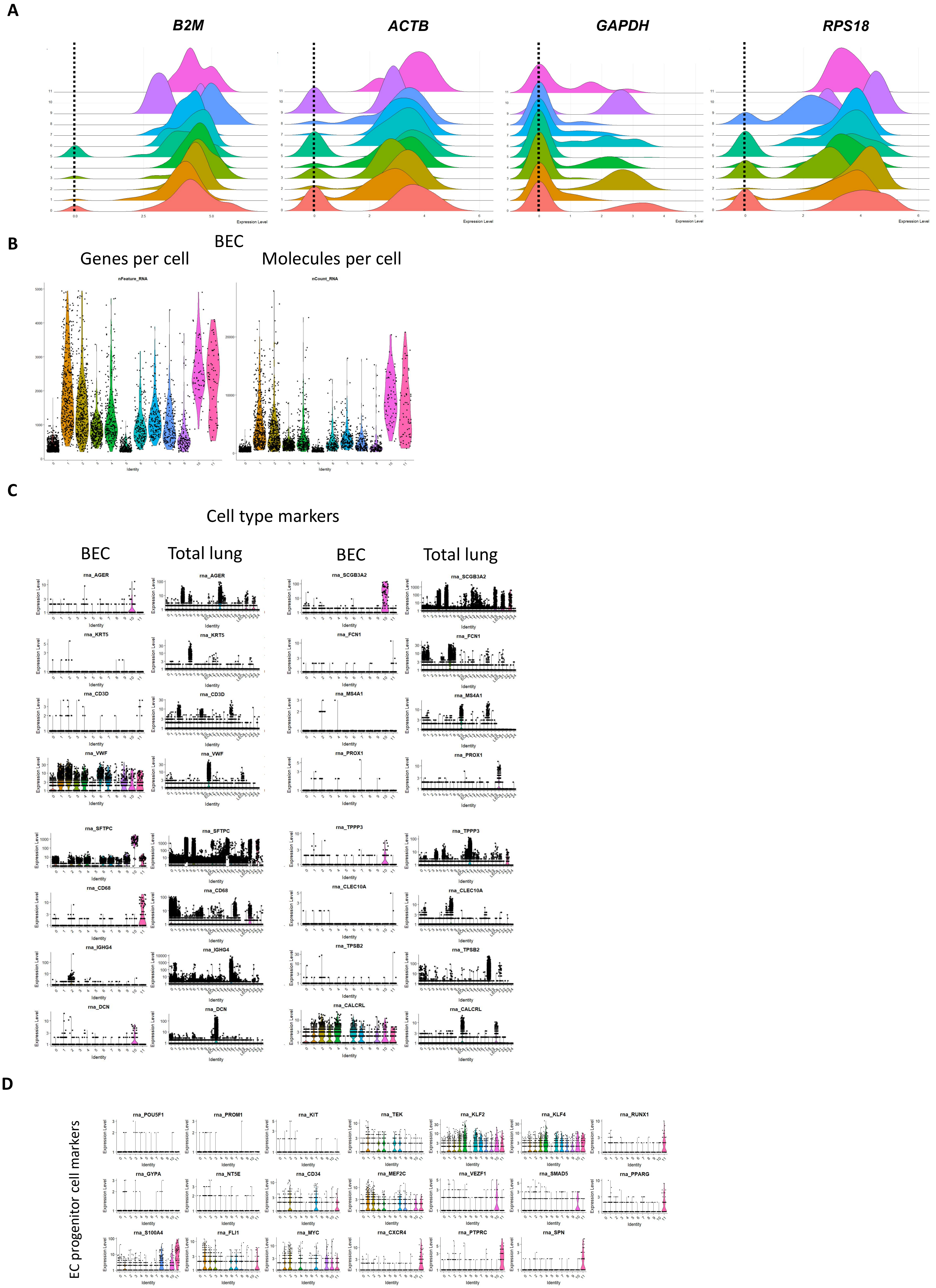
A
B2M
ACTB
GAPDH
RPS18
B
BEC
Genes per cell
Molecules per cell
nFeature_RNA
nCount_RNA
C
Cell type markers
BEC
Total lung
BEC
Total lung
rna_AGER
rna_SCGB3A2
rna_KRT5
rna_FCN1
rna_CD3D
rna_MS4A1
rna_VWF
rna_PROX1
rna_SFTPC
rna_TPPP3
rna_CD68
rna_CLEC10A
rna_IGHG4
rna_TPSB2
rna_DCN
rna_CALCRL
D
EC progenitor cell markers
rna_POU5F1
rna_PROM1
rna_KIT
rna_TEK
rna_KLF2
rna_KLF4
rna_RUNX1
rna_GYPA
rna_NT5E
rna_CD34
rna_MEF2C
rna_VEZF1
rna_SMAD5
rna_PPARG
rna_S100A4
rna_FLI1
rna_MYC
rna_CXCR4
rna_PTPRC
rna_SPN

### FIGURE S13

A

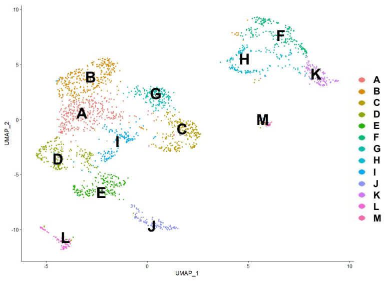


B

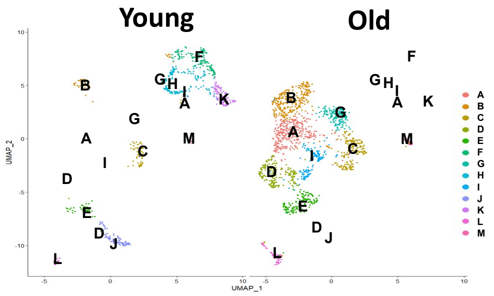


C

| BEC sub-population | Young (n) | Young (%) | Old (n) | Old (%) |
|---|---|---|---|---|
| A | 1 | 0.1 | 335 | 23.5 |
| B | 43 | 5.7 | 129 | 91 |
| C | 68 | 9.7 | 188 | 13.2 |
| D | 3 | 0.4 | 205 | 14.4 |
| E | 41 | 5.5 | 154 | 10.8 |
| F | 183 | 24.4 | 0 | 0.0 |
| G | 3 | 0.4 | 167 | 11.7 |
| H | 162 | 21.6 | 0 | 0.0 |
| I | 1 | 0.1 | 147 | 10.3 |
| J | 117 | 15.6 | 0 | 0.0 |
| K | 113 | 15.1 | 0 | 0.0 |
| L | 13 | 1.7 | 65 | 4.6 |
| M | 2 | 0.3 | 33 | 2.3 |

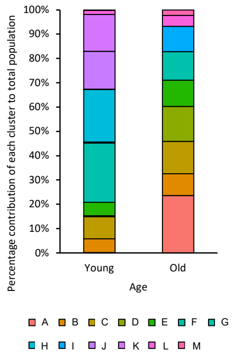


D

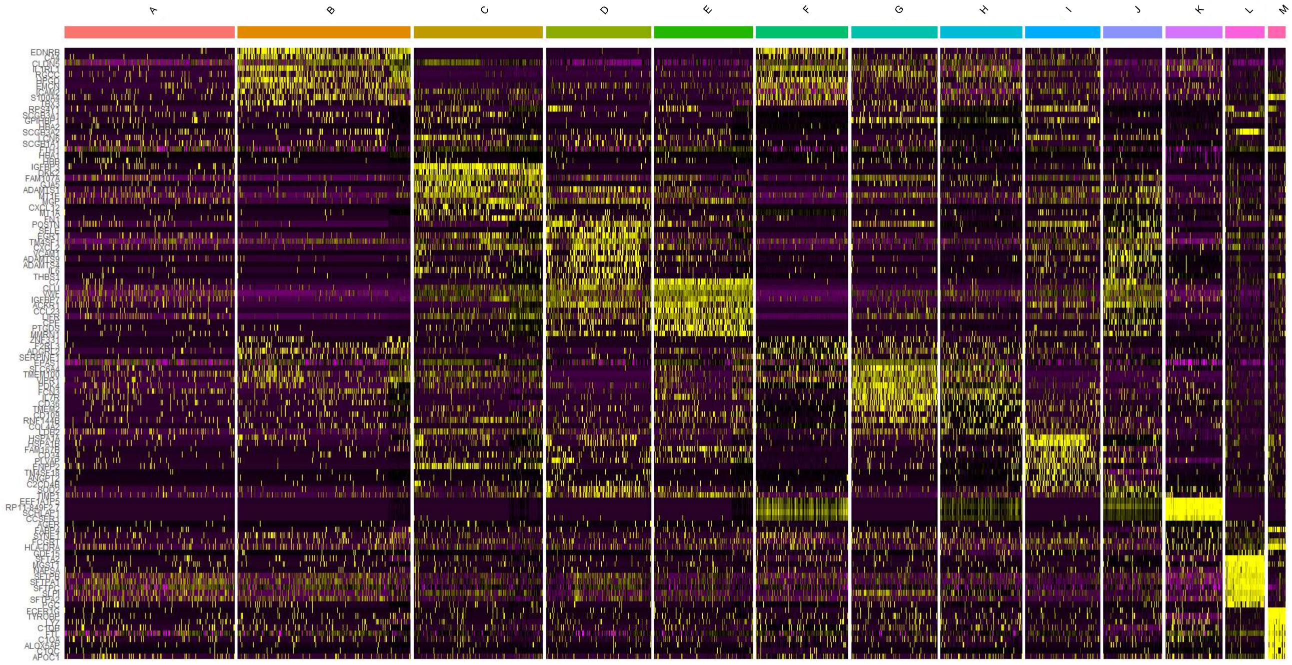

### FIGURE S14

A

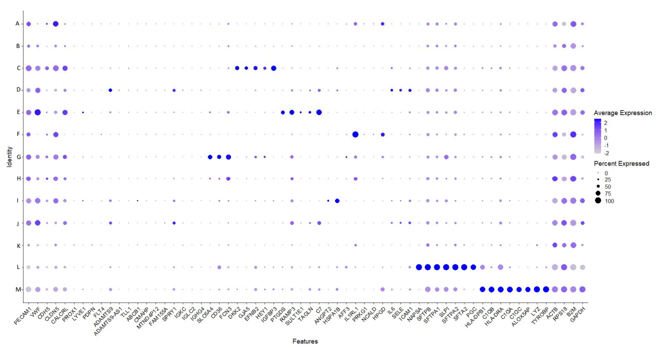


B

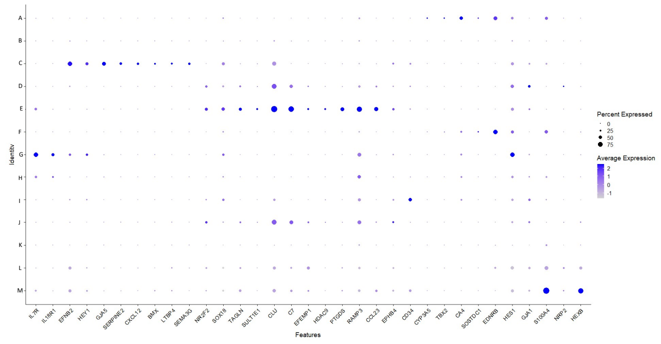


C

#### Young

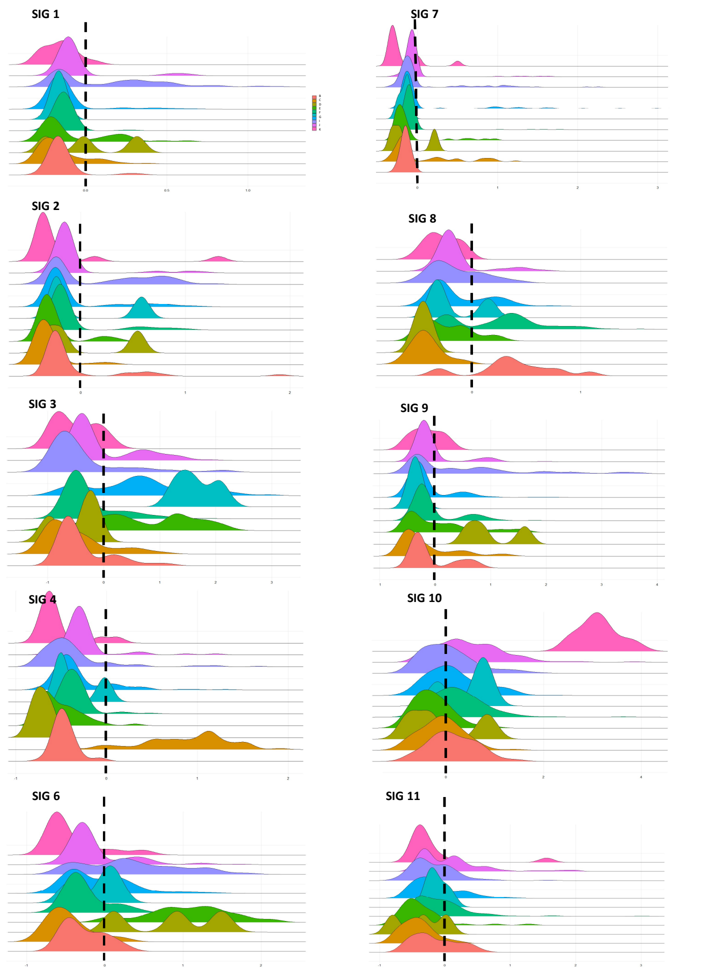


D

#### Old

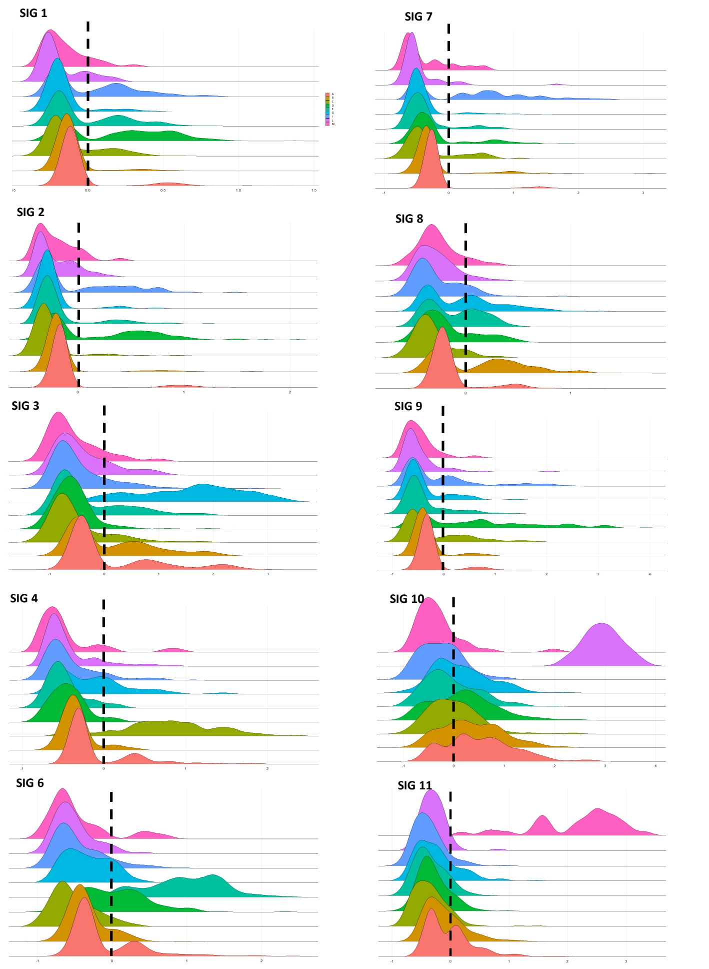


E

**Young**

| Sub-population | S1 | S2 | S3 | S4 | S6 | S7 | S8 | S9 | S10 | S11 |
|---|---|---|---|---|---|---|---|---|---|---|
| A | -0.092 | -0.149 | -0.475 | -0.326 | -0.302 | -0.065 | -0.217 | -0.227 | -0.393 | 0.071 |
| B | -0.148 | -0.07 | -0.382 | -0.466 | -0.266 | -0.151 | 0.375 | -0.075 | 0.145 | -0.153 |
| C | -0.151 | -0.315 | -0.511 | 0.873 | -0.469 | 0.036 | -0.408 | -0.168 | -0.083 | -0.424 |
| D | 0.019 | 0.008 | -0.481 | -0.715 | 0.842 | -0.112 | -0.454 | 1.010 | -0.078 | -0.371 |
| E | -0.060 | -0.253 | 0.661 | -0.507 | 1.061 | -0.102 | -0.237 | 0.079 | -0.275 | -0.355 |
| F | -0.123 | -0.084 | -0.288 | -0.359 | -0.254 | -0.081 | 0.226 | -0.039 | 0.237 | -0.067 |
| G | -0.167 | 0.037 | 1.650 | -0.343 | -0.105 | -0.164 | -0.167 | -0.361 | 0.449 | -0.121 |
| H | -0.118 | -0.070 | 0.661 | -0.290 | -0.087 | 0.079 | -0.117 | -0.149 | 0.172 | -0.139 |
| I | -0.212 | -0.283 | 0.536 | -0.166 | -0.656 | -0.162 | -0.506 | -0.521 | -0.156 | -0.424 |
| J | 0.128 | 0.157 | -0.384 | -0.367 | 0.343 | 0.081 | -0.150 | 0.496 | 0.041 | -0.177 |
| K | -0.057 | -0.077 | -0.055 | -0.233 | -0.111 | -0.020 | -0.139 | -0.058 | 0.487 | 0.101 |
| L | -0.158 | -0.182 | -0.505 | -0.535 | -0.428 | -0.199 | -0.278 | -0.148 | 3.120 | -0.195 |
| M | -0.133 | -0.326 | -1.021 | -0.269 | -0.209 | -0.231 | -0.348 | 0.089 | -0.201 | 2.351 |

**Old**

| Sub-population | S1 | S2 | S3 | S4 | S6 | S7 | S8 | S9 | S10 | S11 |
|---|---|---|---|---|---|---|---|---|---|---|
| A | -0.051 | -0.085 | 0.1 | -0.068 | -0.118 | -0.189 | -0.148 | -0.246 | 0.518 | -0.016 |
| B | -0.115 | -0.159 | 0.101 | -0.332 | -0.343 | -0.230 | 0.121 | -0.335 | 0.368 | -0.117 |
| C | -0.110 | -0.284 | -0.511 | 0.800 | -0.518 | -0.298 | -0.323 | -0.232 | 0.004 | -0.354 |
| D | 0.195 | 0.185 | -0.584 | -0.426 | 0.033 | -0.186 | -0.177 | 0.652 | 0.196 | -0.293 |
| E | -0.020 | -0.160 | -0.318 | -0.483 | 0.951 | -0.360 | -0.124 | -0.372 | -0.051 | -0.352 |
| F | 0.000 | 0.000 | 0.000 | 0.000 | 0.000 | 0.000 | 0.000 | 0.000 | 0.000 | 0.000 |
| G | -0.165 | -0.250 | 1.561 | -0.141 | -0.329 | -0.380 | -0.011 | -0.379 | 0.142 | -0.248 |
| H | 0.000 | 0.000 | 0.000 | 0.000 | 0.000 | 0.000 | 0.000 | 0.000 | 0.000 | 0.000 |
| I | 0.029 | -0.017 | -0.434 | -0.406 | -0.447 | 0.366 | -0.280 | -0.055 | -0.257 | -0.382 |
| J | 0.000 | 0.000 | 0.000 | 0.000 | 0.000 | 0.000 | 0.000 | 0.000 | 0.000 | 0.000 |
| K | 0.000 | 0.000 | 0.000 | 0.000 | 0.000 | 0.000 | 0.000 | 0.000 | 0.000 | 0.000 |
| L | -0.184 | -0.227 | -0.412 | -0.444 | -0.522 | -0.458 | -0.290 | -0.467 | 2.889 | -0.297 |
| M | -0.157 | -0.210 | -0.575 | -0.461 | -0.467 | -0.365 | -0.270 | -0.492 | -0.295 | 2.142 |

F

**Young**

| | S1 | S2 | S3 | S4 | S6 | S7 | S8 | S9 | S10 | S11 |
|---|---|---|---|---|---|---|---|---|---|---|
| S1 | 1 | 0.073 | 0.164 | -0.347 | 0.309 | 0.082 | -0.345 | 0.427 | -0.182 | -0.336 |
| S2 | 0.073 | 1 | 0.4 | 0.209 | 0.618 | 0.236 | 0.409 | 0.473 | 0.245 | 0.555 |
| S3 | 0.164 | 0.4 | 1 | 0.309 | 0.527 | 0.282 | 0.391 | 0.264 | -0.182 | 0.009 |
| S4 | -0.347 | 0.209 | 0.309 | 1 | 0.091 | 0.527 | 0.209 | -0.436 | -0.164 | 0.0455 |
| S6 | 0.309 | 0.618 | 0.527 | 0.091 | 1 | 0.164 | -0.1 | 0.545 | -0.191 | 0.055 |
| S7 | 0.082 | 0.236 | 0.282 | 0.527 | 0.164 | 1 | -0.155 | -0.309 | -0.527 | -0.434 |
| S8 | -0.345 | 0.409 | 0.391 | 0.209 | -0.1 | -0.155 | 1 | 0.173 | 0.4 | 0.564 |
| S9 | 0.427 | 0.473 | 0.264 | -0.436 | 0.545 | -0.309 | 0.173 | 1 | -0.118 | 0.282 |
| S10 | -0.182 | 0.245 | -0.182 | -0.164 | -0.191 | -0.527 | 0.4 | -0.118 | 1 | 0.673 |
| S11 | -0.336 | 0.555 | 0.009 | 0.0455 | 0.055 | -0.434 | 0.564 | 0.282 | 0.673 | 1 |

Pearson Correlation 1.0 0.5 0.0 -0.5 -1.0

**Old**

| | S1 | S2 | S3 | S4 | S6 | S7 | S8 | S9 | S10 | S11 |
|---|---|---|---|---|---|---|---|---|---|---|
| S1 | 1 | 0.273 | 0.152 | 0.079 | 0.358 | 0.539 | 0.236 | 0.297 | -0.333 | -0.43 |
| S2 | 0.273 | 1 | -0.356 | -0.6 | 0.345 | 0.418 | -0.188 | 0.079 | 0.079 | -0.115 |
| S3 | 0.152 | -0.356 | 1 | 0.358 | 0.382 | 0.03 | 0.855 | 0.382 | 0.164 | 0.297 |
| S4 | 0.079 | -0.6 | 0.358 | 1 | -0.667 | 0.042 | 0.358 | 0.248 | 0.2 | -0.042 |
| S6 | 0.358 | 0.345 | 0.382 | -0.667 | 1 | 0.127 | 0.394 | 0.418 | 0.115 | -0.164 |
| S7 | 0.539 | 0.418 | 0.03 | 0.042 | 0.127 | 1 | 0.418 | 0.552 | -0.382 | 0.176 |
| S8 | 0.236 | -0.188 | 0.855 | 0.358 | 0.394 | 0.418 | 1 | 0.636 | 0.139 | 0.37 |
| S9 | 0.297 | 0.079 | 0.382 | 0.248 | 0.418 | 0.552 | 0.636 | 1 | 0.079 | -0.188 |
| S10 | -0.333 | 0.079 | 0.164 | 0.2 | 0.115 | -0.382 | 0.139 | 0.079 | 1 | -0.067 |
| S11 | -0.43 | -0.115 | 0.297 | -0.042 | -0.164 | 0.176 | 0.37 | -0.188 | -0.067 | 1 |

Pearson Correlation 1.0 0.5 0.0 -0.5 -1.0

#### FIGURE S15

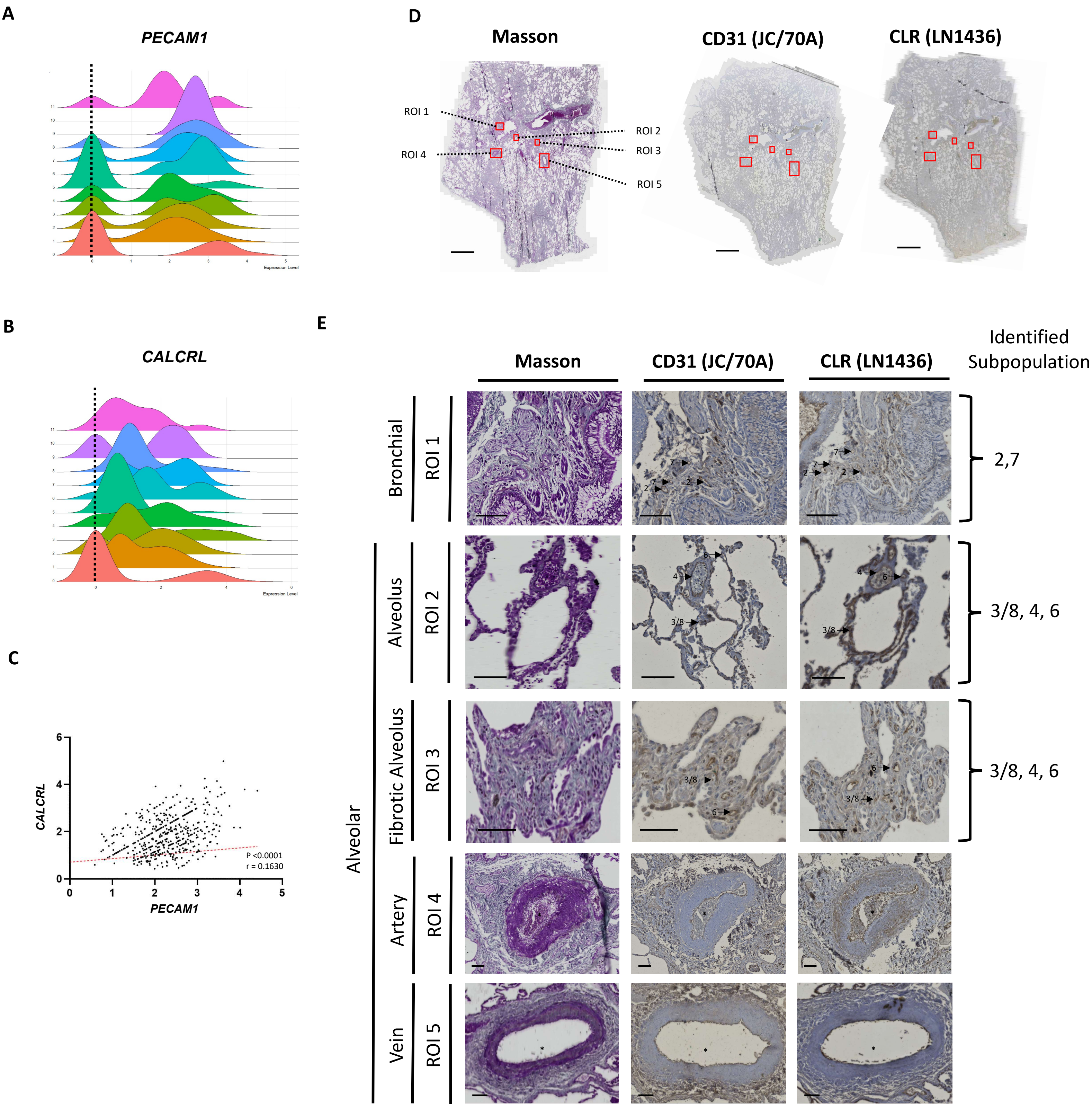
A
PECAM1
Expression Level
B
CALCRL
Expression Level
C
CALCRL
PECAM1
P <0.0001
r = 0.1630
D
Masson
CD31 (JC/70A)
CLR (LN1436)
ROI 1
ROI 2
ROI 3
ROI 4
ROI 5
E
Masson
CD31 (JC/70A)
CLR (LN1436)
Identified Subpopulation
Bronchial
ROI 1
2,7
Alveolar
Alveolus
ROI 2
3/8, 4, 6
Fibrotic Alveolus
ROI 3
3/8, 4, 6
Artery
ROI 4
Vein
ROI 5

#### FIGURE S16

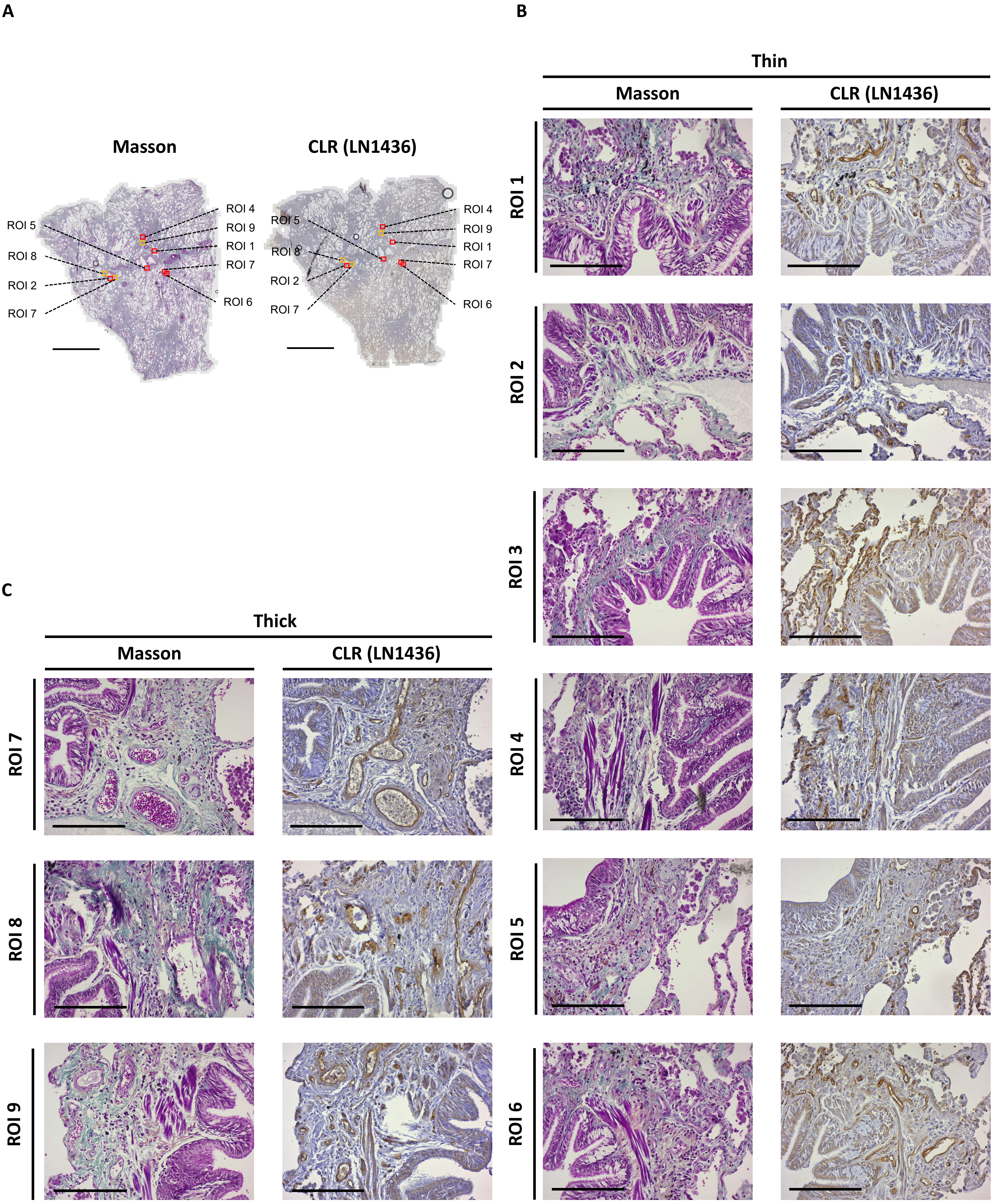
A
Masson
CLR (LN1436)
ROI 4
ROI 9
ROI 1
ROI 7
ROI 6
ROI 5
ROI 8
ROI 2
ROI 7
B
Thin
Masson
CLR (LN1436)
ROI 1
ROI 2
ROI 3
ROI 4
ROI 5
ROI 6
C
Thick
Masson
CLR (LN1436)
ROI 7
ROI 8
ROI 9

### FIGURE S17

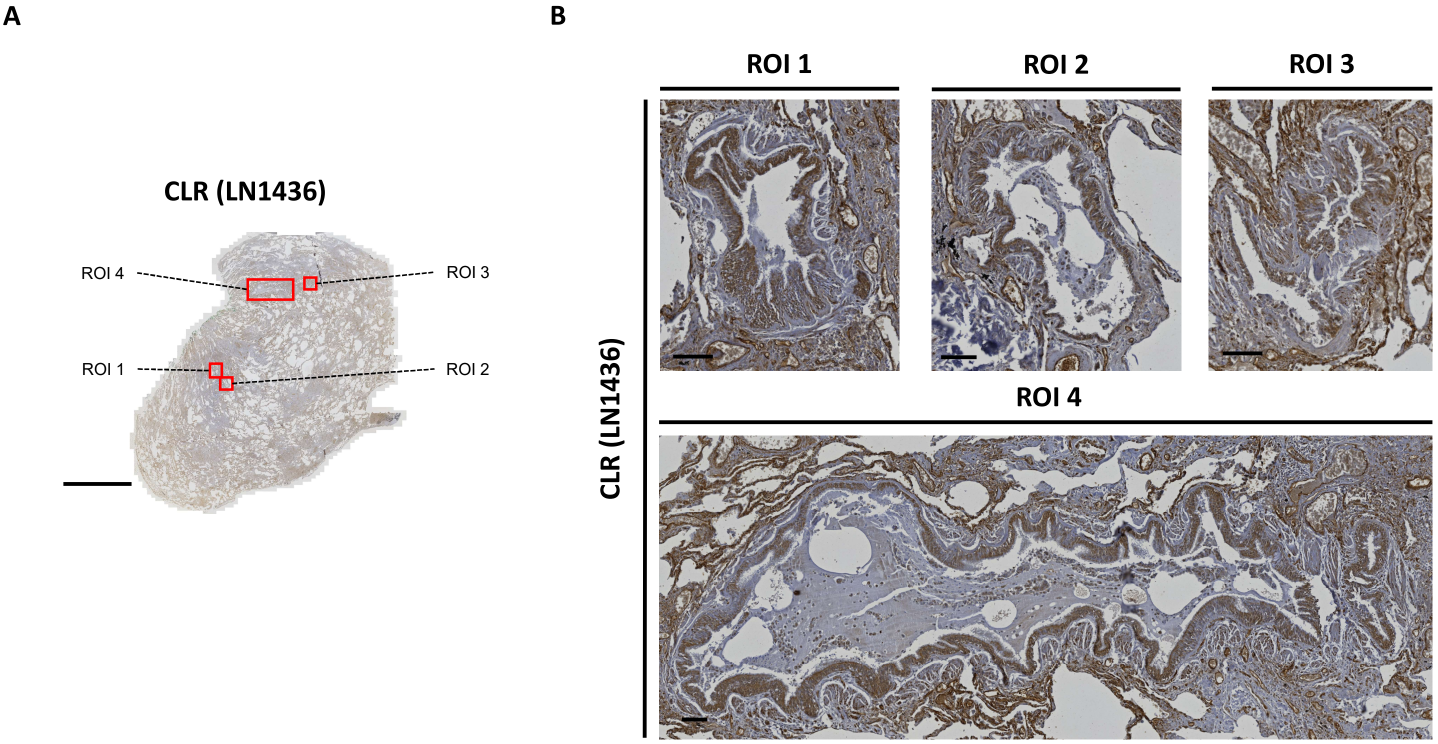
A
CLR (LN1436)
ROI 4
ROI 3
ROI 1
ROI 2
B
ROI 1
ROI 2
ROI 3
ROI 4
CLR (LN1436)

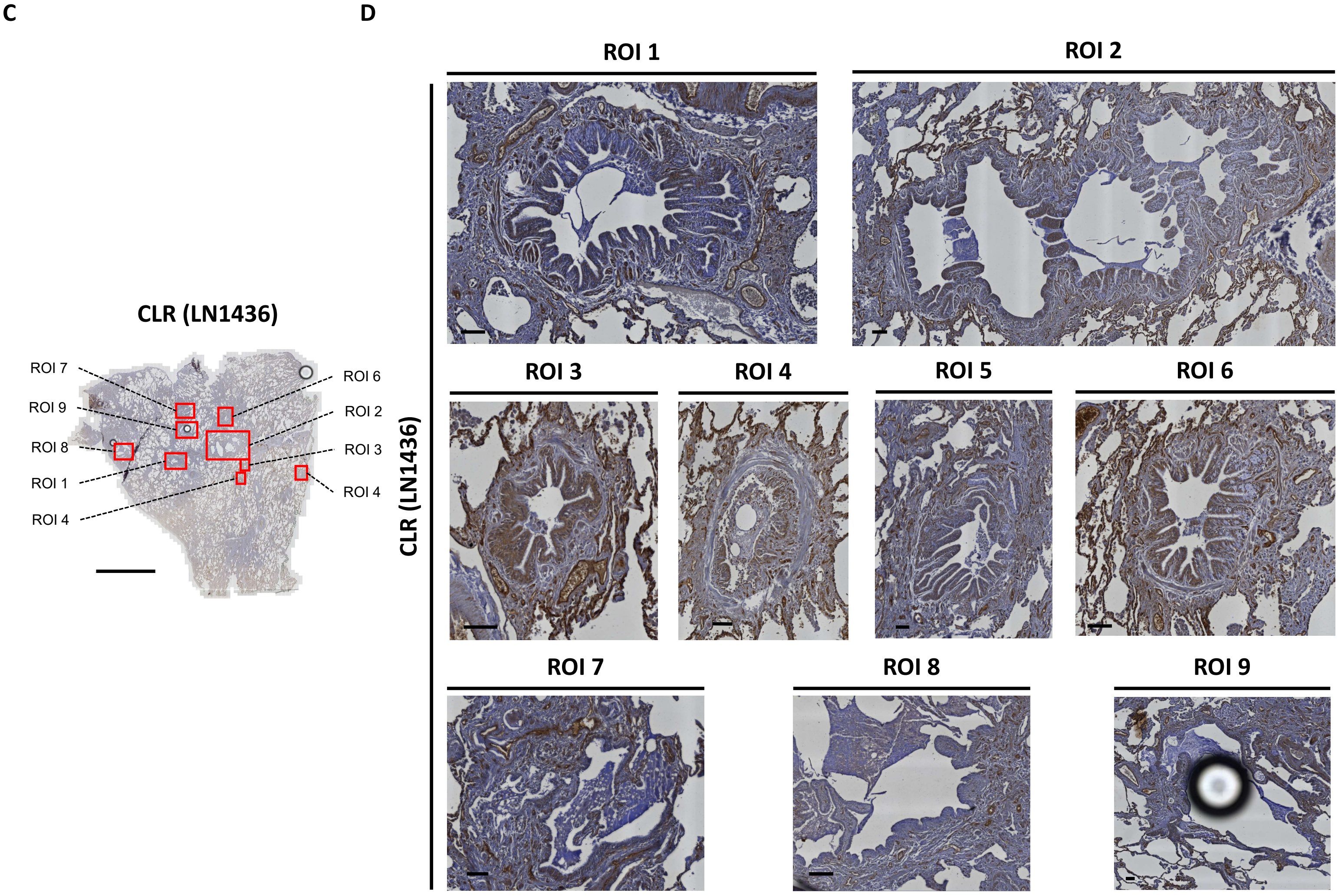
C
CLR (LN1436)
ROI 7
ROI 9
ROI 8
ROI 1
ROI 4
ROI 6
ROI 2
ROI 3
ROI 4
D
ROI 1
ROI 2
ROI 3
ROI 4
ROI 5
ROI 6
ROI 7
ROI 8
ROI 9
CLR (LN1436)

#### FIGURE S18

A

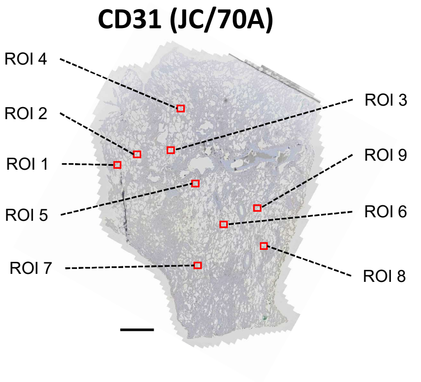


B

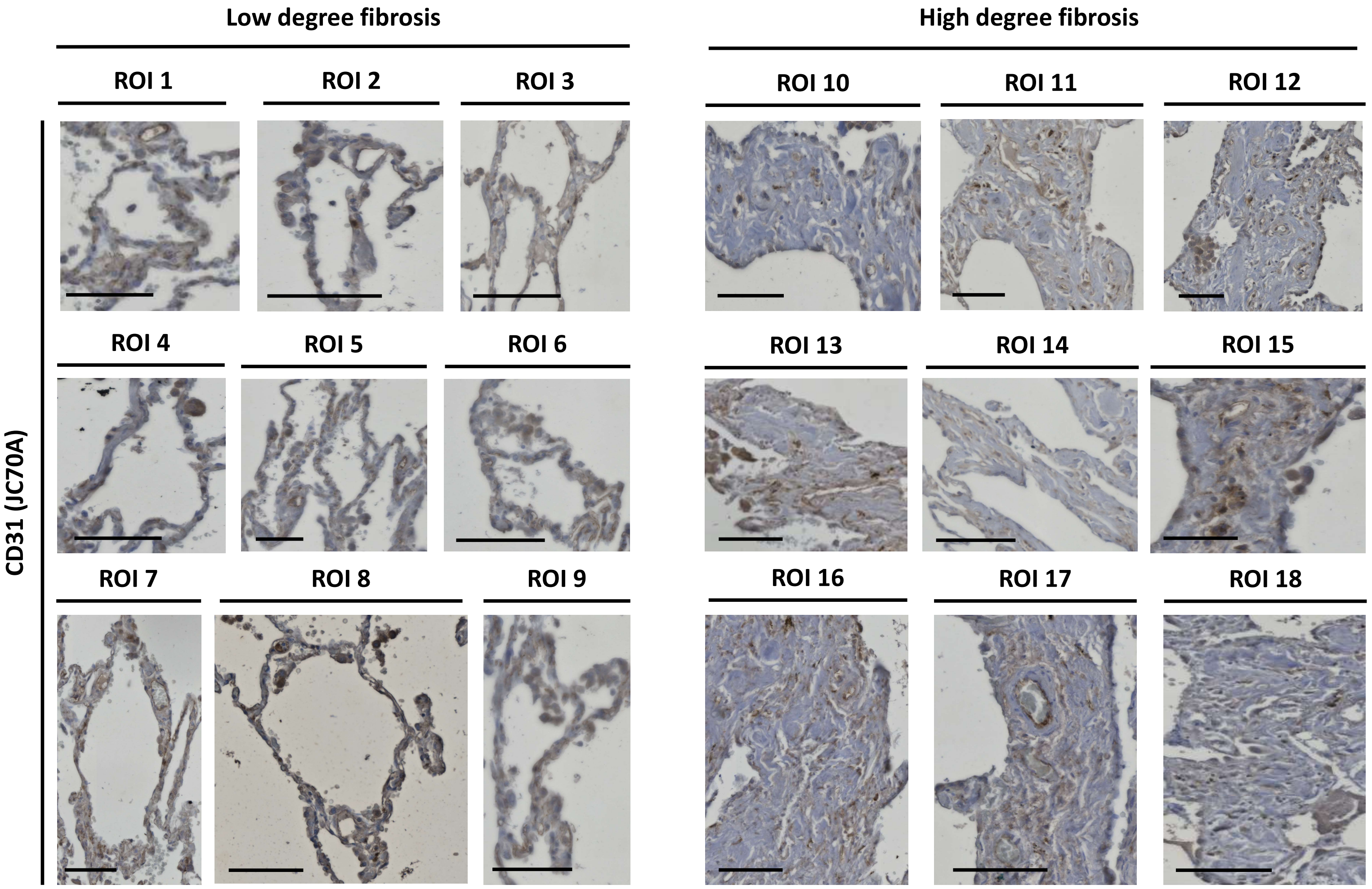

### FIGURE S19

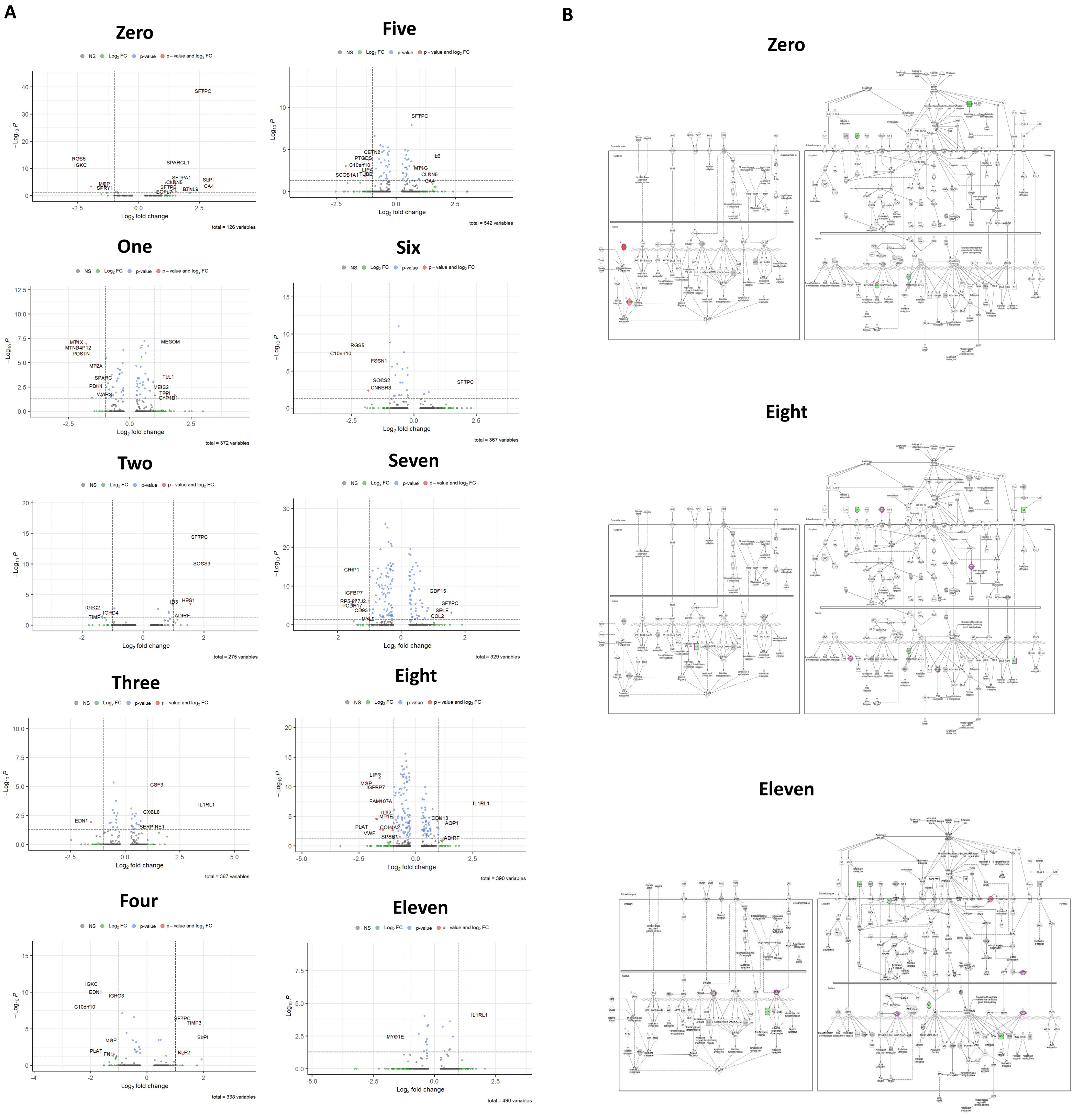
A
Zero
NS Log2 FC p-value p - value and log2 FC
SFTPC
RGS5
IGKC
SPARCL1
SFTPA1
CLDN5
SLPI
CA4
MGP
SPRY1
SFTPB
EGFL7
BTNL9
Log2 fold change
total = 126 variables
Five
SFTPC
CETN2
PTGDS
C10orf10
LIPA
TUBB
SCGB1A1
IL6
MT1G
CLDN5
CA4
total = 542 variables
One
MT1X
MTND4P12
POSTN
MT2A
SPARC
PDK4
WARS
MECOM
TLL1
MEIS2
TPPI
CYP1B1
total = 372 variables
Six
RGS5
C10orf10
FSCN1
SOCS2
CNKSR3
SFTPC
total = 367 variables
Two
SFTPC
SOCS3
IGLC2
IGHG4
TIMP1
ID3
HES1
ADIRF
total = 275 variables
Seven
CRIP1
IGFBP7
RP5-877J2.1
PCDH17
CD93
MYL9
GDF15
SFTPC
SBLE
COL2
total = 329 variables
Three
CSF3
IL1RL1
EDN1
CXCL8
SERPINE1
total = 367 variables
Eight
LIFR
MGP
IGFBP7
FAM107A
IL32
MT1E
PLAT
COL4A2
VWF
SPSB1
IL1RL1
CDH13
AQP1
ADIRF
total = 390 variables
Four
IGKC
EDN1
IGHG3
C10orf10
SFTPC
TIMP3
SLPI
MGP
PLAT
FN1
KLF2
total = 338 variables
Eleven
IL1RL1
MYO1E
total = 490 variables
B
Zero
Eight
Eleven

### FIGURE S20

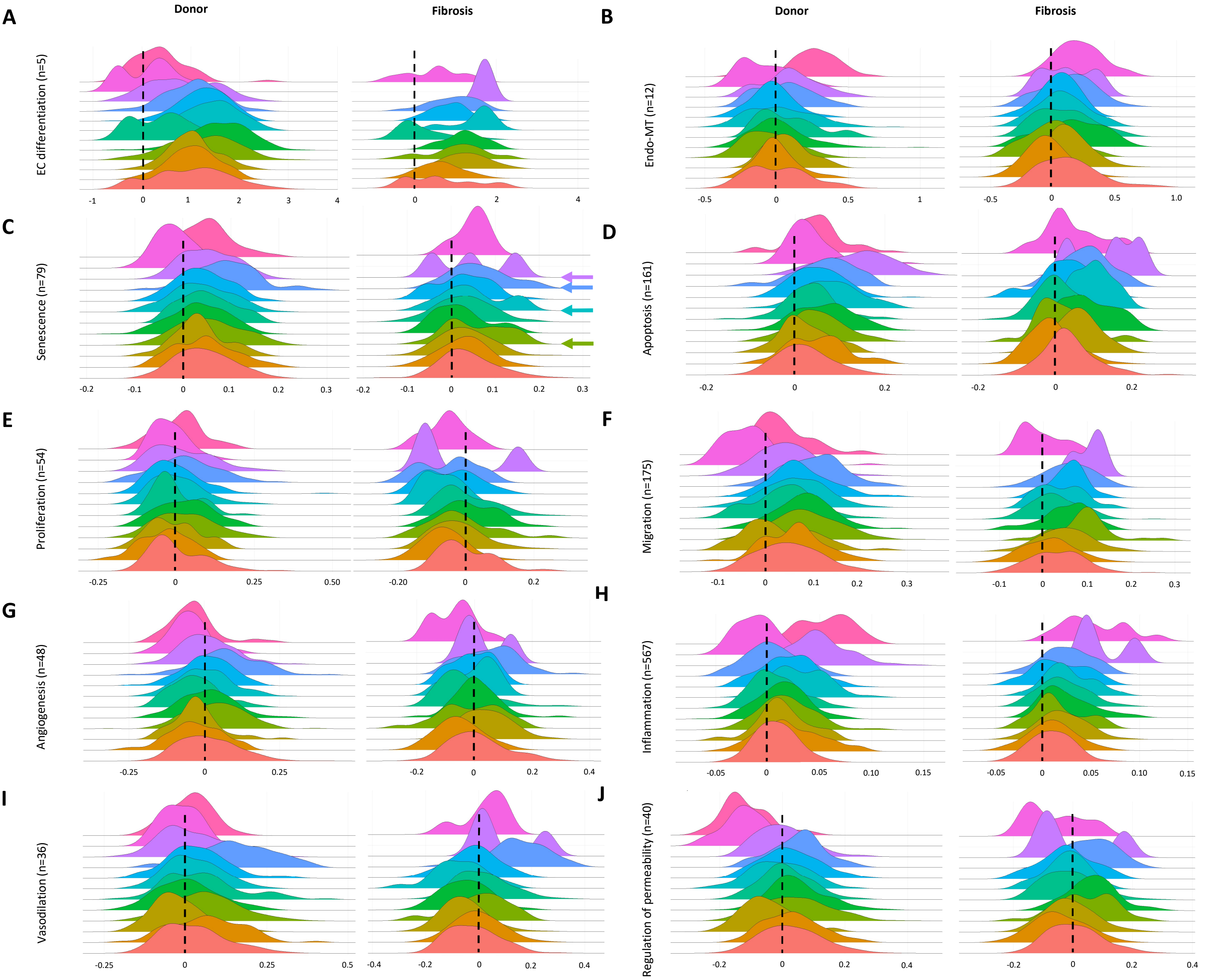


K

| BEC sub-population | | Endothelial cell-relevant processes | | | | | | | | | |
|---|---|---|---|---|---|---|---|---|---|---|---|
| Key | Number | EC Differentiation | Endo MT | Senescence | Apoptosis | Proliferation | Migration | Angiogenesis | Inflammation | Vasodilation | Permeability |
| | 0 | 1.011 | 0.014 | 0.044 | 0.004 | 0.014 | 0.020 | 0.049 | 0.008 | 0.027 | 0.029 |
| | 1 | 1.051 | -0.006 | 0.045 | -0.034 | -0.031 | 0.049 | 0.061 | 0.021 | 0.068 | 0.032 |
| | 2 | 1.033 | 0.072 | 0.035 | -0.010 | -0.023 | 0.021 | 0.030 | 0.014 | -0.022 | -0.050 |
| | 3 | 1.222 | -0.054 | 0.055 | 0.028 | 0.040 | 0.049 | 0.086 | 0.018 | 0.063 | 0.039 |
| | 4 | 1.603 | 0.009 | 0.034 | 0.028 | 0.012 | 0.066 | 0.081 | 0.014 | 0.007 | 0.049 |
| | 5 | 0.443 | 0.076 | 0.050 | -0.004 | -0.031 | 0.034 | 0.038 | 0.007 | 0.053 | 0.011 |
| | 6 | 1.430 | 0.033 | 0.039 | 0.002 | 0.003 | 0.082 | 0.062 | 0.031 | 0.025 | -0.011 |
| | 7 | 1.261 | -0.036 | 0.041 | -0.006 | 0.007 | 0.050 | 0.075 | 0.020 | 0.035 | 0.023 |
| | 8 | 1.029 | 0.054 | 0.090 | 0.029 | 0.075 | 0.066 | 0.101 | 0.005 | 0.162 | 0.049 |
| | 9 | 0.836 | 0.094 | 0.055 | 0.016 | 0.010 | 0.138 | 0.042 | 0.048 | 0.021 | 0.002 |
| | 10 | 0.217 | -0.087 | -0.017 | -0.022 | -0.052 | 0.033 | -0.038 | -0.010 | -0.019 | -0.117 |
| | 11 | 0.374 | 0.224 | 0.050 | 0.012 | -0.037 | 0.047 | 0.036 | 0.049 | 0.014 | -0.116 |

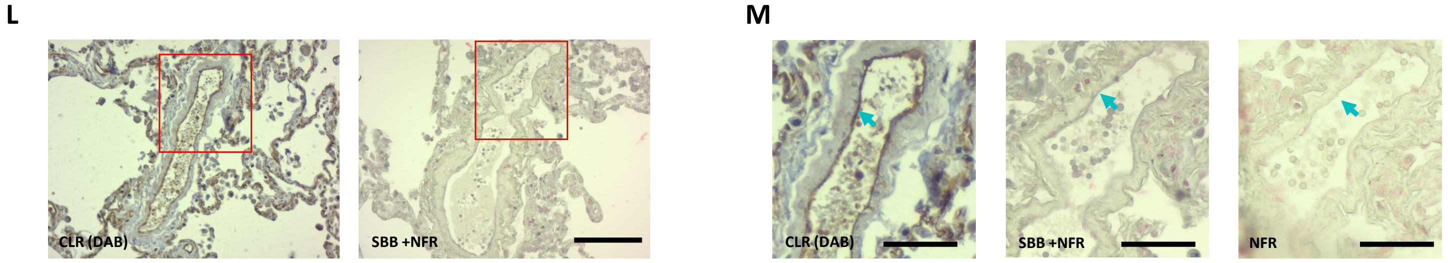

### FIGURE S21

**A**

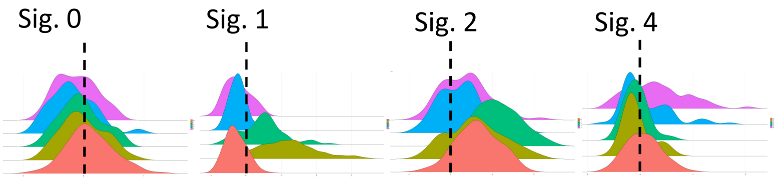


**B**

| Subpopulation | Sig 0 | Sig 1 | Sig 2 | Sig 4 |
|---|---|---|---|---|
| **0** | 0.106 | -0.290 | 0.640 | 0.061 |
| **1** | -0.009 | 1.162 | 0.604 | -0.068 |
| **2** | -0.045 | 0.499 | 1.102 | -0.079 |
| **3** | -0.187 | -0.185 | 0.249 | 0.087 |
| **4** | -0.124 | -0.239 | 0.362 | 0.323 |

**C**

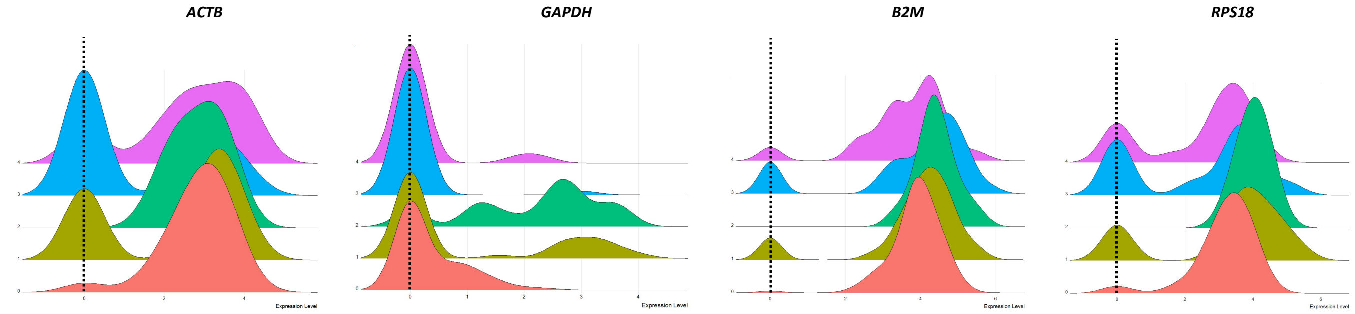


**D**

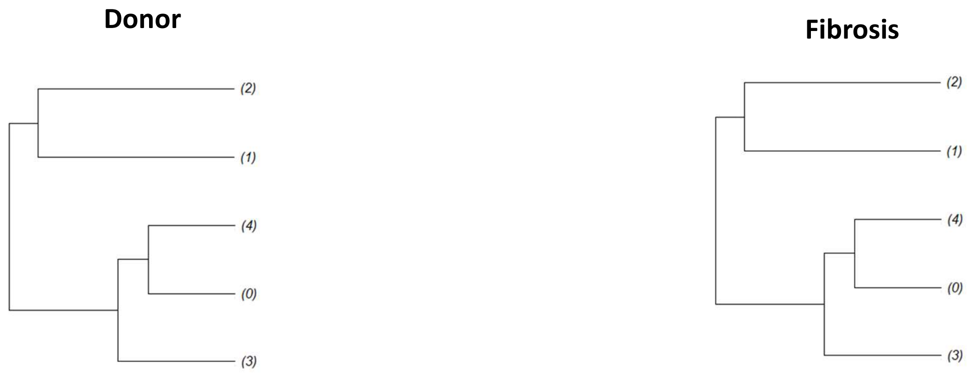


**E**

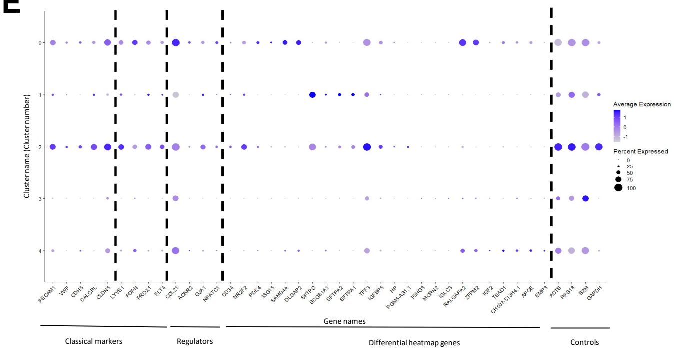


**F**

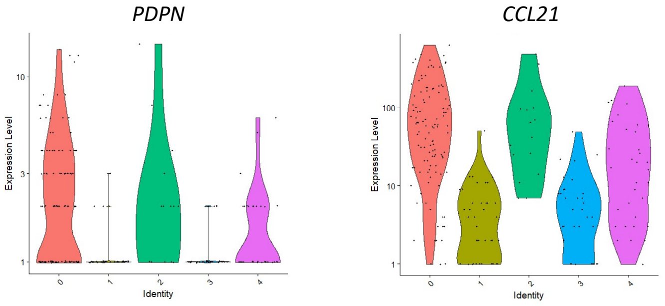

### FIGURE S22

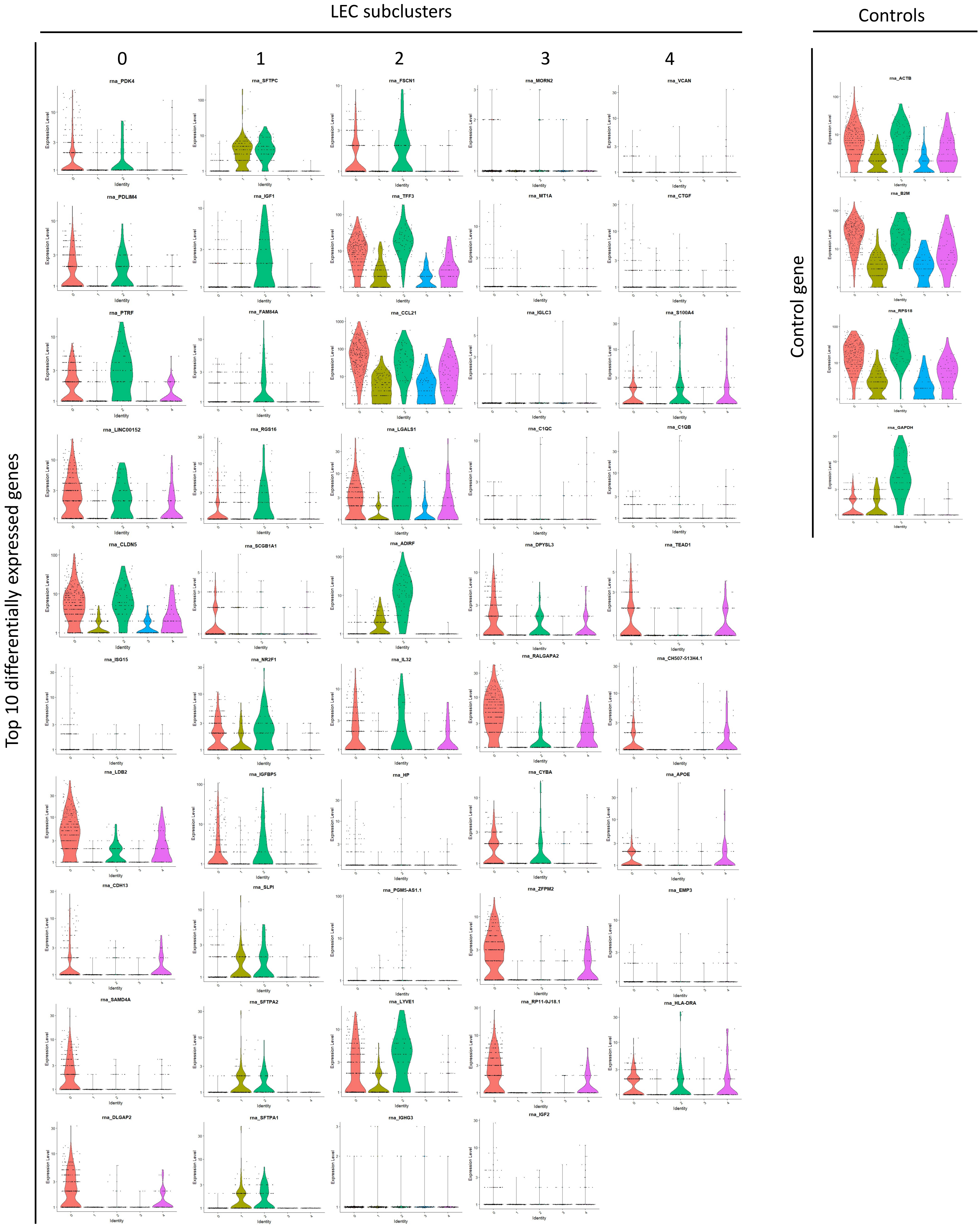
LEC subclusters
Controls
0
1
2
3
4
Top 10 differentially expressed genes
Control gene
rna_PDK4
rna_SFTPC
rna_FSCN1
rna_MORN2
rna_VCAN
rna_PDLIM4
rna_IGF1
rna_TFF3
rna_MT1A
rna_CTGF
rna_PTRF
rna_FAM84A
rna_CCL21
rna_IGLC3
rna_S100A4
rna_LINC00152
rna_RGS16
rna_LGALS1
rna_C1QC
rna_C1QB
rna_CLDN5
rna_SCGB1A1
rna_ADIRF
rna_DPYSL3
rna_TEAD1
rna_ISG15
rna_NR2F1
rna_IL32
rna_RALGAPA2
rna_CH507-513H4.1
rna_LDB2
rna_IGFBP5
rna_HP
rna_CYBA
rna_APOE
rna_CDH13
rna_SLPI
rna_PGM5-AS1.1
rna_ZFPM2
rna_EMP3
rna_SAMD4A
rna_SFTPA2
rna_LYVE1
rna_RP11-9J18.1
rna_HLA-DRA
rna_DLGAP2
rna_SFTPA1
rna_IGHG3
rna_IGF2
rna_ACTB
rna_B2M
rna_RPS18
rna_GAPDH
Expression Level
Identity

### FIGURE S23

**A**

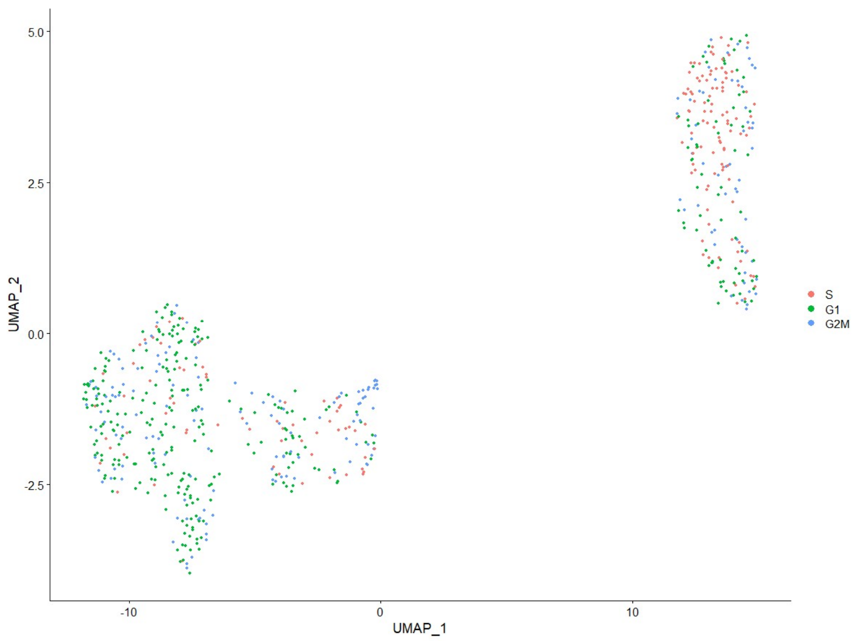


**B**

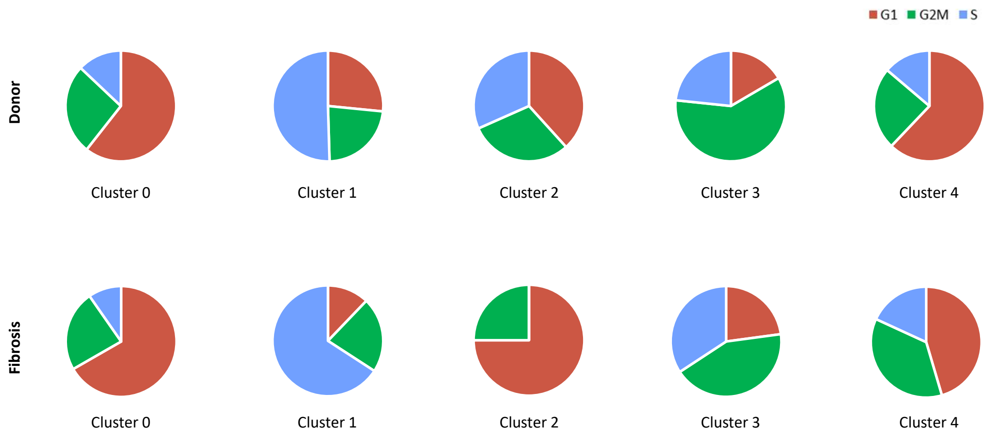


**C**

| LEC sub-population | Significance |
|---|---|
| 0 | ns |
| 1 | ns |
| 2 | 0.0119 |
| 3 | ns |
| 4 | ns |

### FIGURE S24

**A**

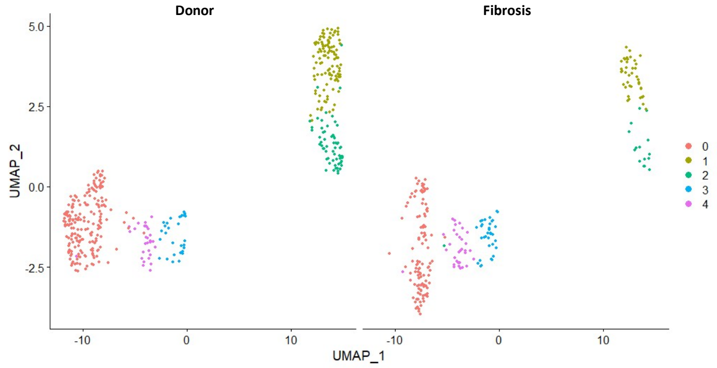


**B**

#### Cell number

| LEC Sub-population | Donor, n (%) | Fibrosis, n (%) | P-value |
|---|---|---|---|
| 0 | 192 (45.7) | 114 (47.7) | 0.671 |
| 1 | 109 (25.9) | 41 (17.2) | 0.878 |
| 2 | 60 (14.3) | 16 (6.69) | 0.837 |
| 3 | 30 (7.1) | 35 (14.6) | 0.291 |
| 4 | 29 (6.9) | 33 (13.8) | 0.264 |

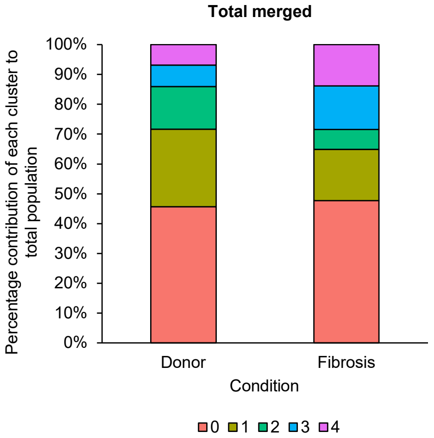


**C**

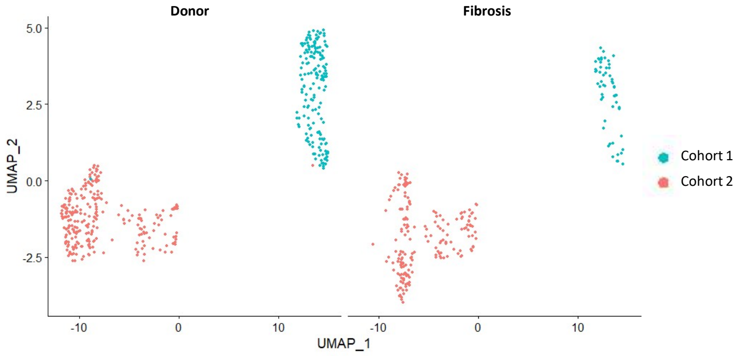


**D**

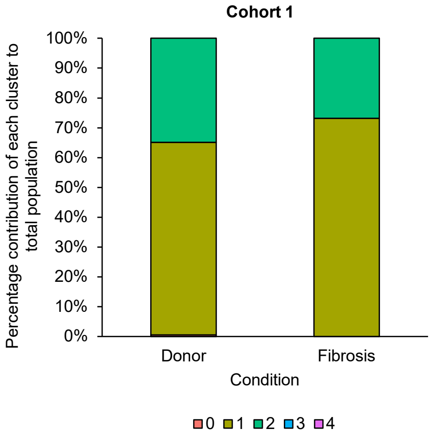


**E**

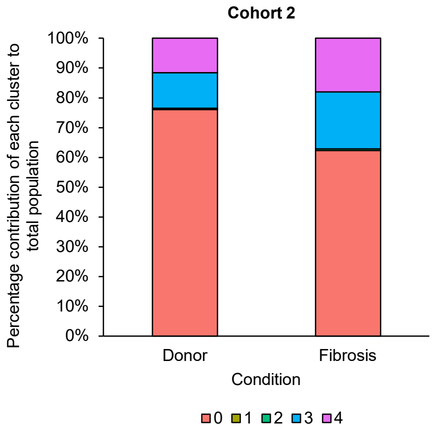

### FIGURE S25

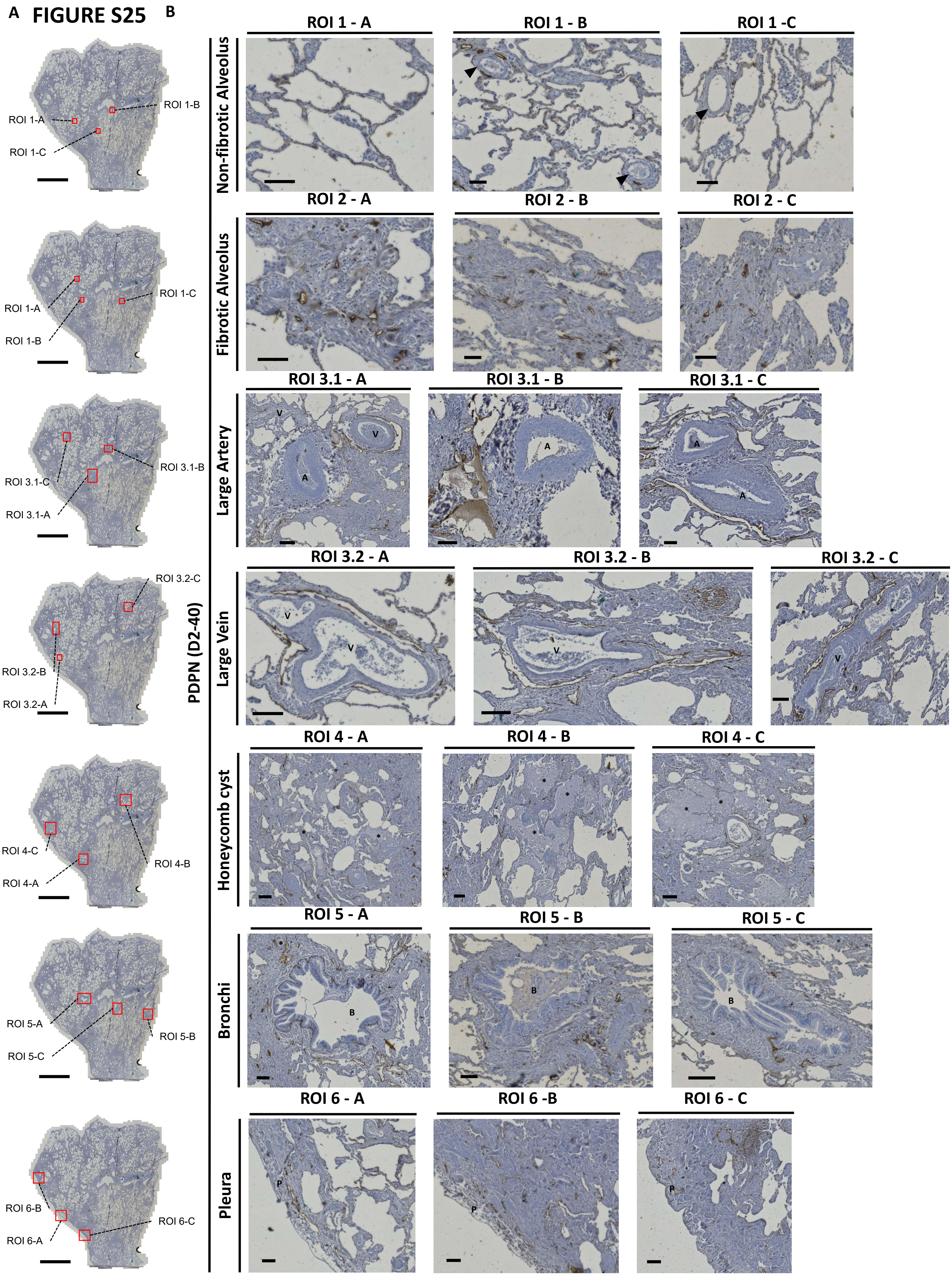
A
B
ROI 1-A
ROI 1-B
ROI 1-C
ROI 3.1-B
ROI 3.1-C
ROI 3.1-A
ROI 3.2-C
ROI 3.2-B
ROI 3.2-A
ROI 4-C
ROI 4-A
ROI 4-B
ROI 5-A
ROI 5-B
ROI 5-C
ROI 6-B
ROI 6-A
ROI 6-C
PDPN (D2-40)
Non-fibrotic Alveolus
ROI 1 - A
ROI 1 - B
ROI 1 -C
Fibrotic Alveolus
ROI 2 - A
ROI 2 - B
ROI 2 - C
Large Artery
ROI 3.1 - A
ROI 3.1 - B
ROI 3.1 - C
Large Vein
ROI 3.2 - A
ROI 3.2 - B
ROI 3.2 - C
Honeycomb cyst
ROI 4 - A
ROI 4 - B
ROI 4 - C
Bronchi
ROI 5 - A
ROI 5 - B
ROI 5 - C
Pleura
ROI 6 - A
ROI 6 -B
ROI 6 - C

### FIGURE S26

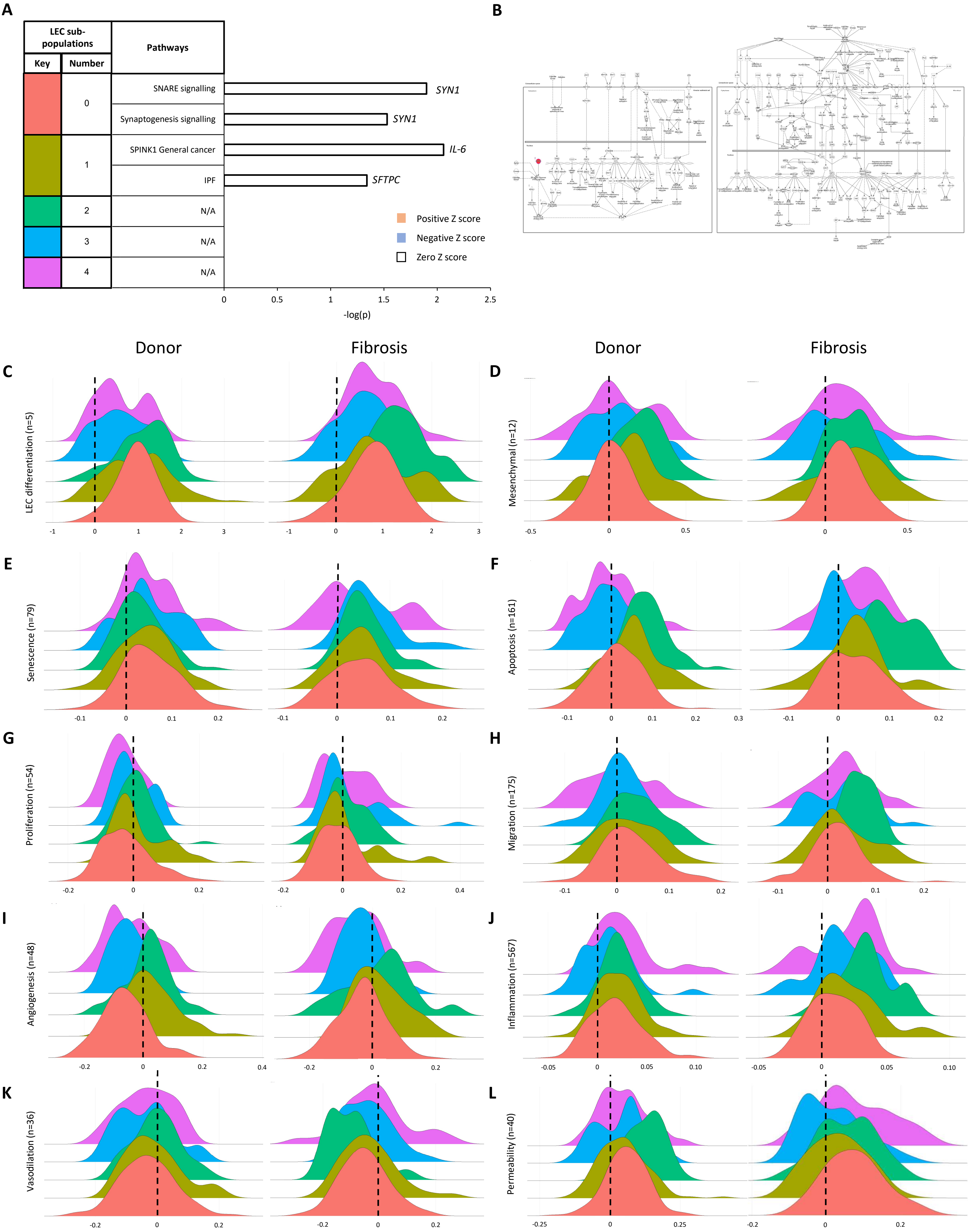

A
B
LEC sub-populations
Key
Number
Pathways
0
1
2
3
4
SNARE signalling
Synaptogenesis signalling
SPINK1 General cancer
IPF
N/A
N/A
N/A
SYN1
SYN1
IL-6
SFTPC
Positive Z score
Negative Z score
Zero Z score
0
0.5
1
1.5
2
2.5
-log(p)
Donor
Fibrosis
Donor
Fibrosis
C
D
E
F
G
H
I
J
K
L
LEC differentiation (n=5)
Mesenchymal (n=12)
Senescence (n=79)
Apoptosis (n=161)
Proliferation (n=54)
Migration (n=175)
Angiogenesis (n=48)
Inflammation (n=567)
Vasodilation (n=36)
Permeability (n=40)
