## Additional file 2: Supplementary figure legends for "Altered Heterogeneity of Ageing Lung Endothelium is a Hallmark of Idiopathic Pulmonary Fibrosis"

**Hallmark of Idiopathic Pulmonary Fibrosis**

Additional file 2: Supplemental figure legends

**Supplementary Figures**

**Figure S1. Total lung clusters and endothelial cell subclusters of cohorts 1 and 2 before integration.** *Left,* cohort 1**.** *Right,* cohort 2. **(A)** UMAP of total lung clusters. Two clusters (LEC and BEC) are labelled with a red circle. **(B)** Violin plots for endothelial cell-specific markers *VWF* (blood vessel endothelial cells), *PROX1* (lymphatic endothelial cells) and *CALCRL* (Pan-endothelial) in total lung, representing a selection of a full panel of marker produced (see Additional file 1: Detailed methods). **(C)** UMAP of total endothelium (blood and lymphatic) clusters. LEC are labelled with a green ellipses. **(D)** Heatmap of top 10 differentially expressed genes by endothelial subpopulation. **(E)** Violin plots for endothelial specific markers *VWF*, *PROX1* and *CALCRL* in total endothelium. The resolution was determined using clustree R package (Additional file 1: Detailed methods), with 0.5 value used for this analysis and throughput the study to avoid over-clustering [6]. UMAP - Uniform Manifold Approximation and Projection.

**Figure S2. Differentially expressed genes by cluster for all clusters in total lung. (A)** Violin plots for the genes which were identified as differentially expressed by cluster, grouped by cluster in whole lung. Screening these DEG (between clusters) enabled the visualization of their expression across all clusters to confirm differential expression as shown by unsupervised analysis in Seurat. **(B)** Violin plots for nFeature_RNA and nCount_RNA in the total lung clusters.

**Figure S3. Differentially expressed genes by cluster for blood vessels endothelial cells.** *Top,* Violin plots for differentially expressed genes, grouped by cluster in blood vessel endothelial cell (BEC) data set. *Bottom,* Violin plots for control housekeeping genes in BEC data set are also shown at the bottom.

**Figure S4. Expression of genes identified as differential in our analysis compared to whole ageing lung sample.** The expression of the 49 blood vessel endothelial cell (BEC) subpopulation-specific differentially expressed genes (DEG) (Figure 2D) were analysed in the total lung to determine if they were endothelial cell-specific. **(A)** Dotplot of endothelial cell marker genes (*left segment* separated by dotted line), DEG between BEC subpopulation (*central segment,* separated by dotted lines) and control housekeeping genes (*right segment,* separated by dotted line) in whole ageing lung sample. 39 (marked in red) of 49 identified subpopulation-specific DEG have not been previously assigned as markers of individual generic subpopulations of ageing human lung BEC. **(B)** Table detailing gene identities, location, and classification of 39 DEG. A pathway analysis was conducted using the ingenuity pathway analysis (Additional file 1: Detailed methods), the data from which was plotted using BioRender. **(C)** Diagram of intracellular location of BEC DEG. Adapted from “NFAT Signalling Pathway”, by BioRender.com (2022). Retrieved from <https://app.biorender.com/biorender-templates>. **(D)** From 50 identified DEG for LEC from the heatmap, 22 were observed to be ‘truly’ differential by violin plot (Additional file 2: Figure S22). From these 22, 20 were novelly expressed in LEC subpopulations. Table detailing the gene identities, location, and classification of the 20 LEC DEG. Note that this data is presented here to enable direct comparison with BEC data.

**Figure S5. Summary of differential genes by blood vessel endothelial cell subcluster.** Violin plots of differential genes, grouped by subcluster/subpopulation in blood vessel endothelial data set.

**Figure S6. Expression of markers proposed by two recent studies investigating endothelial cell heterogeneity in the lung**. Violin plots of gene markers for clusters proposed in Schupp et al., 2021*, left* and Sauler et al., 2022*, right,* in blood vessel endothelial cell data set.

**Figure S7. Marker genes proposed for blood vessel endothelial cells in six recent studies. (A)** Violin plots for marker genes proposed in six recent studies in blood vessel endothelial cell BEC) data set. **(B)** Dotplot of selected EC-sub-type marker genes across 12 identified BEC subpopulation. Genes were selected from differential genes identified from the literature with confirmation of their subpopulation specificity using violin plots. (i) [17]; (ii) [31]; (iii) [22]; (iv) [23]; (v) [25]; (vi) [24]. When applied to 12 aging human lung BEC subpopulations, the expression analysis of pre-selected by us previously reported markers of lung capillary EC in human and murine tissues [17], aerocytes [17], classical regulators of arterial and venous differentiation [18, 31-34] and large vessels [22-25].

**Figure S8. Cell-cycle scoring in blood vessel endothelial cell subclusters. (A)** UMAP of blood vessel endothelial cell (BEC) labelled by “cell-cycle score” based on results of Cell-Cycle Scoring function (Additional file 1: Detailed methods). **(B)** Pie charts comparing the cell-cycle scores in individual BEC subclusters/subpopulations split by sample condition (donor and IPF/fibrosis). **(C)** Table of statistical differences in cell-cycle scores between sample condition (donor and IPF/fibrosis) by subcluster. Statistical analysis was performed using chi- square test. P < 0.05 was considered significant. (UMAP) Uniform Manifold Approximation and Projection.

**Figure S9. Gene signatures proposed by Travaglini et al*.,* 2020.** Dot plot of signatures from Travaglini et al., 2020 in blood vessel endothelial cell data set. Art = Artery, Vn = Vein, aCAP = Aerocyte, gCAP = general capillary, Bro1/2 = Bronchial.

**Figure S10. Expression of markers proposed by two recent studies investigating lung endothelial heterogeneity. (A)** Violin plots of gene markers for endothelial cell (EC) subpopulations “sftp+” EC or “EC-pneumocyte” identified in murine lung by Rodor et al., 2021 and Bondareva et al., 2022 [26, 27]. **(B)** Violin plots of gene markers for EC subpopulation “immune-active EC” identified in porcine tissues by Wang et al., 2022 [28].

**Figure S11. Signature specificity by blood vessel endothelial cell subpopulation. (A)** Ridgeplots of average module scores for signatures across 12 subpopulations of blood vessel endothelial cells (BEC). **(B)** Table of average module scores of signatures (S1-S11) across all BEC subclusters/subpopulations. Signatures 0 and 5 could not be constructed due to a lack of differentially expressed genes (Figure 2D; Additional file 1: Detailed methods). *Coral red* represents a subpopulation with positive score (above 0) for signature, colour intensity is proportional to the score positivity. **(C)** Correlation matrix of created signatures for BEC subpopulations.

**Figure S12. Expression profile of blood vessel endothelial cell subpopulations**. **(A)** Ridgeplots for housekeeping genes: beta-2 microglobulin (*B2M),* actin-beta *(ACTB),* glyceraldehyde 3-phosphate dehydrogenase *(GAPDH)* and ribosomal protein S18 (*RPS18)* in ageing blood vessel endothelial cell (BEC) subpopulations in fibrosis samples. Black dotted line is added to annotate zero (cut-off point for differences).  **(B)** Violin plots of nFeature_RNA *left*, and nCount_RNA *right*, in BEC subpopulation. **(C)** Violin plots of cell type marker genes in BEC and total lung dataset. **(D)** Violin plots of endothelial cell progenitor cell marker genes in BEC subpopulation.

**Figure S13. Identification, percentage contribution and transcriptional profiles of blood vessel endothelial cell subpopulations in lungs from young and old donors. (A-C)** The same colour key for blood vessel endothelial cells (BEC) subclusters/subpopulations is used for all three figures. **(A)** UMAP of BEC from integrated total lung map of four old (49-66 years; donor dataset from cohort 1; Table 1; [27]) and three young (21-29 years; see Additional file 1: Detailed methods; [27]) donors, identifying 13 subpopulations (A-M). **(B)** UMAP of young and old BEC split by age of donors (young and old). **(C)** Table detailing number (n) or percentage of cells per subpopulation from total BEC in lung tissue (%; graphically presented on a stacked bar chart on the *right*) split by age of donors (young and old). **(D)** Heatmap of top 10 differentially expressed genes by BEC subpopulation in integrated dataset (see **A**).

**Figure S14. Comparison of transcriptional profiles of lung blood vessel endothelial cells subpopulations between young and old donors.** The analysis of integrated scRNAseq dataset of human lung tissues from four old and three young donors (see Figure S13 legend and Additional file 1: Detailed methods; [27]) was performed to test the specificity of our novel transcriptional signatures (Figures 4A, S11).  **(A)** Dot plot representing expression of genes from signatures of ageing human blood vessel endothelial cells (BEC) subpopulations (see integrated dataset for both donors and IPF patients; Figure 2D) in integrated (young and old) dataset. **(B)** Dot plot of marker genes proposed for lung BEC in six recent studies (Figure S7; for more details see Additional file 1: Detailed methods) in integrated (young and old) dataset. **(C-D)** Ridgeplots of average module scores for transcriptional signatures (Figures 4A, S11) across all clusters in **(C)** young and **(D)** old donor lung dataset. Sig 1 = signature 1 etc. **(E)**  Tables of average module scores for all 10 signatures split by young (*left*) and old (*right*) datasets. S1 = Signature 1 etc. *Coral red* represents a subpopulation with positive score (above 0) for signature, colour intensity is proportional to the score positivity. **(F)** Correlation matrices for proposed signatures in lung datasets from young (*left*) and old (*right*) donors. This data served as a “proof-of-principle” of the specificity of our novel transcriptional signatures. For full details of analysis please see Additional file 1: Detailed methods. UMAP -Uniform Manifold Approximation and Projection.

**Figure S15. Spatial localisation of endothelial cells within IPF lung tissue regions.** Ridgeplots for **(A)** *PECAM1* and **(B)** *CALCRL* expression in 12 ageing blood vessel endothelial cell (BEC) subpopulations in IPF/fibrosis samples from total integrated object (cohort 1 and 2). Note that subpopulations 0 and 5 have similar expression levels of the housekeeping gene beta-2 macroglobulin (*B2M*; Figure S12A) and comparable to all other ageing human lung BEC subpopulations pan-EC markers’ (*PECAM1*/CD31 and *CALCLR*/CLR) expression profiles (ridgeplots), but considerably larger proportion of cells with lower expression of these genes (this figure). **(C)** Scatter plots showing correlation between *PECAM1* expression and *CALCRL* expression in ageing BEC subpopulations in IPF samples from cohorts 1 and 2. **(D)** Scanned total section of formalin fixed paraffin embedded distal lung parenchyma tissue stained with Masson’s Trichrome (*left*), CD31 (clone JC/70A; *middle*) and CLR (LN1436 [43]; *right*). Red squares represent regions of interest (ROI) detailed in **(E)**. Scale bar represents 5000µm. **(E)** Five regions of interest (ROI) stained with Masson’s Trichrome (*left*), CD31 (*middle*) and CLR (*right*). *Black arrows* label the identifiable ageing BEC subpopulations in ROI. Scale bar represents 100µm. *Far right,* summary of identifiable subpopulations of ageing BEC subpopulations within ROI. Note that subpopulations 0 and 5 could not be identified due to low expression of pan-endothelial markers, whilst subpopulation 1 is not present in distal lung (Figure 3D) and specific location of subpopulation 9, 10 and 11 is unknown.

**Figure S16. Images used for quantification of ageing blood vessel endothelial cell subpopulations in peri-bronchial areas. (A)** Scanned total section of formalin fixed paraffin embedded distal lung parenchyma tissue stained with Masson’s Trichrome (*left*) and CLR (LN1436, [43]; *right*) and used for quantification of ageing blood vessel endothelial cell subpopulations in peri-bronchial areas (Figure 5C-E). Red squares represent regions of interest (ROI) detailed in **(B)**. Yellow squares represent regions of interest (ROI) detailed in **(C)**. Scale bar represents 5000µm. **(B)** Nine regions of interest (ROI) termed ‘thin’ for analysis, stained with Masson’s Trichrome (*left*) and CLR (*right*). Scale bar represents 100µm. **(C)** Three regions of interest (ROI) termed ‘thick’ for analysis, stained with Masson’s Trichrome (*left*) and CLR (*right*)*.* Scale bar represents 100µm.

**Figure S17. Identifiable bronchi used for quantification of blood vessel endothelial cell subpopulations in peri-bronchial areas in ageing lung from IPF patient. (A, C)** Scanned total sections of formalin fixed paraffin embedded distal lung parenchyma tissues S1 and S2 (from two blocks of tissue from the same patient) were immunostained using CLR antibody (LN1436 [46]) and used for quantification of ageing blood vessel endothelial cell subpopulations in peri-bronchial areas (Figure 5 F-I). Red squares represent regions of interest (ROI) detailed in **(B, D).** Scale bar represents 5000µm. **(B)** Four and **(D)** nine ROI used for analysis in S1 and S2 respectively*.* Scale bar represents 100µm.

**Figure S18. Images used for quantification of blood vessel endothelial cell subpopulations in alveolar regions in ageing lung from IPF patient. (A)** Scanned total section of formalin fixed paraffin embedded distal lung parenchyma tissue immunostained using CD31 antibody (clone JC/70A, see Additional file 1: Detailed Methods). Red squares represent regions of interest (ROI) labelled “low degree fibrosis” and detailed in (B)**.** Scale bar represents 5000µm. **(B)** Images of nine “low degree fibrosis” (1-9; *left*) and nine “high degree fibrosis” (10-18; *right*) ROI used for high content image analysis (Figure 6F-H)*.* Scale bar represents 100µm.

**Figure S19. Differentially expressed genes in blood vessel endothelial cell subpopulations between donor and fibrosis and their association with IPF signalling pathways. (A)** Volcano plots for differentially expressed genes (DEG) (donor vs IPF/fibrosis) in blood endothelial cell (BEC) subpopulations. Statistical analysis was done using Shapiro Wilcoxon test and Wilcoxon rank sum test, with P value <0.05 (dotted line indicates cut-off) considered significant. **(B)** Schematic of IPF signalling pathways identified using Ingenuity Pathway Analysis and DEG from subpopulations zero, 8 and 11. Highlighted in *red* genes in figure represent DEG which were statistically significantly linked to the indicated signalling pathway (for details, see Additional file 1: Detailed Methods).

**Figure S20. Expression profiles of gene sets relevant to endothelial cell biology processes in blood vessel endothelial cell subpopulation in donors and IPF patients.** Published gene set enrichment analysis (GSEA) libraries were utilized for the assessment of expression of marker genes associated with ten selected key/relevant to endothelial cell (EC) biology processes. Samples were assigned module scores using the Seurat function AddModualScores based on genes used on GSEA website (Further details are available in Additional file 1: Detailed methods, including scoring and interpretation. Full gene lists are available in Additional file 3: Table S7). **(A-K)** Key colour codes for subpopulations are the same as in Figures 2-4. **(A-J)** Histograms (ridgeplot) of EC differentiation (n= 5), endothelial-mesenchymal transition (Endo-MT) (n= 12), senescence (n= 79), apoptosis (n= 161), proliferation (n= 54), migration (n= 175), angiogenesis (n= 48), inflammation (n= 567), vasodilation (n= 36) and permeability (n= 40) scores in all blood vessel endothelial cell (BEC) subpopulations in donor and fibrosis. Number of genes in sets are indicated in brackets. Crucially, histograms show the distribution of a score across BEC subpopulations (thus reflecting the heterogeneity of gene expression in individual cells within each individual subpopulation) compared to the total cluster/population. **(K)** Table of average module scores per subpopulation. *Coral red* represents a subpopulation with positive score (above 0) for signature, colour intensity is proportional to the score positivity. Note that scRNAseq data reveals that eleven subpopulations in the ageing human lung have positive senescence scores in donor lung (**C, K**; and only for two of them - subpopulations 5 and 8 - the score further changes in IPF; Figure 7E), with distinct ridgeplot shifts detected in four subpopulations (3, 6, 8 and 9 – marked with *arrows*, **C**). Subpopulations 3, 6 and 8 reside in alveolar regions of the ageing human lung, whilst localisation of subpopulation 9 remains to be confirmed. **(L-M)** To assess the presence of senescent endothelium in the alveolar regions of the ageing human lung, we performed immunohistochemistry (using antibody against pan-endothelial marker CLR (LN1436 [46]), allowing for localisation of individual subpopulations in various regions of the lung; see Figure S15) and immunohistochemistry (for senescence using Sudan Black B; see Additional file 1: Detailed methods; [S32]) using formalin fixed paraffin embedded (FFPE) tissue section of distal lung diagnostic biopsy of IPF (with confirmed histopathological pattern of usual interstitial pneumonia, including signs of developing fibrosis and reflecting intra-lung heterogeneity observed clinically) from 58 years old subject. **(L)** Representative images of alveolar region of interest (ROI) recorded from serial sections immunostained for CLR (*left;* DAB, brown colour) or Sudan Black B and Nuclear Fast Red (*right;* SBB+NFR, grey and pink colours). Scale bar is 100um. Red boxes show regions enlarged for presentation in (**M**). **(M)** Enlarged region stained with CLR antibody LN1436 (*left;* DAB, brown colour) or Sudan Black B and Nuclear Fast Red (*centre;* SBB+NFR, grey and pink colours) or NFR only (*right;* NFR, pink colour). Scale bars are 50um. Blue arrow indicates senescent BEC venule (grey colour) in alveolar region, corresponding to subpopulation 6. Note that structures resembling capillary endothelial cells in alveolar region, (corresponding to subpopulations 3 and 8) also have grey colour, although resolution is insufficient for making a conclusion about these subpopulations.

**Figure S21. Cell subtype-specific composition of aging human lung lymphatic endothelial cell cluster from donors and IPF patients from two independent cohorts.** Ageing lymphatic endothelial cell (LEC) sub-type-specific signatures were generated in a similar manner to blood vessel endothelial cell (BEC) subpopulation analysis (Figure 2 A-D; Additional file 1: Figure S11 A, B), using identified DEG and expression of known lymphatic genes (Figure 8 A-D) and were tested for their specificity by comparing cell subpopulations for their similarity to each subpopulation. **(A)** Ridgeplots of developed signatures across 5 identified LEC subclusters/subpopulations. **(B**) Table of average module scores for proposed signatures by subcluster. *Coral red* represents a subpopulation with positive score (above 0) for signature, colour intensity is proportional to the score positivity. **(C)** Ridgeplots for housekeeping genes; *B2M, ACTB, GAPDH* and *RPS18* in ageing LEC subpopulations in fibrosis samples. **(D)** Dendrogram (cluster tree) and dot plot of lymphatic subpopulations, based on unsupervised hierarchical clustering of 5 subpopulations of ageing human lung LEC from integrated total dataset, which was split into two separate objects by condition (donor on the *left* and fibrosis on the *right*). **(E)** Dot plots of genes identified as differentially expressed by subpopulation from the heatmap (Figure 8B), plotted alongside pan-EC and LEC markers and housekeeping (control) genes for comparison. **(F)** Violin plots for the expression on PDPN and CCL21 in ageing LEC subpopulations in fibrosis.

**Figure S22. Differentially expressed genes by cluster for lymphatics.** *Left,* violin plots for the genes which were identified as differentially expressed by subcluster in lymphatic endothelial cell (LEC) data. *Right,* violin plots for housekeeping (control) genes by cluster in LEC data.

**Figure S23. Cell-cycle scoring in lymphatic endothelial cell subclusters. (A)** UMAP of lymphatic vessel endothelial cell (LEC) labelled by “cell-cycle score” based on results of Cell-Cycle Scoring function (Additional file 1: Detailed methods) utilising scRNAseq/ transcriptional profiles. **(B)** Pie charts comparing the cell-cycle scores in individual LEC subclusters/subpopulations split by sample condition (donor and IPF/fibrosis). **(C)** Table of statistical differences in cell-cycle scores between sample condition (donor and IPF/fibrosis) by subcluster. Statistical analysis was performed using chi- square test. P < 0.05 was considered significant. (UMAP) Uniform Manifold Approximation and Projection.

**Figure S24. Proportions of lymphatic endothelial cell subpopulation in lung tissues from donors and IPF patients**. **(A)** UMAP of lymphatic endothelial cell (LEC) split by sample condition (donor and IPF/fibrosis). **(B)** Table detailing number (n) or percentage of cells per subpopulation from total LEC in lung tissue (%; graphically presented on a stacked bar chart on the *right*) split by condition (donor or IPF/fibrosis). **(C)** UMAP of LEC total population split by sample condition (donor or IPF/fibrosis) and labelled by cohort (1 and 2). **(D-E)** Stacked bar chart of percentage contribution of each LEC subcluster to total LEC population by sample condition (donor or IPF/fibrosis) for cohorts 1 **(D)** and 2 **(E)**. (UMAP) Uniform Manifold Approximation and Projection.

**Figure S25. Identifiable lung structures used for quantification of podoplanin-positive ageing lymphatic endothelial cell subpopulations in IPF lung. (A-B)** Immunostaining for podoplanin (PDPN, clone D2-40) was conducted on formalin fixed paraffin embedded (FFPE) tissue sections of distal lung diagnostic biopsy of IPF (with confirmed histopathological pattern of usual interstitial pneumonia) from 58 years old subject (for full details, see Additional file 1: Detailed methods). **(A)** Scanned total section. Red squares represent regions of interest (ROI), which are presented in (**B**). Scale bar represents 5000µm. **(B)** Images of 21 ROI used for high content image analysis (Figure 8H). Scale bars represent 100µm. A - arteries, V - veins, arrow heads - small vessels, stars - cysts, B - bronchioles and P - pleura.

**Figure S26. Association of differentially expressed genes in ageing human lung lymphatic endothelial cell with specific signalling pathways and endothelial cell-relevant processes in IPF. (A)** Bar chart detailing the results of the Ingenuity Pathway Analysis (IPA) using Core expression analysis function [77] (Additional file 1: Detailed methods) of statistically significant altered pathways using differentially expressed genes (DEG) (donor vs IPF/fibrosis) in lymphatic endothelial cell (LEC) (Figure 8I; Additional file 3: Table S10). *Blue colour* represents a negative Z score (down), *orange* represents a positive Z score (up) and *white* represents a Z score of zero. DEG from three subpopulations (2, 3 and 4) did not significantly associate with any pathways, and therefore bars are absent. **(B)** IPF signalling pathways identified using IPA in (**A**). **(C-L)** Published gene set enrichment analysis (GSEA) libraries were utilized for the assessment of expression of marker genes associated with ten selected key/relevant to endothelial cell (EC) biology processes. Samples were assigned module scores using the Seurat function AddModualScores based on genes used on GSEA website (Further details are available in Additional file 1: Detailed methods, including scoring and interpretation. Full gene lists are available in Additional file 3: Table S7). Key colour codes for subpopulations are the same as in (A) and in Figure 8. Histograms (Ridgeplot) of LEC differentiation (n= 5), endothelial-mesenchymal transition (Endo-MT) (n= 12), senescence (n= 79), apoptosis (n= 161), proliferation (n= 54), migration (n= 175), angiogenesis (n= 48), inflammation (n= 567), vasodilation (n= 36) and permeability (n= 40) scores in all LEC subpopulations in donors (*Donor*) and IPF (*Fibrosis*). Number of genes in sets are indicated in brackets. Crucially, histograms show the distribution of a score across LEC subpopulations (thus reflecting the heterogeneity of gene expression in individual cells within each individual subpopulation) compared to the total cluster/population. For full details see Additional file 1: Detailed Methods. Statistical analysis was in the form of Shapiro Wilcoxon test and Mann-Whitney U test (see Figure 8J).
