## Additional file 3: Supplementary table legends for "Altered Heterogeneity of Ageing Lung Endothelium is a Hallmark of Idiopathic Pulmonary Fibrosis"

Eamon C. Faulkner, Adam A. Moverley, Simon P. Hart and Leonid L. Nikitenko

Supplemental tables

**Supplementary Tables**

**Table S1**. Differentially expressed genes between clusters in total lung.

**Table S2**. The comparison of identified total lung clusters in the present study to 6 recent single cell RNA sequencing studies [1, 2, 6, 20, 35 and 36].

**Table S3.** Quantification of cell numbers and percentage of each cluster separated by cohort and sample condition of whole lung.

**Table S4.** Correlation matrix of differentially expressed gene signatures from heatmap of blood endothelial cells (Figure 2B). For more details on signature creation see section Annotation of BEC Subpopulation. S1 = Signature 1.

**Table S5.** Quantification of the mean number of cells in each subpopulation of blood endothelial cells per sample split by cohort and condition.

**Table S6.** Quantification of cell numbers and percentage of each cluster separated by cohort and sample condition of blood endothelial cells.

**Table S7.** Differentially expressed genes identified in blood endothelial cells between donor and fibrosis for each cluster, which were later used for IPA.

**Table S8.** Gene libraries used for ‘scoring’.

**Table S9.** Summary table of cell numbers and percentage of each cluster separated by cohort and sample condition of lymphatic endothelial cells.

**Table S10.** Differentially expressed genes identified in lymphatic endothelial cells between donor and fibrosis for each cluster, which were later used for IPA.
